# Association between *Plasmodium falciparum Kelch13* mutations and malaria parasite clearance half-life after artemisinin-based therapy: an updated WWARN systematic review and individual patient data meta-analysis

**DOI:** 10.64898/2026.09.08.26362414

**Authors:** Stephanie van Wyk, K13 Genotype-Phenotype Study Group, Philip J Rosenthal, Philippe Guerin, Mehul Dhorda, Karen Barnes, Prabin Dahal

## Abstract

**Background:** Artemisinin-based combination therapies remain the first-line treatment for uncomplicated *Plasmodium falciparum* malaria. *Plasmodium falciparum Kelch13* mutations emerge and spread within distinct malaria epidemiological and immunological contexts, shaping the expression of artemisinin resistance (ART-R). Understanding the clinical phenotypes of these mutations requires evaluation across diverse transmission settings and geographic regions.

**Methods:** A systematic review (SR) and individual patient data meta-analysis (IPDMA) were conducted (PROSPERO: CRD42019133366) to identify studies that included serial parasite density measurements and *Kelch13* genotyping. Associations between *Kelch13* mutations and parasite clearance half-lives (PC1/2) and Day 3 parasite positivity were assessed. Receiver operating characteristic (ROC) analyses identified PC1/2 thresholds most strongly associated with relevant *Kelch13* mutations.

**Findings:** The SR identified 86 eligible studies, and individual patient data were obtained from 45 studies (n=16,823 patients from 539 study sites in 33 countries). After excluding hyperparasitaemia, Day 3 parasite positivity exceeded 10% overall across all WHO-validated, candidate, and potential mutations, but not for other *Kelch13* mutations. Parasite clearance rates were consistently shorter in moderate-to-high-transmission than in lower-transmission areas. The WHO threshold PC1/2>5h had a sensitivity of 36% (27.9-44.6) for detecting WHO-validated mutations in moderate-to-high transmission areas and 81% (78.5-82.8) in lower transmission areas. In moderate-to-high transmission settings, a PC1/2 threshold of 3.1h optimally discriminated WHO-validated mutations (ROC AUC 0.78; 95% CI 0.74–0.82; sensitivity 75% (95% CI:67%-82%); specificity 71%, 95% CI: 69%-73%). Additional emerging *Kelch13* mutations associated with delayed parasite clearance were identified in relatively small African and Asian sample sets.

**Interpretation:** Compared with WT parasites, parasites carrying WHO-validated ART-R mutations were associated with a 34%-59% longer mean PC1/2, regardless of endemicity, treatment regimen, or age. Given the low sensitivity of the PC1/2>5h threshold for identifying WHO-validated ART-R parasites in moderate-to-high transmission settings, where its recalibration is indicated. Broader genotyping is warranted for identifying emerging *Kelch13* mutations associated with delayed parasite clearance.

**Research in Context: Panel:** *Evidence before this study:* Artemisinin-based combination therapies (ACTs) remain the first-line treatment for *Plasmodium falciparum* malaria. The emergence and spread of artemisinin partial resistance (ART-R), characterised by delayed parasite clearance after treatment and mediated primarily by non-synonymous mutations in the *Kelch13* propeller domain, pose a major threat to global malaria control. Previous individual-patient data meta-analyses (IPDMAs) have established an association between *Kelch13* mutations and delayed clearance, but these analyses were largely restricted to low-transmission settings in Southeast Asia (SEA). Comprehensive evaluation of these relationships in Africa, which bears 95% of the global falciparum malaria burden and where transmission intensity, host immunity, and infection complexity differ, is needed.

*Added value of this study:* This study represents the largest and most geographically comprehensive IPDMA of *Kelch13*-associated ART-R to date, integrating published and unpublished data from 16,823 patients at 539 sites across 33 countries, including 22 in Africa. Extending on the previous analysis and incorporating the newly updated WHO compendium of relevant *Kelch13* mutations, this study evaluated parasite clearance phenotypes across diverse transmission intensities, including underrepresented African populations. WHO-listed mutations of ART-R were strongly associated with Day 3 positivity and prolonged parasite clearance half-life (PC1/2) after treatment, while additional *Kelch13* mutants were also associated with delayed clearance, thereby expanding the contemporary molecular landscape of ART-R. The WHO-recommended PC1/2 threshold of >5h showed low sensitivity for detecting ART-R in moderate-to-high transmission settings. Our findings support transmission-specific interpretation of PC1/2 and indicate that a threshold of 3.1 h more accurately identifies WHO-validated ART-R mutations in moderate-to-high transmission settings, which bear the greatest global malaria burden.

*Implications of all the available evidence:* ART-R is now established across much of SEA and is emerging independently in multiple African parasite populations. Building on the previous WorldWide Antimalarial Resistance Network (WWARN) IPDMA and the expanding WHO evidence base on drug-resistance molecular markers, this study strengthens the evidence linking *Kelch13* mutations to delayed parasite clearance following treatment in Africa, while demonstrating that the phenotypic expression of ART-R varies with transmission intensity and epidemiological context. These findings support context-specific interpretation of parasite clearance metrics, integrated with molecular surveillance and therapeutic efficacy studies, to improve early detection of emerging ART-R and guide timely public health responses.

## Introduction

Despite being preventable and treatable, malaria remains a major global health problem, with 282 million cases and 610,000 deaths (2024) (1). Effective case management with artemisinin-based combination therapies (ACTs) has reduced this burden, which has formed the foundation of treatment for uncomplicated *Plasmodium falciparum* malaria in nearly all endemic countries, scaled-up after 2006 (2, 3). However, the efficacy of ACTs is now under increasing threat due to the emergence and spread of parasites with artemisinin partial resistance (ART-R) (4–6).

ART-R was first reported in Southeast Asia (SEA) in 2008 (4, 7) and is now a growing concern in Africa, which bears 95% of the malaria burden (1). The independent emergence of ART-R has been confirmed in parts of East Africa and the Horn of Africa (1, 8). Recent reports of WHO-candidate ART-R mutations from southern Africa, including Namibia and Zambia, have raised concern regarding the spread of ART-R across the continent (1, 9–11). However, much of the current understanding is derived from ART-R parasite populations in SEA (12) and knowledge of the emergence, spread, genetic epidemiology, and clinical implications of ART-R in Africa remains limited (3, 8, 13). Characterisation of ART-R is complicated by differences in transmission intensity and host immunity between Africa and SEA. Much of the existing evidence base has been derived from low-transmission settings in SEA, where delayed parasite clearance more directly reflects parasite genotype (14). In contrast, many African settings are characterised by moderate-to-high transmission intensity, greater heterogeneity in acquired immunity, higher complexity of infection, and distinct epidemiological patterns that may influence parasite clearance dynamics and the clinical expression of ART-R (8,13). Moreover, in SEA, the emergence of ART-R was soon compounded by resistance to longer-acting partner drugs and declining ACT efficacy. A similar trajectory in high-burden regions of Africa could lead to substantial increases in malaria-related morbidity and mortality (8, 13, 15).

The interpretation of ART-R requires integrating parasite genotyping and clinical phenotyping across transmission settings and regions to assess the clinical relevance of specific *Kelch13* mutations. The clinical phenotype of ART-R is defined by delayed parasite clearance following artemisinin-based treatment and is primarily associated with mutations in the *Kelch13* propeller domain (12, 16, 17). The WHO periodically updates the *Kelch13* compendium, which classifies mutations according to the strength of evidence linking them to ART-R phenotype (18, 19). The parasite clearance half-life (PC1/2), the log-linear decline in parasitaemia following treatment initiation, is widely regarded as the most robust *in vivo* indicator of artemisinin susceptibility (14), with a threshold of >5h used to define delayed clearance (14, 20). Persistent parasitaemia on Day 3 post-treatment is a pragmatic alternative metric for assessing delayed clearance. These complementary phenotypic measures can be evaluated relative to baseline *Kelch13* genotypes to gauge ART-R status. Integrating molecular and clinical data is therefore essential for identifying emerging ART-R-associated mutations, validating surveillance markers, and understanding the evolving dynamics of ART-R across diverse settings.

Much of the current understanding of *Kelch13-*associated ART-R is derived from investigations conducted almost a decade ago (12). This may not fully reflect contemporary parasite populations, treatment landscapes, or the diversity of transmission settings in which ACTs are currently deployed. Therefore, we set out to conduct an updated evaluation using newly available data to characterise the current dynamics of *Kelch13-*associated ART-R and their clinical implications. This represents the most comprehensive evaluation of *Kelch13*-associated ART-R to date, integrating molecular and clinical data across the broadest geographical range investigated to date.

## Methods

We collated and harmonised individual patient data (IPD) from diverse malaria-endemic regions to investigate the relationship between *Kelch13* mutations and treatment outcomes by evaluating PC1/2 (primary outcome) and Day 3 parasite positivity (secondary outcome) using a unified statistical framework aligned with the PRISMA IPDMA standards (21).

### Literature review and screening

We conducted a systematic literature search and screening in accordance with the PRISMA-IPD guidelines (PROSPERO CRD42019133366). The searches included published articles and unpublished clinical trials across bibliographic databases and data repositories (21), with coverage from 1 January 2014 to 31 August 2024 (12). Eligible studies included prospective trials of uncomplicated *P. falciparum* malaria with at least one arm receiving an artemisinin-based treatment as an ACT or monotherapy, reporting patient characteristics and clinical phenotype (PC1/2 or Day 2 or 3 positivity), and providing baseline *Kelch13* genotypes and antimalarial dosing information (21).

### Data acquisition and harmonisation

Principal investigators of eligible studies were invited to contribute IPD to the WWARN *Kelch13* study group, which is managed by the Infectious Diseases Data Observatory (IDDO) data platform. Data were collated and harmonised into a Clinical Data Interchange Standards Consortium (CDISC)-compliant format, using the IDDO study data tabulation model (SDTM), de-identified and pseudonymised.

### Statistical analyses

Statistical analyses followed a prespecified, published statistical analysis plan (21). We investigated the proportion of Day 2 and Day 3 positivity, defined as detectable asexual parasitaemia at 48h and 72h (±6h), respectively, after treatment initiation. Parasite positivity was analysed using multivariable logistic regression with a random intercept for study site and fixed effects for baseline parasitaemia, transmission intensity, and age.

The PC1/2 was estimated for each patient using their parasitaemia-time profile (12, 21). The average multiplicative effect of carrying a mutation on the prolongation of the PC1/2 was estimated for each mutation using a linear regression of log-transformed half-life, and the effect size was presented with 95% confidence interval (95% CI).

We performed a receiver operating characteristic (ROC) curve analysis to identify a data-driven PC1/2 threshold for discriminating WHO-validated mutations from wild-type (WT) parasites; this approach assigns equal weight to both sensitivity and specificity.

Diagnostic performance of Day 2 and Day 3 positivity in identifying delayed parasite clearance (defined as PC1/2>5h) was assessed by estimating sensitivity, specificity, and corresponding 95% CIs, both overall and stratified by transmission intensity, region, and baseline hyperparasitaemia. To contextualise the WHO >10% Day 3 positivity threshold, the predicted probability of Day 3 parasite positivity was modelled as a function of PC1/2 overall and stratified by transmission setting.

The diagnostic accuracy of the different PC1/2 thresholds was presented as the area under the ROC curve (AUROC), with 95% CI, along with sensitivity and specificity. This was undertaken separately for transmission settings: low-to-very-low and moderate-to-high (defined in (21)). PC1/2 was modelled using a linear regression with a random intercept for study site and fixed effects for age and baseline parasitaemia.

All analyses used a one-stage IPDMA approach, and all prespecified analyses were implemented in R and/or Stata version 18.

### Risk of bias assessment

Risk of bias in studies included was assessed based on the following three domains: i) genotyping strategy used (i.e. if whole gene/propeller region genotyped or only specific mutations genotyped), ii) the frequency of serial parasite sampling, and iii) the number of parasitaemia-time profiles excluded for unsatisfactory fit for PC1/2 estimation.

## Results

### Literature search

The literature search identified 381 articles and clinical trials published between 2014 and 2024, of which 86 met the inclusion criteria. We requested IPD from all these studies and received data from 45 studies (30 additional studies to complement previous analyses (12), **Supplementary file 1**).

### Description of the study sites

The 45 studies were conducted in 33 countries, including 22 in Africa, nine in Asia, and three multicentre, multi-country studies spanning both regions **(Supplementary file 1)**. In total, the dataset comprised 539 study sites and 16,823 individual patient records (**Figure 1**).

**Figure 1:**
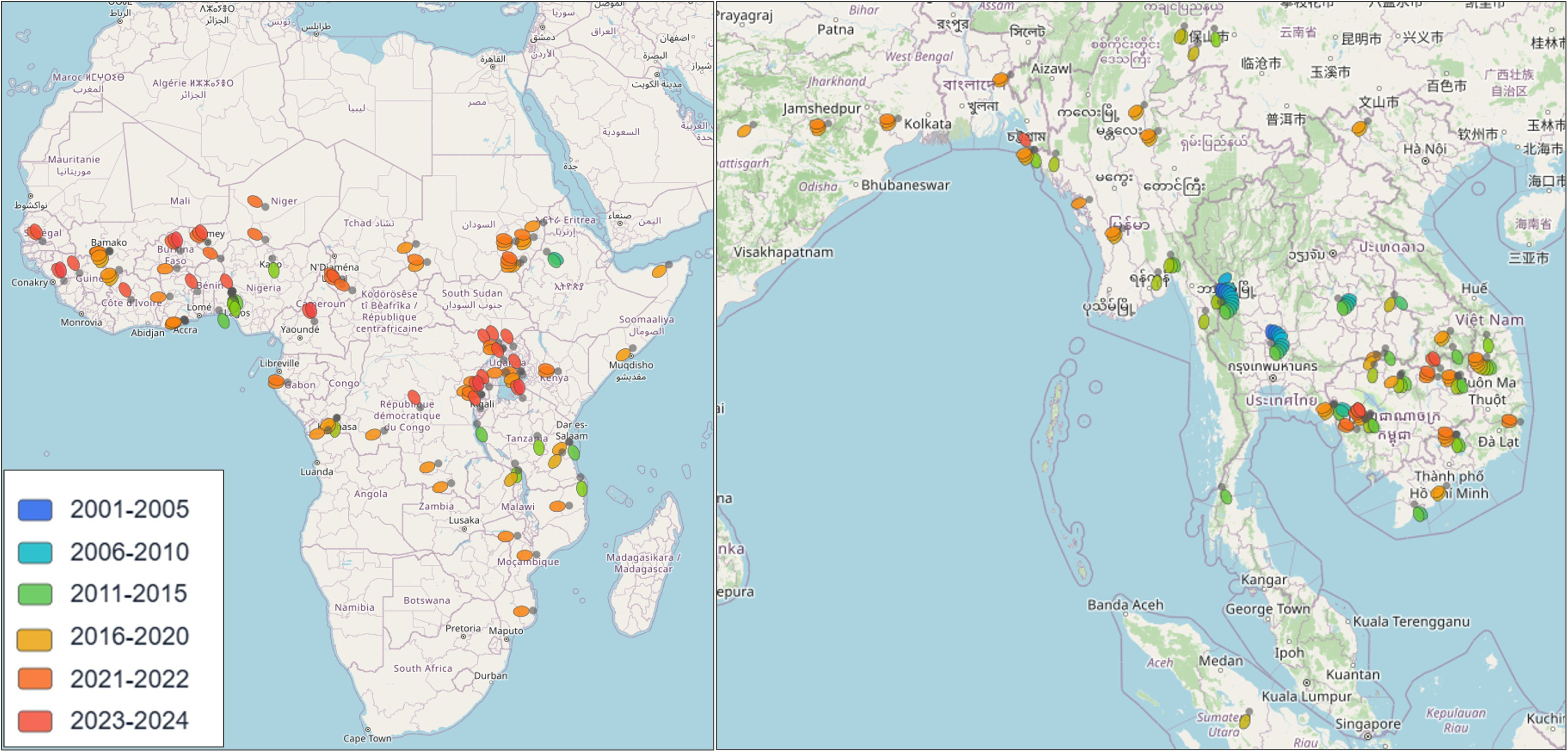
Geographical distribution of included study sites across Africa and Asia, coloured by study period.

### Baseline characteristics

Complete records of post-treatment parasitological outcomes, baseline genotypic data, and key demographic variables were available for 12,023 unique participants across 42 studies (Patient characteristics and treatment distributions are summarised in **Supplementary file 1**). Estimates of PC1/2 were available for a subset of 7,559 patients across 22 studies, with sufficient longitudinal parasite measurements to model parasite clearance. Of these, 2,953 patients were from 9 studies conducted in Africa, 4,604 were from 16 studies conducted in Africa, and 2 had unknown geography.

The median age of African patients was 6 years [IQR: 3-9.1], and 12% (1,103/6,658) were >12 years; the median age of Asian patients was 22 years [IQR: 14-32], and 81% (4,615/5,363) were >12 years. The median age decreased with increasing transmission intensity, from 25 years (2,150) in very low (21, 22) to 14 years (803) in low, to 5 years (3,201) in moderate, to 4 years (1,362) in high transmission settings. Overall, 62% (7,656/12,294) were male.

Artemether–lumefantrine was the most used treatment (28% [3,493/12,294]), with higher use in Africa (44% [2,925/6,658]) than in Asia (10% [568/5,636]), while dihydroartemisinin–piperaquine was more widely used in SEA (25% [1,423/5,636]) and very low transmission settings (35% [996/2,812]) (**Table 1**).

**Table 1:**
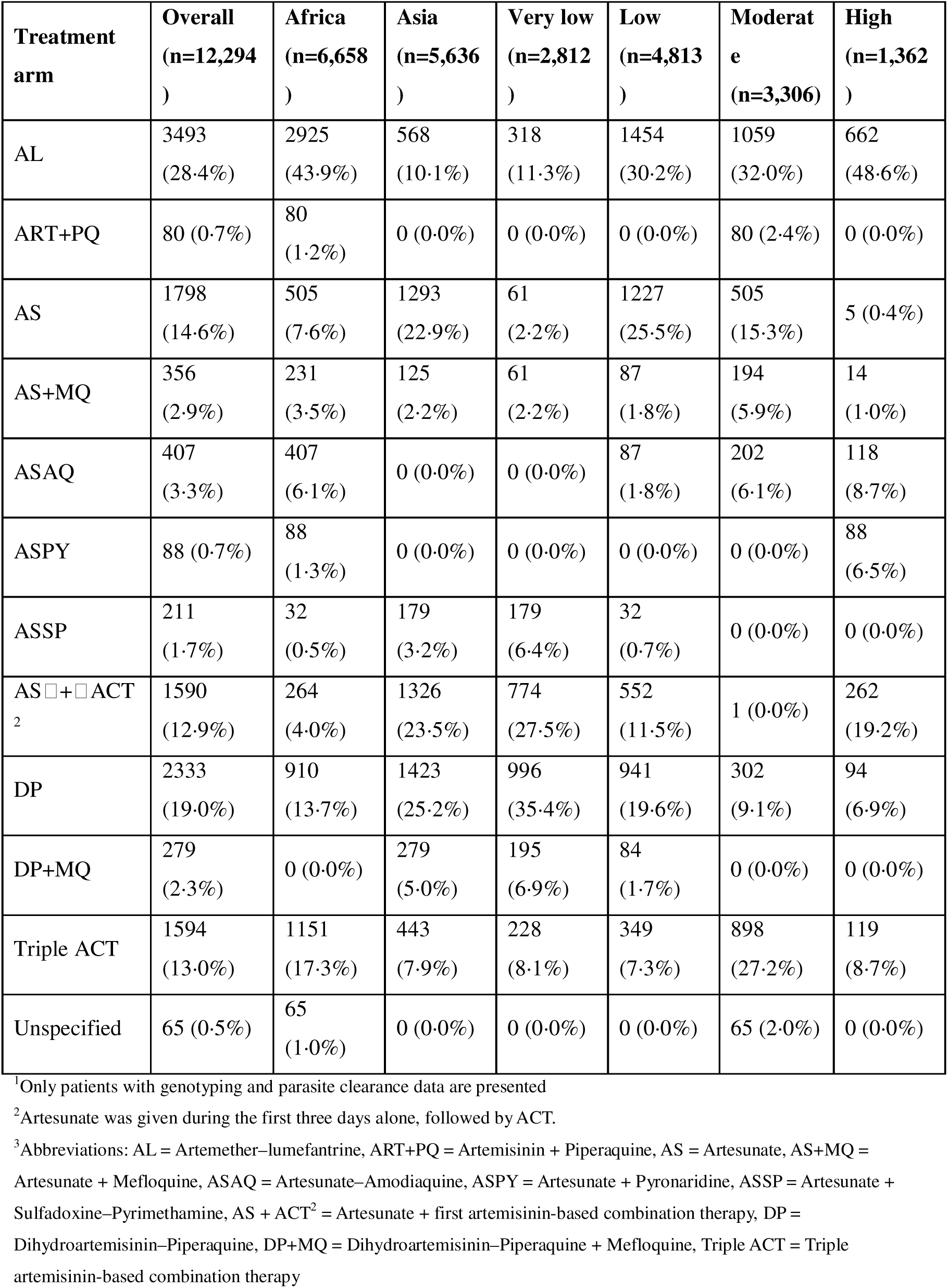
Summary of treatments received by participants given per transmission setting and geographical region (Africa or Asia)^1,2,3^.

### Molecular analyses of *Kelch13*

At baseline, 9,382 retrieved samples were *Kelch13-*WT, and 60 distinct non-synonymous mutations were identified. The dataset constituted 2,541 patients with WHO-listed mutations (potential [64], candidate [209], and validated [2,268]) and 371 other unassigned mutations; counts are not mutually exclusive, as some patients had multiple mutations within a single sample.

*Kelch13* mutant genotypes were more common in Asian samples (42% [2382/5,636]) than in Africa samples (8% [527/6,659]). For WHO-listed mutations in Africa, most samples were collected from countries where ART-R was not confirmed or suspected (86% [221/226]), whereas in SEA, most were collected in countries that had confirmed ART-R (70% [1598/2,275]) or suspected ART-R (19% [421/2,275]).

### Day 3 Parasite Positivity

Day 2 and 3 positivity were reported in 67% (5,870/8,814) and 21% (1,666/7,588) of patients, respectively, and varied across regions and transmission settings (**Supplementary files 2–3**). Day 2 and Day 3 positivity were consistently higher in parasites carrying WHO-listed mutations (validated, candidate, and potential) than in WT or other mutations (**Figure 2a-b**). Day 3 positivity was observed in 70% (1,126/1,602) of patients with WHO-validated mutations, 74% (86/116) with WHO-candidate mutations, and 82% (37/45) with WHO-potential mutations, compared with 14% (50/371) with other *Kelch13* mutations and 6% (316/5,475) with WT infections. Excluding participants with baseline hyperparasitaemia (defined in **Supplementary files 3** and (21)) reduced positivity estimates across all mutation classes with a threshold of 10% prevalence overall, seen only with WHO-validated, candidate and potential mutations, but not other unassigned *Kelch13* mutations (**Figure 2c-d**).

**Figure 2:**
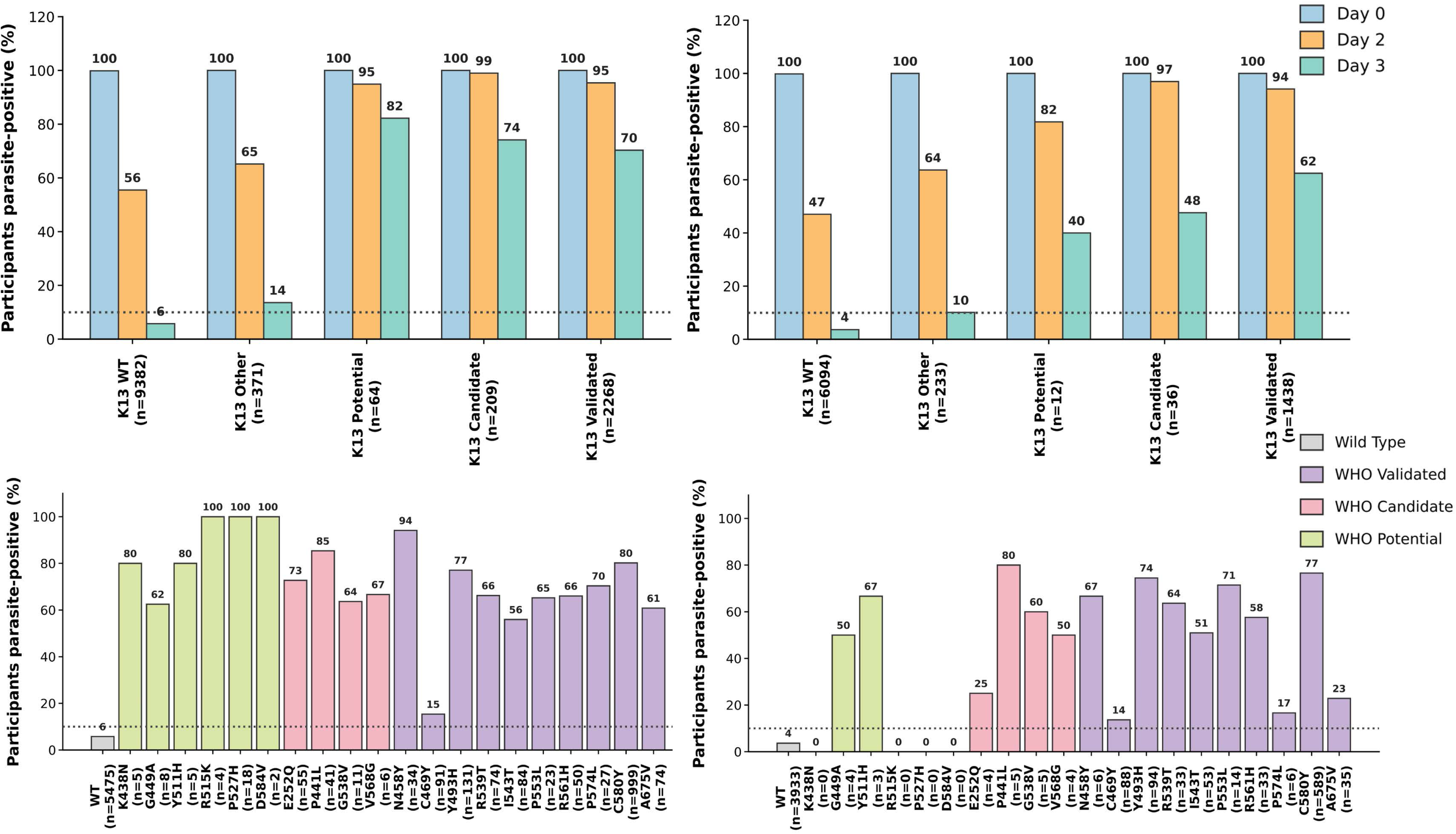
The top-left panel shows the proportion of participants remaining parasite-positive on Day 0, Day 2, and Day 3 across *Kelch13* mutation classes, including those with baseline hyperparasitaemia. The top-right shows the corresponding proportions of positivity after excluding participants with baseline hyperparasitaemia. Bottom-left and -right, show Day 3 positivity for individual *Kelch13* mutations, grouped by WHO mutation classification, including participants with baseline hyperparasitaemia, whereas bottom right shows the same after excluding baseline hyperparasitaemia. The horizontal dotted line indicates the 10% Day 3 parasite positivity threshold currently used by WHO to trigger investigations for suspected ART-R. Numbers in parentheses indicate the number of participants contributing data to each mutation class or mutation category.^1^ ^1^WT=wild-type

In adjusted analyses, WHO-listed mutations were associated with higher odds of Day 3 positivity compared with WT (WHO-validated: adjusted OR 19.8 [95% CI 16.6–23.6]; WHO-candidate: 13.7 [8.9–21.3]; WHO-potential: 27.1 [12.0–61.5]). Baseline hyperparasitaemia was also independently associated with increased odds of Day 3 positivity (OR: 2.12, 95% CI 1.8–2.5). By contrast, transmission intensity was not independently associated with Day 3 positivity after adjustment for genotype, age, baseline hyperparasitaemia, and study site (moderate-to-high vs low-to-very low transmission: adjusted OR 0.88 [95% CI 0.61–1.25]). Age showed no consistent independent association with Day 3 positivity across categories (**Supplementary file 2**).

### Parasite clearance half-life (PC1/2) and *Kelch13* mutations

Of the 7,558 patients for whom sufficient PC1/2 data were available, *Kelch13* genotype data were available for 6,172 patients across 19 studies (**Supplementary file 4; Figure S4.2**). In moderate-to-high transmission settings, the PC1/2 was 34% (95% CI: 1.25-1.44) longer among the patients carrying a WHO-validated mutation (median PC1/2 of 4.4h; IQR: 3.1h-5.7h; n=123) compared to those with WT infections (median: 2.5h, IQR: 2.0-3.3h); the effect size was adjusted for age and treatment regimen. The proportion of patients with a PC1/2>5h was 7% (138/1,992) with WT infections and 36% (44/123) with WHO-validated mutations (**Table 2**). Data on WHO-candidate and potential mutations in moderate-to-high transmission settings were limited, with a PC1/2 of 5.2h observed in one patient carrying E252Q and 2h in two patients carrying Q613E (**Table 3; Supplementary file 4**).

**Table 2:** Parasite clearance half-life by mutation status and transmission settings.

| Age | Mutation status <sup>a</sup> | Low-to-very low setting |  |  | Moderate-to-high setting |  |  |
| --- | --- | --- | --- | --- | --- | --- | --- |
|  |  | *n* | Median PC½ | PC½>5h | *n* | Median PC½ | PC½>5 |
| <1y | Wild-Type | 11 | 2.95 [2.75–3.32] | 0% (0/11) | 46 | 2.38 [1.71–3.10] | 6.5% (3/46) |
| <1y | WHO-Validated | 2 | 6.34 [5.76–6.92] | 100% (2/2) | 12 | 3.15 [2.56–4.78] | 16.7% (2/12) |
| <1y | WHO-Candidate | 1 | 7.23 | 100% (1/1) | - | - | - |
| <1y | WHO-Potential | - | - | - | - | - | - |
| <1y | Other | 1 | 3.56 | 0% (0/1) | 3 | 2.37 [2.27–2.39] | 0% (0/3) |
| 1-4y | Wild-Type | 235 | 2.87 [2.39–3.51] | 4.3% (10/235) | 724 | 2.41 [1.89–3.27] | 6.5% (47/724) |
| 1-4y | WHO-Validated | 33 | 6.32 [5.77–7.51] | 84.8% (28/33) | 56 | 4.66 [3.31–5.91] | 41.1% (23/56) |
| 1-4y | WHO-Candidate | 15 | 4.69 [4.22–5.66] | 40% (6/15) | - | - | - |
| 1-4y | WHO-Potential | 3 | 3.45 [3.11–4.38] | 33.3% (1/3) | - | - | - |
| 1-4y | Other | 10 | 4.02 [2.91–5.26] | 40% (4/10) | 41 | 1.93 [1.64–2.47] | 2.4% (1/41) |
| 5-12y | Wild-Type | 649 | 2.77 [2.22–3.55] | 4.8% (31/649) | 1164 | 2.55 [2.02–3.40] | 7.6% (88/1164) |
| 5-12y | WHO-Validated | 123 | 6.59 [5.16–7.87] | 76.4% (94/123) | 51 | 4.31 [3.06–5.29] | 33.3% (17/51) |
| 5-12y | WHO-Candidate | 42 | 4.98 [4.48–5.46] | 50% (21/42) | 0 | - | - |
| 5-12y | WHO-Potential | 15 | 4.09 [3.04–5.3] | 40% (6/15) | 2 | 1.98 [1.97–1.99] | 0% (0/2) |
| 5-12y | Other | 21 | 2.95 [2.43–4.51] | 9.5% (2/21) | 32 | 2.70 [2.06–3.87] | 3.1% (1/32) |
| >12y | Wild-Type | 1436 | 2.75 [2.13–3.43] | 5.5% (79/1436) | 34 | 2.37 [1.93–2.85] | 0% (0/34) |
| >12y | WHO-Validated | 1182 | 6·62 [5·38–7·92] | 80·7% (954/1182) | 4 | 4·90 [3·39–6·42] | 50% (2/4) |
| >12y | WHO-Candidate | 131 | 4·61 [4·08–6·16] | 44·3% (58/131) | 1 | 5·15 | 100% (1/1) |
| >12y | WHO-Potential | 28 | 5·28 [4·46–6·49] | 53·6% (15/28) | - | - | - |
| >12y | Other | 53 | 3·91 [2·52–5·44] | 30·2% (16/53) | 1 | 3·13 | 0% (0/1) |

<sup>a</sup> Patients with mutations across multiple categories are counted more than once; the percentage will add to >100%. PC<sub>1/2</sub> = Parasite clearance half-life; *n* = Number of patients; IQR = Interquartile range.

**Table 3:**
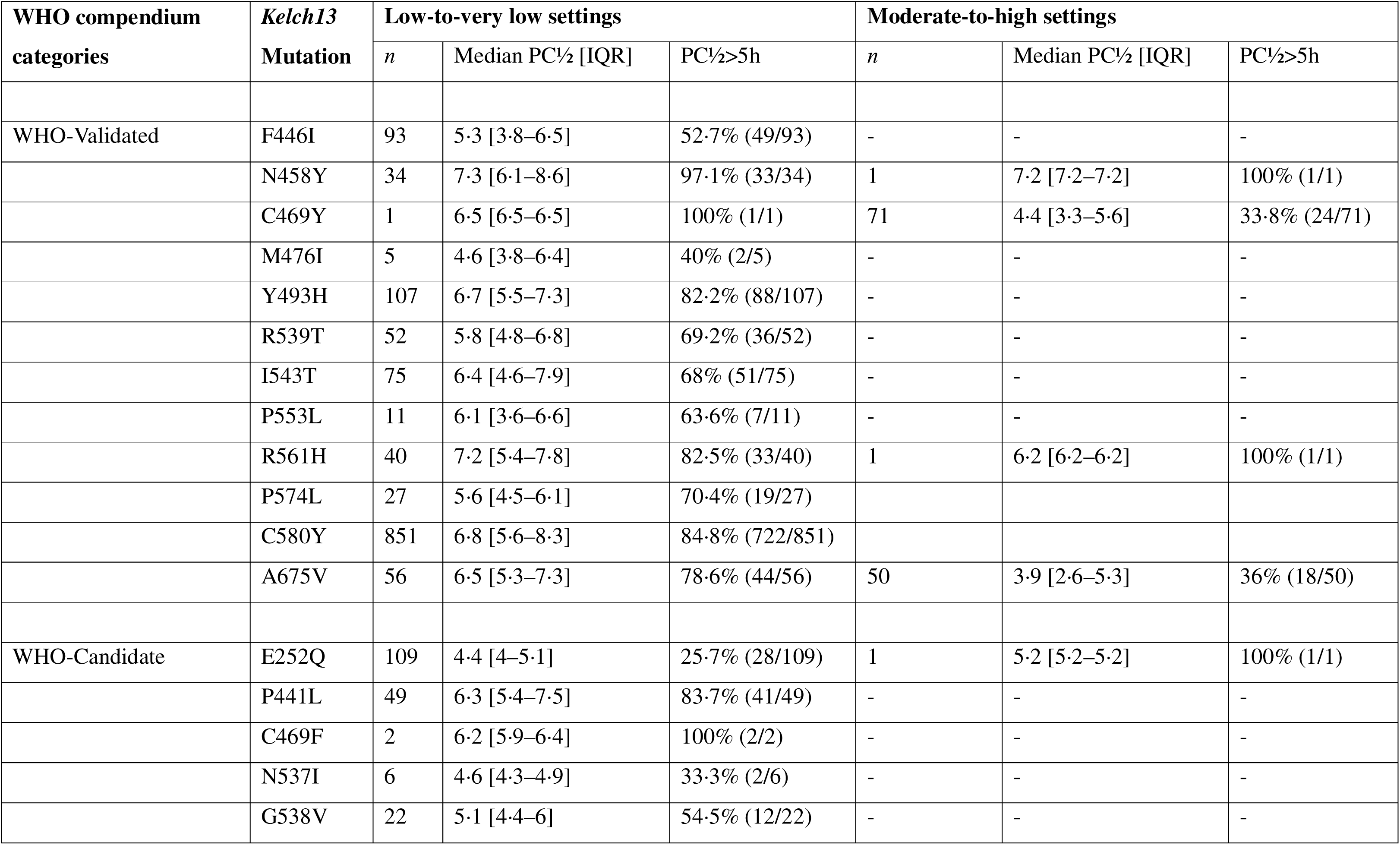
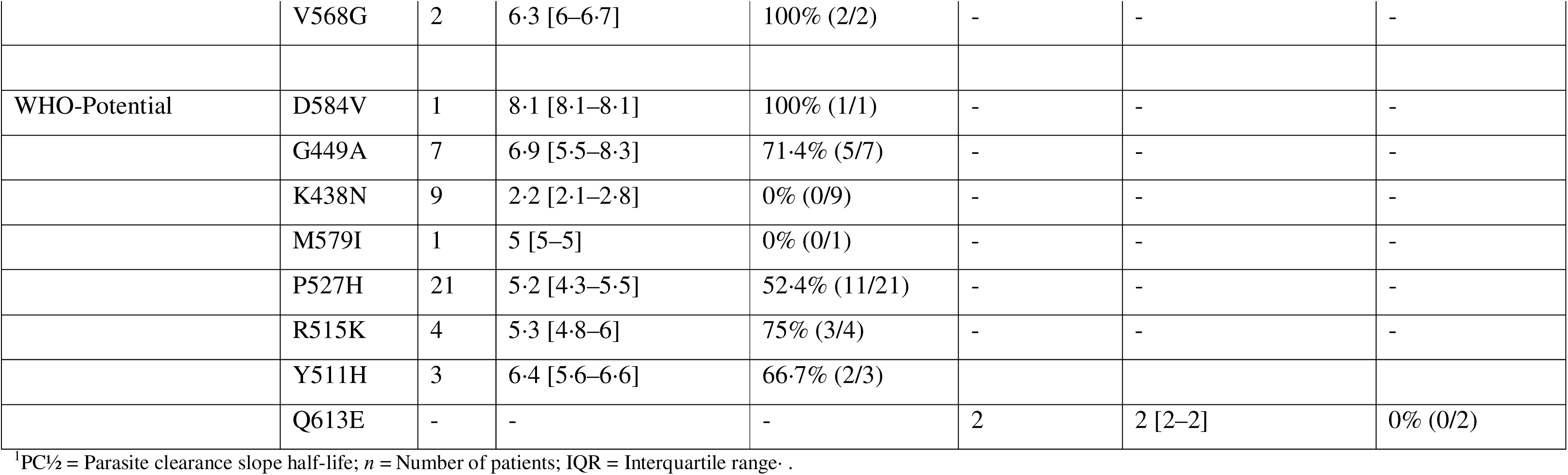
PC½ by mutation and WHO compendium classification^1^.

| WHO compendium categories | *Kelch13* Mutation | Low-to-very low settings |  |  | Moderate-to-high settings |  |  |
| --- | --- | --- | --- | --- | --- | --- | --- |
|  |  | *n* | Median PC½ [IQR] | PC½>5h | *n* | Median PC½ [IQR] | PC½>5h |
| WHO-Validated | F446I | 93 | 5·3 [3·8–6·5] | 52·7% (49/93) | - | - | - |
|  | N458Y | 34 | 7·3 [6·1–8·6] | 97·1% (33/34) | 1 | 7·2 [7·2–7·2] | 100% (1/1) |
|  | C469Y | 1 | 6·5 [6·5–6·5] | 100% (1/1) | 71 | 4·4 [3·3–5·6] | 33·8% (24/71) |
|  | M476I | 5 | 4·6 [3·8–6·4] | 40% (2/5) | - | - | - |
|  | Y493H | 107 | 6·7 [5·5–7·3] | 82·2% (88/107) | - | - | - |
|  | R539T | 52 | 5·8 [4·8–6·8] | 69·2% (36/52) | - | - | - |
|  | I543T | 75 | 6·4 [4·6–7·9] | 68% (51/75) | - | - | - |
|  | P553L | 11 | 6·1 [3·6–6·6] | 63·6% (7/11) | - | - | - |
|  | R561H | 40 | 7·2 [5·4–7·8] | 82·5% (33/40) | 1 | 6·2 [6·2–6·2] | 100% (1/1) |
|  | P574L | 27 | 5·6 [4·5–6·1] | 70·4% (19/27) |  |  |  |
|  | C580Y | 851 | 6·8 [5·6–8·3] | 84·8% (722/851) |  |  |  |
|  | A675V | 56 | 6·5 [5·3–7·3] | 78·6% (44/56) | 50 | 3·9 [2·6–5·3] | 36% (18/50) |
| WHO-Candidate | E252Q | 109 | 4·4 [4–5·1] | 25·7% (28/109) | 1 | 5·2 [5·2–5·2] | 100% (1/1) |
|  | P441L | 49 | 6·3 [5·4–7·5] | 83·7% (41/49) | - | - | - |
|  | C469F | 2 | 6·2 [5·9–6·4] | 100% (2/2) | - | - | - |
|  | N537I | 6 | 4·6 [4·3–4·9] | 33·3% (2/6) | - | - | - |
|  | G538V | 22 | 5·1 [4·4–6] | 54·5% (12/22) | - | - | - |
|  | V568G | 2 | 6·3 [6–6·7] | 100% (2/2) | - | - | - |
| WHO-Potential | D584V | 1 | 8·1 [8·1–8·1] | 100% (1/1) | - | - | - |
|  | G449A | 7 | 6·9 [5·5–8·3] | 71·4% (5/7) | - | - | - |
|  | K438N | 9 | 2·2 [2·1–2·8] | 0% (0/9) | - | - | - |
|  | M579I | 1 | 5 [5–5] | 0% (0/1) | - | - | - |
|  | P527H | 21 | 5·2 [4·3–5·5] | 52·4% (11/21) | - | - | - |
|  | R515K | 4 | 5·3 [4·8–6] | 75% (3/4) | - | - | - |
|  | Y511H | 3 | 6·4 [5·6–6·6] | 66·7% (2/3) |  |  |  |
|  | Q613E | - | - | - | 2 | 2 [2–2] | 0% (0/2) |

<sup>1</sup>PC½ = Parasite clearance slope half-life; n = Number of patients; IQR = Interquartile range .

In areas of low-to-very low transmission settings, median PC1/2 was 2.8h [IQR: 2.2-3.5, n=2,333] among patients with WT infections, 6.6h [IQR: 5.4-7.9; n=1,340] among patients with a WHO-validated mutation, 4.8h [IQR: 4.2-5.9; n=189] among patients with WHO-candidate mutations, 4.9h [IQR: 3.4-5.9; n=46] among patients with WHO-potential mutations, and 3.8h [IQR: 2.5-5.2; n=85] among those carrying other mutations (**Tables 2 and 3**). Adjusted for age and treatment regimen, patients carrying a WHO-validated mutation had a 59% increase in PC1/2 [95% CI: 54% to 64%] compared to those with WT infection. The corresponding effect sizes were 49% [95% CI: 40%-58%] for WHO-candidate mutations and 43% [95% CI: 27%-61%] for WHO-potential mutations. The proportion of patients with PC1/2>5h was 5.1% (120/2,333) in those with WT infections and 80.4% (1078/1,340) in those with a WHO-validated mutation. Increased PC1/2 among those carrying a WHO-validated mutation was observed across all age groups and transmission settings (**Table 3**).

### *Kelch13* mutant genotypes and delayed parasite clearance

The pooled analysis identified additional *Kelch13* mutant genotypes that were associated with delayed parasite clearance (**Supplementary file 4**). In areas of moderate-to-high transmission, two mutations not included in the WHO 2025 compendium with PC1/2>5h (K189T and A569S) were identified, although the number of samples with these mutations was limited to assess statistical association. The PC1/2 by the mutations are presented in **Figure 3**.

**Figure 3:**
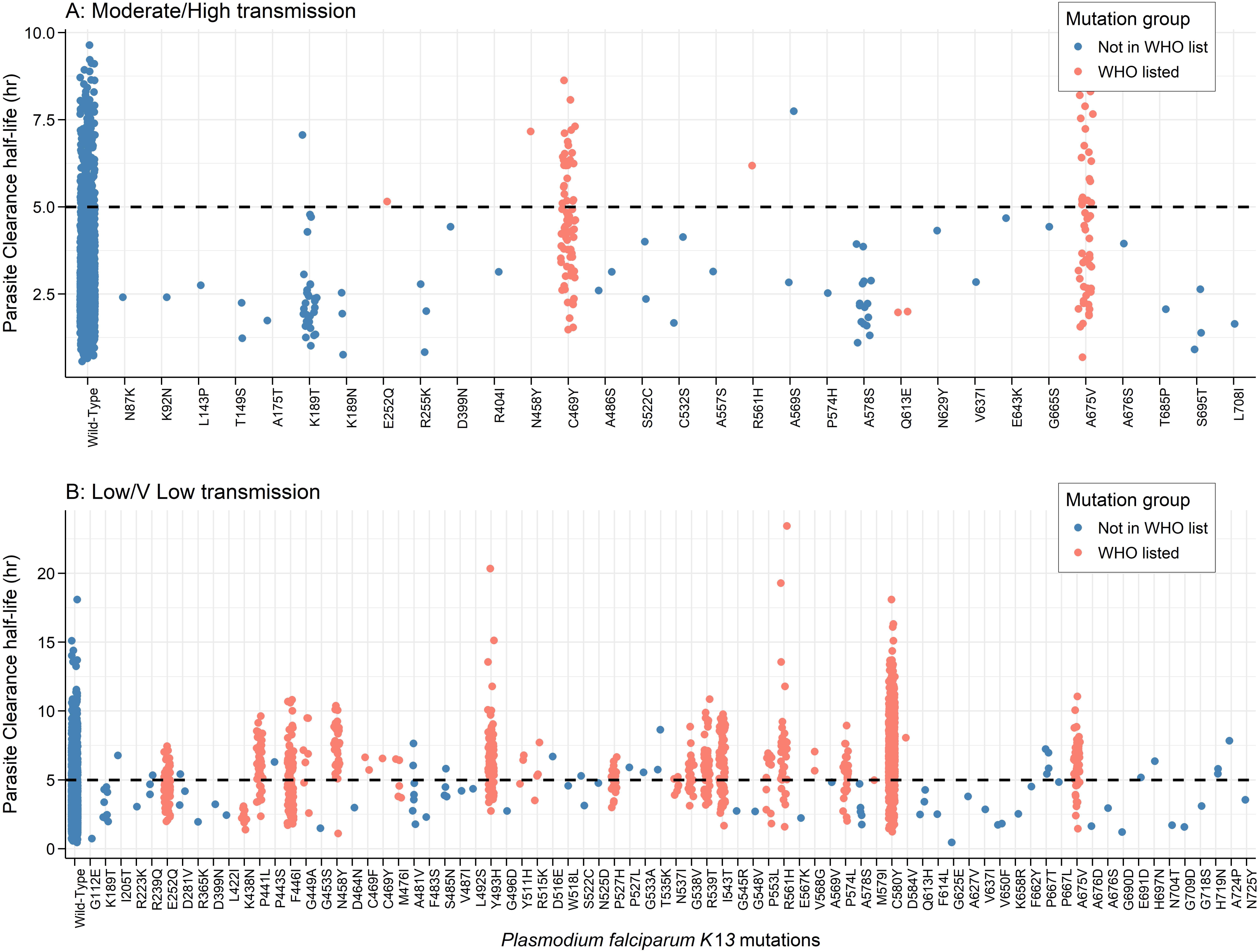
Parasite clearance half-life by mutation in the areas of (top) moderate-to-high transmission settings (bottom) low-to-very low transmission settings.

Additional *Kelch13* mutations not currently associated with ART-R in the WHO compendium, but with PC1/2>5h, included K189T, P667T, S485N, A481V, D281V, R239Q, S522C, T535K, A569S, and A724P in Africa, and G533A, P527L, D516E, A481V, P443S, and I205T in SEA.

Other WHO-listed mutations showed little evidence of delayed parasite clearance. The WHO-candidate mutations E252Q (n=109), and N537I (n=6) and the WHO-potential mutation K438N (n=9) in low-transmission settings, and the candidate mutation Q613E (n=2) in high-to moderate-to-high transmission settings all had a median PC1/2 <5h.

### Diagnostic performance of Day 2 and Day 3 positivity for identifying infections exceeding the PC1/2 >5 h benchmark

Day 2 positivity was highly sensitive but poorly specific for PC1/2, whereas Day 3 positivity provided improved specificity but lower sensitivity. Day 3 positivity demonstrated moderate sensitivity (75% [1119/1487]; 95% CI 73.0–77.0) and high specificity (96% [2814/2939]; 95% CI 95.0–96.0) for detecting delayed parasite clearance (PC1/2>5h), although performance varied across epidemiological contexts (**Supplementary file 3**). Sensitivity was higher in Asia (82%, 1042/1277) than in Africa (37%, 77/210), despite higher specificity in Africa (99% vs 92%). Similarly, sensitivity was lower in moderate-to-high transmission settings (27.6%, 43/156; 95% CI 20.7–35.8) than in low-to-very low transmission settings (81%, 1076/1331; 95% CI 78.6–82.8).

The predicted probability of Day 3 positivity increased with increasing PC1/2. However, the relationship between Day 3 positivity and parasite clearance depends on transmission intensity. A predicted probability of 10% Day 3 positivity corresponded to a PC1/2 of approximately 3.5h in very low-to-low transmission settings, but approximately 5h in moderate-to-high transmission settings. Conversely, at a PC1/2 of 5h, the predicted probability of Day 3 positivity was approximately 10% in moderate-to-high transmission settings compared with 38% in low-to-very-low transmission settings. (**Supplementary file 3**).

### PC1/2 thresholds distinguishing wild-type and *Kelch13* mutations across transmission settings

PC1/2>5h had a sensitivity of 81% (78.5 - 82.8) and a specificity of 95% (93.9 - 95.7) in low- to very-low settings for identifying WHO-validated mutations. The corresponding estimates in the moderate-to-high transmission settings were 36% (27.9 - 44.6) and 93.1% (91.9 - 94.1), indicating a poor sensitivity for the currently adopted threshold of 5h (**Table 4**). With equal weight assigned to sensitivity and specificity, the PC1/2 threshold that best separated the distributions of WT and WHO-validated mutations in moderate-to-high transmission settings was 3.1h (AUROC=0.78, 95% CI: 0.74-0.82, sensitivity:75%, 95% CI: 67%-82%, specificity: 71%, 95% CI: 69%-73%) (**Figure 4**). When sensitivity was weighted at 70%, the threshold was reduced to 2.6h (sensitivity: 85%, specificity: 56%). The corresponding threshold (assuming equal weight to sensitivity and specificity) in the lower-transmission settings was 4.2h (AUROC: 0.94, 95% CI: 0.93-0.95), with sensitivity and specificity of 89% (95% CI: 87%-91%) and 90% (95% CI: 88%-91%), respectively (**Figure 4**).

**Figure 4:**
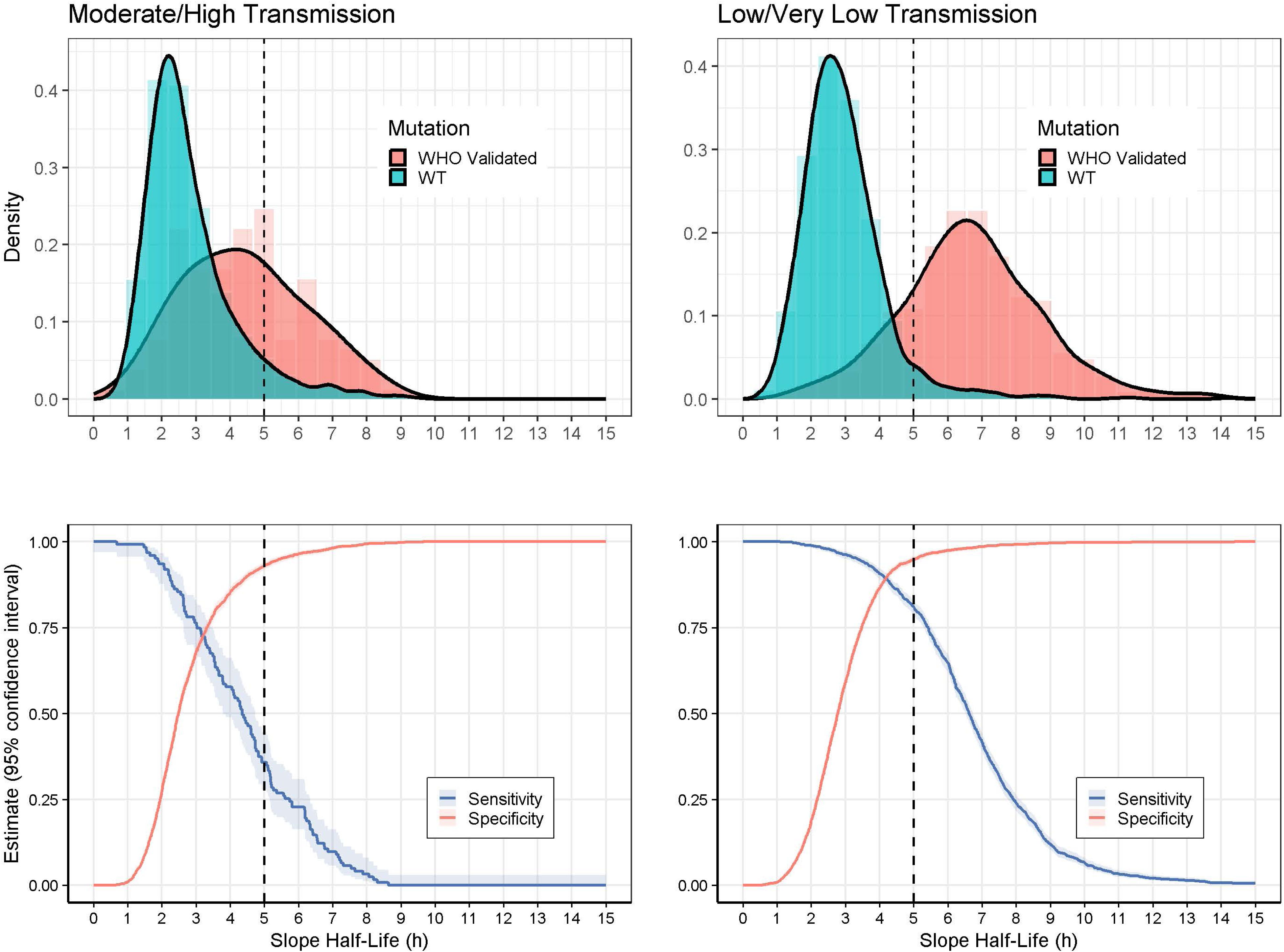
Parasite clearance half-life (PC1/2) distribution for WHO-validated mutations and wild-type, along with sensitivity and specificity of different PC1/2 values in identifying WHO-validated mutations **Legend:** Parasite clearance half-life distribution (top panels) and the diagnostic accuracy of different half-life thresholds (bottom panels). The left panels present data for moderate-to-high transmission settings, and the right panels present data for low and very low transmission settings. For the bottom panels, the shaded interval indicates the 95% confidence interval estimated from all data for the respective transmission settings and ignores clustering. The vertical dotted line indicates the current 5h threshold.

**Table 4:** PC½ thresholds for phenotypic resistance (WHO-validated mutation vs Wild-Type)^1^.

|  | Low-to-very low transmission |  |  |  | Moderate-to-high transmission |  |  |  |
| --- | --- | --- | --- | --- | --- | --- | --- | --- |
| PC½ (hours) | Sensitivity [95% CI] | Specificity [95% CI] | PPV | NPV | Sensitivity [95% CI] | Specificity [95% CI] | PPV | NPV |
| >3 | 96·2% (95·1 - 97·1) | 59·1% (57·1 - 61·1) | 12·8% | 97·9% | 76·4% (68·2 - 83·1) | 68% (65·9 - 70) | 57·2% | 96·5% |
| >3·5 | 94·2% (92·8 - 95·3) | 76% (74·2 - 77·6) | 16·0% | 97·4% | 66·7% (57·9 - 74·4) | 78·4% (76·6 - 80·2) | 69·0% | 95·8% |
| >4 | 90·7% (89 - 92·2) | 86·5% (85·1 - 87·8) | 19·5% | 97·0% | 57·7% (48·9 - 66·1) | 85·3% (83·7 - 86·8) | 79·2% | 94·3% |
| >4·5 | 86·2% (84·2 - 87·9) | 92·5% (91·4 - 93·5) | 22·5% | 96·5% | 47·2% (38·6 - 55·9) | 90% (88·6 - 91·2) | 86·7% | 92·2% |
| >5 | 80·8% (78·5 - 82·8) | 94·9% (93·9 - 95·7) | 24·2% | 95·9% | 35·8% (27·9 - 44·6) | 93·1% (91·9 - 94·1) | 89·9% | 89·7% |
| >5·5 | 72·9% (70·4 - 75·2) | 96·7% (95·8 - 97·3) | 25·4% | 95·5% | 26·8% (19·8 - 35·3) | 95·1% (94·1 - 96) | 92·5% | 86·3% |

<sup>1</sup>PC½ = Parasite clearance slope half-life; PPV: Positive Predictive Value; NPV: Negative Predictive Value; 95% CI: 95% confidence interval; 95% CI estimated using all data for the respective transmission settings and ignores clustering. WHO-validated mutations were identified based on the 2025 revised compendium.

## Discussion

This IPDMA provides the largest available global assessment of the molecular and epidemiological determinants of phenotypic expression of ART-R *Kelch13* mutations. By integrating harmonised data across Africa and SEA, our findings demonstrate that ART-R expression and detection were strongly shaped by transmission intensity, aspects of host immunity, and the complexity of infection, rather than by parasite genotype alone. Nevertheless, certain *Kelch13* mutations remained consistently associated with delayed parasite clearance across all settings..

The widely used ART-R threshold of PC1/2>5h had a low sensitivity in moderate-to-high transmission settings, suggesting that this benchmark may not be appropriate across all epidemiological contexts. These findings support the need for transmission intensity-specific interpretation of phenotypic markers in ART-R surveillance. This IPDMA suggested that in low-to-very-low transmission settings, a PC1/2 threshold of 4h would be more sensitive than 5h (91% vs 81%) while retaining high specificity (86%). In areas of moderate-to-high transmission, a positivity threshold of 3h will improve sensitivity from 36% under the current 5h threshold to 76%, while reducing specificity to 68%. The Positive Predictive Values (PPVs) remained low regardless of the PC1/2 threshold, whereas the Negative Predictive Values (NPVs) were consistently high (>95%), indicating high accuracy of PC1/2 values in ruling out mutations, but less reliability as a “rule-in” test. In contrast, in areas of low-to-very low transmission, both PPVs and NPVs were generally high for PC1/2 values >4h.

Compared to moderate-to-high transmission settings, delayed parasite clearance was more pronounced and more readily detectable in low- and very low-transmission settings, where reduced acquired immunity means parasite clearance depends largely on the intrinsic activity of the antimalarial drug, allowing ART-R phenotypes to be more clearly expressed (8, 12, 23–25). In contrast, in moderate- to high-transmission settings, partial immunity and multiclonal infections likely accelerate parasite clearance and obscure ART-R phenotypes, particularly where resistant parasites incur a fitness cost (8, 14, 23). These patterns are consistent with the longer history of ART-R in SEA, which likely contributes to the higher burden of slow-clearing infections observed (8, 14, 23).

Importantly, despite faster overall clearance in Africa, delayed clearance remained associated with WHO-validated mutations (C469Y, R561H, P441L, and A675V), indicating that ART-R is becoming established across diverse epidemiological contexts on the continent (8, 14, 23)., Ethiopia now being the fifth country to meet WHO ART-R confirmation criteria (30). Our findings suggest that both molecular markers and phenotypic thresholds must be interpreted within their epidemiological context, and that integrating molecular surveillance with clinical phenotyping and refining transmission intensity-specific thresholds can improve ART-R characterisation.

This study expands the current understanding of the molecular landscape of ART-R by identifying potentially novel *Kelch13* genotypes associated with delayed parasite clearance, the majority in the propeller gene region. These occurred at low frequencies and require broader surveillance (beyond WHO-validated and -candidate markers) to determine their ART-R association. Notably, the T535K mutation, recently detected in samples in Kigali, Rwanda, in 2023, was associated with markedly prolonged clearance (three studied samples), a pattern reminiscent of that observed in the first reports on ART-R mediated through R561H in Rwanda (26), and C469Y in Uganda (27).

In agreement with recent WHO findings (28), and those of others (29), WHO-validated *Kelch13* mutations, including C469Y, R561H, and A675V, were associated with delayed parasite clearance and persistent Day 3 positivity. Several WHO-candidate and potential mutations, including P441L, G538V, G449A, Y511H, and P527H, were also associated with prolonged parasite clearance, although sample sizes were smaller. However, the identification of emerging genotypes in African parasite populations provides further evidence that ART-R in Africa is not solely driven by mutations originally described in SEA (12) and offers recent insights beyond East African data (29). These findings suggest that ART-R in Africa is evolving along distinct trajectories. As such, assaying only for known mutations may underestimate the diversity and spread of ART-R, particularly in Africa.

After accounting for baseline hyperparasitaemia, the simple measure of Day 3 positivity retained high specificity for delayed parasite clearance measured by PC1/2. However, its sensitivity was lower in moderate-to-high transmission settings, indicating that its absence should not be interpreted as evidence against emerging ART-R. Importantly, much of the evidence supporting current surveillance frameworks predates the emergence and spread of ART-R in Africa. Continued evaluation of Day 3 positivity alongside PC1/2 and molecular surveillance will therefore be essential to refine surveillance strategies as ART-R continues to emerge and spread across Africa.

Several limitations should be considered. This analysis did not evaluate partner-drug resistance. Heterogeneity in study design, sampling frequency, genotyping approaches, and reporting practices may have influenced estimates despite data harmonisation; however, profiles with poor fit of the parasite clearance estimator were excluded from the primary analyses (**Supplementary File 5**). Many studies from sub-Saharan Africa predated the emergence of ART-R, and surveillance often prioritised known mutations, potentially underrepresenting novel or rare variants. Recent data were also limited from parts of central and southern Africa. Finally, IPD were unavailable for 47% of the requested studies; however, a systematic assessment of these studies indicated that they predominantly reported mutations already present in the IPD-MA (**Supplementary File 5**).

By integrating harmonised molecular and clinical IPD, this study provides important insights into the evolving epidemiology and phenotypic expression of ART-R across diverse transmission settings. The findings strengthen the evidence base supporting WHO-validated, -candidate, and -potential mutations as markers of ART-R and identifying additional emerging variants associated with delayed parasite clearance in Africa and SEA parasite populations. Together, these results support the continued refinement of the WHO-listed mutations, the development of region-specific phenotypic thresholds for parasite clearance, and the strengthening of molecular surveillance frameworks to guide evidence-based malaria treatment and control strategies.

## Declarations

We declare no competing interests.

## Generative AI declaration

Generative AI-assisted tools were used to support language editing and improve readability. All scientific content was reviewed and approved by the authors.

## Data availability

This study analysed data from the Infectious Diseases Data Observatory (IDDO) data inventory, and therefore the datasets are owned by the original controllers, who are responsible for ensuring that data were collected in accordance with applicable laws and ethical approval in the countries where the study was conducted. Access requests can be submitted via the IDDO platform (https://www.iddo.org/data-sharing/accessing-data). More information about IDDO’s data governance can be found on IDDO’s website (https://iddo.org)

## Acknowledgements

We acknowledge the contributions of the WorldWide Antimalarial Resistance Network Correlation between *Kelch13* mutations and clinical phenotype Study Group (https://www.iddo.org/correlation-between-Kelch13-mutations-and-clinical-phenotype-study-group-update). We thank the IDDO data curation and harmonisation team for their efforts in collating, standardising, and preparing the individual participant data used in this analysis. We also thank the investigators, study teams, laboratory personnel, and participants across malaria-endemic regions whose therapeutic efficacy studies and laboratory work generated the primary data synthesised in this review. We are grateful to colleagues from the World Health Organization Global Malaria Programme for constructive discussions. The findings and conclusions in this paper are those of the authors and do not necessarily represent the views of the US Centers for Disease Control and Prevention.

## Ethics declaration

In addition to the de-identification processes completed by data contributors, IDDO performs additional checks before or during data curation to ensure that data are pseudonymised. This process is conducted before releasing it to data requestors to ensure full compliance with international regulations, including the UK Data Protection Regulation (UK GDPR), the European Union Data Protection Regulation (EU GDPR), and the Data Protection Act 2018. More information about IDDO’s data governance is available on IDDO’s website (https://iddo.org).

In accordance with the guidelines of the University of Oxford Central University Research Ethics Committee, secondary analyses of pseudonymised data do not require additional ethics review.

## Contributors

SvW, PD, KIB, PR, JW, PJG, and MD comprised the core writing and scientific interpretation group. SvW led the study, including conceptualisation, methodology, project administration, data curation, formal analysis, software, validation, investigation, visualisation, and preparation of the original manuscript. PD contributed to conceptualisation, methodology, project administration, supervision, formal analysis, software, validation, investigation, visualisation, funding acquisition and review and editing of the manuscript. KB contributed to conceptualisation, methodology, supervision, project administration, funding acquisition, and review and editing of the manuscript. PJG contributed to conceptualisation, methodology, supervision, project administration, funding acquisition, and writing and critical revision of the manuscript. PR contributed to the interpretation of the findings, the critical review, and the editing of the manuscript. MD contributed to the investigation, data curation, provision of study resources, interpretation of the findings, and review and editing of the manuscript.

Members of the WWARN Kelch13 Genotype-Phenotype Study Group contributed individual participant data and associated study resources, data curation and interpretation, methodological or analytical expertise where applicable, and critical review and editing of the manuscript. The Study Group includes investigators responsible for the design, conduct, data generation, and scientific oversight of the contributing studies, as well as collaborators involved in data harmonisation, curation, management, and analysis.

SvW and PD directly accessed and verified the underlying individual participant data reported in the manuscript.

All authors critically reviewed the manuscript, approved the final version for submission, and accept responsibility for the work.

## References

1. World Health Organization The World Malaria Report 2025.

2. World Health Organization Antimalarial drug combination therapy. Report of a WHO technical consultation. 2001.

3. Li J, Docile HJ, Fisher D, Pronyuk K, Zhao L. Current status of malaria control and elimination in Africa: epidemiology, diagnosis, treatment, progress and challenges. Journal of Epidemiology and Global Health. 2024;14(3):561–79.

4. Boni MF, Soulama I, Opigo J, Watson OJ, Ogutu B. Slowing artemisinin resistance in Africa. Science Advances. 2025;11(43):eaeb7618.

5. World Health Organization. Global malaria programme operational strategy 2024-2030: World Health Organization; 2024.

6. Noedl H, Se Y, Schaecher K, Smith BL, Socheat D, Fukuda MM. Evidence of artemisinin-resistant malaria in western Cambodia. New England Journal of Medicine. 2008;359(24):2619–20.

7. Africa MS. Antimalarial Drug Resistance Profile for Southern African Development Community Countries. 2025.

8. Rosenthal PJ, Asua V, Conrad MD. Emergence, transmission dynamics and mechanisms of artemisinin partial resistance in malaria parasites in Africa. Nature Reviews Microbiology. 2024;22(6):373–84.

9. Eloff L, Aranda-Díaz A, Routledge I, Wesolowski A, Chisenga M, Mangena B, et al. High prevalence of molecular markers associated with artemisinin, sulphadoxine and pyrimethamine resistance in northern Namibia. medRxiv. 2025:2025.01. 09.25320247.

10. Fola AA, Kobayashi T, Hamapumbu H, Musonda M, Katowa B, Matoba J, et al. Temporal genomics in Southern Zambia shows rising prevalence of *Plasmodium falciparum* mutations linked to delayed clearance after artemisinin-lumefantrine treatment. Scientific reports. 2024;14(1):26789.

11. Martin AC, Sadler JM, Simkin A, Musonda M, Katowa B, Matoba J, et al. Emergence and rising prevalence of artemisinin partial resistance marker *kelch13* P441L in a low malaria transmission setting in Southern Zambia. The Journal of Infectious Diseases. 2025;232(4):918–22.

12. Association of mutations in the *Plasmodium falciparum Kelch13* gene (Pf3D7_1343700) with parasite clearance rates after artemisinin-based treatments—a WWARN individual patient data meta-analysis. BMC Medicine. 2019;17(1):1.

13. Rosenthal PJ, Asua V, Bailey JA, Conrad MD, Ishengoma DS, Kamya MR, et al. The emergence of artemisinin partial resistance in Africa: how do we respond? The Lancet Infectious Diseases. 2024;24(9):e591–e600.

14. Dondorp AM, Nosten F, Yi P, Das D, Phyo AP, Tarning J, et al. Artemisinin resistance in *Plasmodium falciparum* malaria. New England journal of medicine. 2009;361(5):455–67.

15. Angwe MK, Mwebaza N, Nsobya SL, Vudriko P, Dralabu S, Omali D, et al. Day 3 parasitemia and *Plasmodium falciparum Kelch 13* mutations among uncomplicated malaria patients treated with artemether-lumefantrine in Adjumani district, Uganda. Plos one. 2024;19(6):e0305064.

16. Ariey F, Witkowski B, Amaratunga C, Beghain J, Langlois A-C, Khim N, et al. A molecular marker of artemisinin-resistant *Plasmodium falciparum* malaria. Nature. 2014;505(7481):50–5.

17. Ashley EA, Dhorda M, Fairhurst RM, Amaratunga C, Lim P, Suon S, et al. Spread of artemisinin resistance in *Plasmodium falciparum* malaria. New England Journal of Medicine. 2014;371(5):411–23.

18. World Health Organization. Report on antimalarial drug efficacy, resistance and response: 10 years of surveillance (2010-2019): World Health Organization; 2020.

19. World Health Organization. Minutes of the Expert Review Committee on K13 molecular marker of artemisinin resistance. 2014.

20. Flegg JA, Guerin PJ, White NJ, Stepniewska K. Standardizing the measurement of parasite clearance in falciparum malaria: the parasite clearance estimator. Malaria Journal. 2011;10(1):339.

21. van Wyk S, Dahal P, Vouvoungui C, Ayuen DS, Shokraneh F, Soma A, et al. Investigating the relationship between Pfkelch13 mutations and response to artemisinin-based treatment for uncomplicated falciparum malaria: a protocol for a systematic review and individual patient data meta-analysis. BMJ Open. 2025;15(7):e100251.

22. Guerra CA, Hay SI, Lucioparedes LS, Gikandi PW, Tatem AJ, Noor AM, et al. Assembling a global database of malaria parasite prevalence for the Malaria Atlas Project. Malaria Journal. 2007;6(1):17.

23. Ataide R, Ashley EA, Powell R, Chan J-A, Malloy MJ, O’Flaherty K, et al. Host immunity to *Plasmodium falciparum* and the assessment of emerging artemisinin resistance in a multinational cohort. Proceedings of the National Academy of Sciences. 2017;114(13):3515–20.

24. Ataíde R, Powell R, Moore K, McLean A, Phyo AP, Nair S, et al. Declining transmission and immunity to malaria and emerging artemisinin resistance in Thailand: a longitudinal study. The Journal of Infectious Diseases. 2017;216(6):723–31.

25. Barati S, Haghi AM, Nateghpour M, Zamani Z, Khodaveisi S, Etemadi S. Induction of Artesunate Resistance in *Plasmodium falciparum* 3D7 Strain Using Intermittent Exposure Method and Comparing *Pfk13* Sequence between Susceptible and Resistant Strains. Iran J Parasitol. 2023;18(4):445–55.

26. Uwimana A, Legrand E, Stokes BH, Ndikumana J-LM, Warsame M, Umulisa N, et al. Emergence and clonal expansion of in vitro artemisinin-resistant *Plasmodium falciparum kelch13* R561H mutant parasites in Rwanda. Nature medicine. 2020;26(10):1602–8.

27. Awor P, Copositivityée R, Khim N, Rondepierre L, Roesch C, Khean C, et al. Indigenous emergence and spread of kelch13 C469Y artemisinin-resistant *Plasmodium falciparum* in Uganda. Antimicrobial Agents and Chemotherapy. 2024;68(8):e01659–23.

28. World Health Organization. Compendium of molecular markers for antimalarial drug resistance. 2025.

29. Hosangadi V, Fidock DA. Artemisinin partial resistance at a crossroads: evidence for continental variation in *Plasmodium falciparum* clearance phenotypes. The Lancet Infectious Diseases. 2026.

30. Chala B, Platon L, Tibiri YN, Strubel PE, Thiebaut L, Wade A, Caspar E, Muller A, Bastien M, Nigussie H, Zhang Q. Clinical, molecular, and in vitro evidence of artemisinin partial resistance in Ethiopian *Plasmodium falciparum*: a prospective, multisite, surveillance study. The Lancet Infectious Diseases. 2026.

