## Appendix for "Association between *Plasmodium falciparum Kelch13* mutations and malaria parasite clearance half-life after artemisinin-based therapy: an updated WWARN systematic review and individual patient data meta-analysis": van_Wyk_2026_Kelch13_Supplementary_Materials.pdf

**Supplementary Materials**
**Table of Contents**

- WWARN *Kelch13* Genotype-Phenotype Study Group Membership Authors and Affiliations pages I- VIII

**Supplementary File 1**

| Contents | Page |
|---|---|
| Figure S1: PRISMA flow diagram of study selection for the IPD meta-analysis | 2 |
| Figure S2: Study profile and data availability for parasite clearance analyses following treatment with artemisinin-based therapies | 3 |
| Table S1: Summary of studies from which the pooled data were included in this analysis | 4 |
| Table S2: Genotyping results of included studies | 11 |
| Table S3: Demographic summary of participants included in the pooled analysis | 19 |
| Supplementary references | 21 |

**Supplementary File 2**

| Contents | Page |
|---|---|
| Table S4: Distribution of included studies and patients for Day 2 and Day 3 parasite positivity by region and transmission settings, with linked clinical and Kelch13 genotype data | 2 |
| Supplementary Text S1: Day 2 and Day 3 Parasite Positivity | 2 |
| Table S5: Association between Kelch13 mutation classification and Day 3 parasitaemia in patients with uncomplicated falciparum malaria | 4 |
| Supplementary Text S2: Association between geographic region, resistance status, transmission setting and Day 3 parasitaemia in patients with falciparum malaria | 5 |
| Table S6: Association between geographic region and Day 3 parasitaemia in patients with falciparum malaria | 5 |
| Figure S4: Study-specific crude associations between Plasmodium falciparum Kelch13 mutation status and Day 3 parasite positivity | 6 |
| Table S7: Cross-reference between forest plot labels, study characteristics, and corresponding publication | 7 |
| Figure S5: Study-specific crude associations between Kelch13 status and parasite clearance half-life | 9 |
| Figure S6: Day 3 parasitaemia by country resistance status | 10 |
| Supplementary Text S3: Exploratory effect-modification analysis by transmission intensity | 10 |
| Supplementary References 2 | 11 |

**Supplementary File 3**

| Contents | Page |
|---|---|
| Supplementary Text S4: Correspondence between PC½ >5 h and Day 3 parasitaemia | 2 |
| Table S8: Diagnostic performance of Day 3 parasitaemia for detecting delayed parasite clearance (PC½ >5 h) | 2 |
| Supplementary Figure S7: Predicted probability of Day 3 parasitaemia by parasite clearance half-life (PC½), overall and stratified by transmission intensity | 3 |
| Supplementary Text S5: Correspondence between Day 3 positivity and PC½ >5 h | 3 |
| Table S9: Diagnostic performance of Day 2 parasitaemia | 4 |

**Supplementary File 4: Additional results summarising parasite clearance slope half-life (PC½)**

| Contents | Page |
|---|---|
| S Figure 4.1: Distribution of slope half-life by country and calendar period | 3 |
| S Figure 4.2: Distribution of slope half-life by transmission, continent, and age group | 3 |
| S Figure 4.3: Distribution of slope half-life for WHO-validated mutations, by country | 4 |
| S Figure 4.4: Distribution of slope half-life for WHO candidate mutations, by country | 5 |
| S Figure 4.5: Distribution of slope half-life for WHO potential mutations, by country | 6 |
| S Figure 4.6: Distribution of slope half-life for mutations not current in the 2025 WHO compendium of markers, by country | 7 |
| S Figure 4.7: Slope half-life distribution by evolution of individual mutation over time period, Asia | 8 |
| S Figure 4.8: Slope half-life distribution by evolution of individual mutation over time period, Africa | 9 |
| S Figure 4.9: Slope half-life distribution by mutation category, by transmission groups | 10 |
| S Figure 4.10: Slope half-life distribution by mutation category, by continent | 11 |
| S Figure 4.11: ROC curves for diagnostic accuracy of slope half-life thresholds for identifying a WHO-validated mutation versus WT, by transmission | 12 |
| S Figure 4.12: Slope half-life by mutation categories in areas of moderate/high transmission | 13 |
| S Figure 4.13: Slope half-life by mutation categories in areas of moderate/high transmission, for selected mutations | 14 |
| S Table 4.1: Parasite clearance slope half-life by regions | 15 |

S Table 4.1B: Parasite clearance slope half-life by transmission settings 16
S Table 4.2: Distribution of slope half-life (PC½) by country 17
S Table 4.3: Distribution of slope half-life (PC½) by country, above the given half-life threshold 18
S Table 4.4: Additional mutations observed in areas of moderate/high transmission that were not listed in the WHO 2025 compendium 19
S Table 4.5: Additional mutations observed in areas of low/very low transmission that were not listed in the WHO 2025 compendium 20
S Table 4.6: Diagnostic accuracy of different slope half-life thresholds in identifying WHO-validated mutations, by transmission settings 22

**Supplementary File 5: Risk of availability bias due to unavailable IPD in studies with parasite clearance slope half-life (PC½) measured**

| Contents | Page |
|---|---|
| Text S5.1: Risk of bias assessment | 2 |
| Table S5.1: Risk of bias in the studies based on profile exclusion | 2 |
| S Table 5.2: Assessment of bias due to studies not available for IPD-MA from Asia (n=24) | 3 |
| S Table 5.3: Assessment of bias due to studies not available for IPD-MA from Africa (n=15) | 9 |
| S Table 5.4: Assessment of bias due to studies not available for IPD-MA from South America (n=1) | 12 |

WWARN *Kelch13* Genotype-Phenotype Study Group membership Authors and Affiliations

**Authors** Stephanie van Wyk; WWARNE K13 Genotype-Phenotype Study Group; Philip J Rosenthal; Philippe J Guerin; Mehul Dhorda; Karen Barnes; Prabin Dahal.

Lead author

**Stephanie van Wyk** — Mitigating Antimalarial Resistance Consortium for Southern and Eastern Africa (MARC SE-Africa), Cape Town, South Africa; Collaborating Center for Optimizing Antimalarial Therapy (CCOAT), Division of Clinical Pharmacology, Department of Medicine, University of Cape Town, Cape Town, South Africa; WorldWide Antimalarial Resistance Network (WWARN), Oxford, UK; Infectious Diseases Data Observatory (IDDO), Oxford, UK

**WWARN *Kelch13* Genotype-Phenotype Study Group membership:**

**Mohamed Abdelrahim** — Institute for Immunology and Infection Research, School of Biological Sciences, The University of Edinburgh, UK and Institute of Nuclear Applications in Biological Sciences (NABSI), Sudan Atomic Energy Commission (SAEC), Khartoum, Sudan, Institute of Endemic Diseases, University of Khartoum, Khartoum, Sudan

**Desmond Omane Acheampong** — Department of Biomedical Sciences, School of Allied Health Sciences, University of Cape Coast, Cape Coast

**George Obeng Adjei** — Centre for Tropical Clinical Pharmacology and Therapeutics, University of Ghana Medical School, College of Health Sciences, University of Ghana

**Samuel Yao Ahorhorlu** — Centre for Tropical Clinical Pharmacology and Therapeutics, University of Ghana Medical School, College of Health Sciences, University of Ghana, P.O. Box 4236, Accra, Ghana.

**Mohammad Shafiul Alam** — International Centre for Diarrhoeal Disease Research, Bangladesh (icddr,b)

**Chanaki Amaratunga** — Mahidol-Oxford Tropical Medicine Research Unit, Faculty of Tropical Medicine, Mahidol University, Bangkok, Thailand Centre for Tropical Medicine and Global Health, Nuffield Department of Medicine, University of Oxford, Oxford, United Kingdom

**Martin Kamilo Angwe** — 1. School of Biosecurity, Biotechnical and Laboratory Science, College of Veterinary Medicine, Animal Resources and Biosecurity, Makerere University, Kampala, Uganda. 2. Department of Pharmacology and Therapeutics, Institute of Systems, Molecular and Integrative Biology, University of Liverpool, Liverpool, UK

**Enoch Aninagyei** — Department of Biomedical Sciences, School of Basic and Biomedical Sciences, University of Health and Allied Sciences, PMB 31, Ho, Volta region, Ghana

**Elizabeth A Ashley** — 1Lao-Oxford-Mahosot Hospital-Wellcome Trust Research Unit (LOMWRU), Mahosot Hospital, Vientiane, Lao PDR. 2 Centre for Tropical Medicine, Nuffield Department of Clinical Medicine, University of Oxford, Oxford, United Kingdom

**Ashenafi Assefa** — Malaria and Neglected Tropical Diseases Research Directorate, Ethiopian Public Health Institute, Addis Ababa, Ethiopia, Institute of Global Health, University of North Carolina at Chapel Hill, Chapel Hill, USA

**Essoham Ataba** — National Malaria Control Program, Ministry of Health, Public Hygiene, Universal Health Coverage, and Insurance, Lomé, Togo, Higher School of Biological and Food Technology, University of Lomé, Lomé, Togo

**Mwaka Kakolwa Athuman** — Shinda Malaria program at the Ifakara Health Institute, Tanzania

**Dhol Samuel Ayuen** — Worldwide Antimalarial Resistance Network (WWARN), Oxford, United Kingdom, Infectious Diseases Data Observatory (IDDO), Oxford, United Kingdom, Centre for Tropical Medicine and Global Health, University of Oxford, Oxford, United Kingdom

**Abdoul Habib Beavogui** — Centre National de Formation et de Recherche en Santé Rurale de Mafèrinyah, Forécariah, Guinea

**Balikagala Betty** — Tropical Medicine and Parasitology at Juntendo University in Tokyo, Japan

**Emmanuel Bottieau** — Department of Clinical Sciences, Institute of Tropical Medicine, Antwerp, Belgium

**Zbynek Bozdech** — School of Biological Sciences, Nanyang Technological University,

**Arlindo Chidimatembue** — Centro de Investigação em Saúde de Manhiça, Maputo, Mozambique

**Umberto D'Alessandro** — London School of Hygiene and Tropical Medicine, Gambia

**Sabyasachi Das** — Dept. of Physiology, Faculty of Medicine, Universiti Malaya

**Nicholas P J Day** — 1. Mahidol Oxford Tropical Medicine Research Unit, Faculty of Tropical Medicine, Mahidol University, Bangkok, Thailand. 2. Centre for Tropical Medicine and Global Health, Nuffield Department of Medicine, University of Oxford, Oxford, United Kingdom.

**Das Debashish** — Infectious Diseases Data Observatory (IDDO), Oxford, UK WorldWide Antimalarial Resistance Network (WWARN), Oxford, UK

**Abdoulaye Djimdé** — Université des Sciences, des Techniques et des Technologies de Bamako United States

**Arjen M Dondorp** — Mitigating Antimalarial Resistance Consortium for Southern and Eastern Africa (MARC SE-Africa), WorldWide Antimalarial Resistance Network (WWARN), Oxford, United Kingdom, Infectious Diseases Data Observatory (IDDO), Oxford, United Kingdom, Nuffield Department of Medicine, University of Oxford, Mahidol-Oxford Tropical Medicine Research Unit in Bangkok

**Annette Erhart** — London School of Hygiene and Tropical Medicine, Institute Of Tropical Medicine, Belgium

**Rick Fairhurst** — Oncology, AstraZeneca

**Abul Faiz** — Health Services, Dhaka, Dev Care Foundation, Dhaka, Bangladesh

**Huang Fang** — Shanghai Municipal Centre for Disease Control and Prevention, Shanghai, China

**Freya Fowkes** — Centre for Epidemiology and Biostatistics, Melbourne School of Population and Global Health, University of Melbourne, Melbourne, Australia, Disease Elimination Program, Burnet Institute, Melbourne, Australia, Department of Epidemiology and Preventive Medicine, Monash University, Melbourne, Australia

**Filbert Francis** — National Institute for Medical Research, Dar es Salaam, Tanzania

**Preteem Gandhi** — Novartis Pharma AG, Basel, Switzerland

**Mesia Kahunu Gauthier** — Unite De Pharmacologie Clinique, University Of Kinshasa, Democratic Republic of the Congo

**Samwel Gesase** — National Institute for Medical Research, Tanga Centre, PO Box 5004, Tanga, Tanzania

**Ousmane Guindo** — Epicentre, Niamey, Niger

**Felix Habarugira** — Department of Biomedical Laboratory Sciences, School of Health Sciences, College of Medicine and Health Sciences, University of Rwanda, Kigali P.O. Box 3286, Rwanda Pathology Department, Research Directorate, University Teaching Hospital of Butare, Huye P.O. Box 254, Rwanda

**Kasturi Haldar** — Department of Biological Sciences, Eck Institute of Global Health, University of Notre Dame, Notre Dame, Indiana 46556

**Mainga Hamaluba** — KEMRI-Wellcome Trust Research Programme, Kenya; Centre for Geographic Medicine Research (Coast), Kenya Medical Research Institute-Wellcome Trust Research Programme, Kilifi, Kenya

**Muzamil M Abdel Hamid** — Institute of Endemic Diseases, University of Khartoum

**Eun-Taek Han** — Kangwon National University

**Lucinda E Harrison** — Worldwide Antimalarial Resistance Network (WWARN), the University of Oxford, UK; Infectious Diseases Data Observatory, the University of Oxford, UK; Centre for Global Health Research, the University of Oxford, UK

**Anet Heart** — MRC Unit, The Gambia at the LSHTM, Atlantic Boulevard. POBOX273 Fajara. The Gambia

**Rupert Higgins** — The University of Oxford

**Deus S. Ishengoma** — Ifakara Health Institute, Dar es Salaam, Tanzania

**Hannah Jauncey** — Infectious Diseases Data Observatory (IDDO), University of Oxford, Oxford, UK

**Lucie Kafkova** — Infectious Diseases Data Observatory, University of Oxford, Oxford, United Kingdom, Centre for Tropical Medicine and Global Health, Nuffield Department of Medicine, University of Oxford, Oxford, United Kingdom

**Moses R Kamya** — Department of Medicine, Makerere University, Kampala, Uganda and Infectious Diseases Research Collaboration, Uganda

**Simon Kariuki** — Malaria Branch, Kenya Medical Research Institute (KEMRI)/U.S. Centres for Disease Control and Prevention (CDC) Research and Public Health Collaboration, Kenya

**Eline Kattenberg** — Department of Biomedical Sciences, Institute of Tropical Medicine, Antwerp, Belgium

**Sadie Kelly** — Infectious Diseases Data Observatory (IDDO), University of Oxford

**Aminatou Kone** — University of Bamako, Mali

**Ibrahim Maman Laminou** — Centre de Recherche Médicale et Sanitaire

**Jennifer Lee** — Infectious Diseases Data Observatory (IDDO), University of Oxford, Oxford, England, UK

**Pharath Lim** — Axle Research and Technologies, North Bethesda, Maryland, USA

**Jeffrey Livezey** — Walter Reed Army Institute of Research, Grifols Diagnostic Solutions Inc., San Diego, CA, USA

**Chanthap Lon** — Armed Forces Research Institute of Medical Sciences, Thailand

**Inke Nadia Diniyanti Lubis** — Faculty of Medicine, Universitas Sumatera Utara

**Celine Mandara** — National Institute for Medical Research, Dar es Salaam, Tanzania

**Andreas Mårtensson** — Global Health and Migration Unit, Department of Women's and Children's Health, Uppsala University, Uppsala, Sweden. Department of Infectious Diseases, Uppsala University Hospital, Uppsala, Sweden.

**Djibril Mbarushimana** — Djibril Mbarushimana, MM, MD University Teaching Hospital of Butare Hospital Avenue, Mamba Ngoma Huye, Rwanda

**Lwidiko E Mhamilawa** — Department of Women's and Children's Health, International Maternal and Child Health, Uppsala University, Uppsala, Sweden; Department of Parasitology and Medical Entomology, Muhimbili University of Health and Allied Sciences, Dar es Salaam, Tanzania

**Jules Ndoli Minega** — University Teaching Hospital of Butare

**Toshihiro Mita** — Department of Tropical Medicine and Parasitology, Faculty of Medicine, Juntendo University

**Eulambius Mlugu** — Muhimbili University of Health and Allied Sciences in the United Republic of Tanzania

**Frank Mockenhaupt** — Charité - Universitätsmedizin Berlin, Charité Center for Global Health, Institute of International Health

**Olugbenga Mokuolu** — Department of Paediatrics and Child Health, University of Ilorin, Ilorin, Nigeria. Centre for Malaria and Other Tropical Diseases Care, University of Ilorin Teaching Hospital, Ilorin, Nigeria

**Leah F Moriarty** — Centres for Disease Control & Prevention (CDC) Department of Health and Human Services (HHS)

**Hypolite Muhindo-Mavoko** — Department of Tropical Medicine, University of Kinshasa

**Mavuto Mukaka** — Medical Statistics and Epidemiology at University of Oxford (UK), Mahidol-Oxford Tropical Medicine Research Unit, Bangkok, Thailand

**Leon Mutesa** — Centre for Human Genetics and Genomics, College of Medicine and Health Sciences, University of Rwanda

**Joaniter Nankabirwa** — Infectious Diseases Research Collaboration, Kampala, Uganda, Makerere University College of Health Sciences, Kampala, Uganda

**Billy Ngasala** — Department of Parasitology & Medical Entomology, Muhimbili University of Health and Allied Sciences

**Chau Hoang Nguyen** — Oxford University Clinical Research Unit, Ho Chi Minh City, Vietnam

**Francois Nosten** — Centre for Tropical Medicine and Global Health MORU

**Myat Htut Nyunt** — Advanced Molecular Research Centre, Department of Medical Research, Ministry of Health, Yangon, Myanmar

**Henry Tony Oduor** — University of Oxford - IDDO

**Marie A Onyamboko** — Kinshasa School of Public Health, University of Kinshasa, Avenue Tombalbaye 68-78, Kinshasa, Democratic Republic of Congo

**Justine Padua** — Infectious Diseases Data Observatory, Oxford, UK | Centre for Tropical Medicine and Global Health, Nuffield Department of Medicine, Oxford, UK

**Rhys Peploe** — Infectious Diseases Data Observatory, University of Oxford, Oxford, United Kingdom. Centre for Tropical Medicine and Global Health, Nuffield Department of Medicine, University of Oxford, Oxford, United Kingdom

**Thomas Julian Peto** — Mahidol Oxford Tropical Medicine Research Unit, Faculty of Tropical Medicine, Mahidol University, Bangkok, Thailand, Centre for Tropical Medicine and Global Health, Nuffield Department of Medicine, University of Oxford, Oxford, UK

**Aung Pyae Phyo** — Shoklo Malaria Research Unit

**Jan Pierreux** — Department of General Internal Medicine and Infectious Diseases, Brussels University Hospital, Brussels 1000, Belgium

**Dylan R Pillai** — University of Calgary

**Riley Quah** — Infectious Diseases Data Observatory: Oxford, England, GB

**Anna Rosanas-Urgell** — Department of Biomedical Sciences, Institute of Tropical Medicine, Antwerp, Belgium

**Yurika Sakai** — Infectious Diseases Data Observatory (IDDO), Oxford, UK; WorldWide Antimalarial Resistance Network (WWARN), Oxford, UK

**Aaron Samuels** — CDC United States

**David Saunders** — Uniformed Services University School of Medicine

**Ester Schmitt** — Novartis Pharma AG, Basel, Switzerland

**Farhad Shokraneh** — Infectious Diseases Data Observatory, Oxford, UK; Centre for Tropical Medicine and Global Health, Nuffield Department of Medicine, University of Oxford, UK

**Issa Mahamat Souleymane** — Dr Issa Mahamat Souleymane, Chef de Service Laboratoire PNLP/Faculté des Sciences de la Santé Humaine-Université de N'Djaména

**Michele Spring** — Department of Microbiology and Immunology, State University of New York (SUNY) Upstate Medical University

**Sokunthea Sreng** — Cambodia National Malaria Centre, Cambodia, National Centre for Parasitology, Entomology and Malaria Control,

**Samantha Strudwick** — Infectious Diseases Data Observatory (IDDO), University of Oxford, Oxford, UK

**Sheila Suon** — Cambodia National Malaria Centre, Cambodia, National Centre for Parasitology, Entomology and Malaria Control,

**Pham Vinh Thanh** — National Institute of Malariology, Parasitology and Entomology, Hanoi, Vietnam

**Kamala Thriemer** — Global and Tropical Health Division, Menzies School of Health Research and Charles Darwin University, Darwin, NT, Australia

**Nhien Nguyen Thanh Thuy** — Oxford University Clinical Research Unit (OUCRU) in Vietnam

**Rupam Tripura** — Mahidol Oxford Tropical Medicine Research Unit (MORU), Faculty of Tropical Medicine, Mahidol University, Bangkok, Thailand Centre for Tropical Medicine and Global Health, Nuffield Department of Medicine, University of Oxford, Oxford, UK

**Rob W. van der Pluijm** — Mahidol-Oxford Tropical Medicine Research Unit, Faculty of Tropical Medicine, Mahidol University, Bangkok, Thailand. Centre for Tropical Medicine and Global Health, Nuffield Department of Medicine, University of Oxford, Oxford, UK. Infectious Disease Epidemiology and Analytics G5 Unit, Institut Pasteur, Université Paris Cité, Paris, France.

**Nguyen Van Hong** — National Institute of Malariology, Parasitology and Entomology (NIMPE), Hanoi, Vietnam

**Welmoed van Loon** — Charité Universitätsmedizin Berlin, Malaria & Infectious Disease Epidemiology at the Institute of International Health

**Thanh Ngo Viet** — Oxford University Clinical Research Unit, Wellcome Trust Major Overseas Programme, 764 Vo Van Kiet Street, Cho Quan Ward, Ho Chi Minh City

**Marian Warsame** — World Health Organisation (WHO, Switzerland)

**James Watson —** Infectious Diseases Data Observatory (IDDO), Oxford, UK; WorldWide Antimalarial Resistance Network (WWARN), Oxford, UK; Centre for Tropical Medicine and Global Health, Nuffield Department of Medicine, University of Oxford, Oxford, UK

**Nelli Westercamp** — Malaria Branch, U.S. Centres for Disease Control and Prevention

**Rakiswendé Serge Yerbanga** — Institut des Sciences et Techniques (INSTech) / Institut de Recherche en Sciences de la Santé (IRSS)

**Gashem Zamani** — KEMRI-Wellcome Trust Research Programme

**Core writing and scientific interpretation group**

**Philip J Rosenthal** — Professor of Pediatrics and Surgery, University of California, San Francisco, CA, USA

**Mehul Dhorda** — Mitigating Antimalarial Resistance Consortium for Southern and Eastern Africa (MARC SE-Africa); WorldWide Antimalarial Resistance Network (WWARN), Oxford, UK; Infectious Diseases Data Observatory (IDDO), Oxford, UK; Nuffield Department of Medicine, University of Oxford, Oxford, UK; Centre for Tropical Medicine and Global Health, Mahidol-Oxford Tropical Medicine Research Unit, Bangkok, Thailand

**Philippe J Guerin** — Infectious Diseases Data Observatory (IDDO), Oxford, UK; WorldWide Antimalarial Resistance Network (WWARN), Oxford, UK; Centre for Tropical Medicine and Global Health, Nuffield Department of Medicine, University of Oxford, Oxford, UK

**Karen Barnes** — Mitigating Antimalarial Resistance Consortium for Southern and Eastern Africa (MARC SE-Africa), Cape Town, South Africa; Collaborating Center for Optimizing Antimalarial Therapy (CCOAT), Division of Clinical Pharmacology, Department of Medicine, University of Cape Town, Cape Town, South Africa; WorldWide Antimalarial Resistance Network (WWARN), Oxford, UK; Infectious Diseases Data Observatory (IDDO), Oxford, UK

**Senior author**

**Prabin Dahal** — Infectious Diseases Data Observatory (IDDO), Oxford, UK; WorldWide Antimalarial Resistance Network (WWARN), Oxford, UK; Centre for Tropical Medicine and Global Health, Nuffield Department of Medicine, University of Oxford, Oxford, UK

**Supplementary file 1.**

**Contents:**

- **Figure S1:** PRISMA flow diagram of study selection for the IPDMA
- **Figure S2:** Study profile and data availability for parasite clearance analyses following treatment with artemisinin-based therapies.
- **Table S1:** Summary of studies from which the pooled data were included in this analysis
- **Table S2:** Genotyping results of included studies
- **Table S3:** Demographic summary of participants included in the pooled analysis
- **Supplementary references**

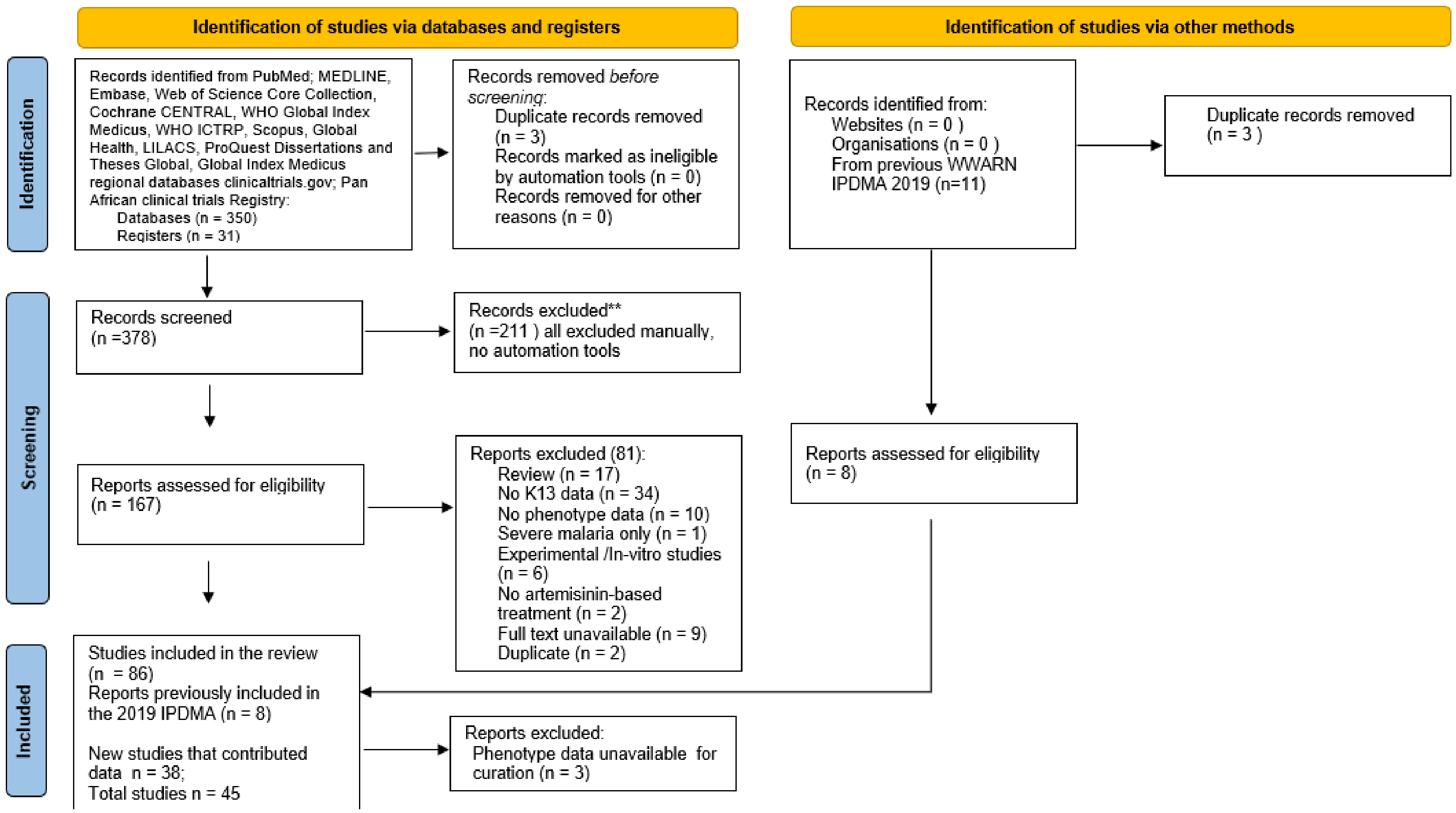

**Figure S1:** PRISMA flow diagram of study selection for the IPD meta-analysis. Studies were identified, screened, and assessed for eligibility according to PRISMA-IPD guidelines, with eligible studies invited to contribute individual participant data for pooled analysis.

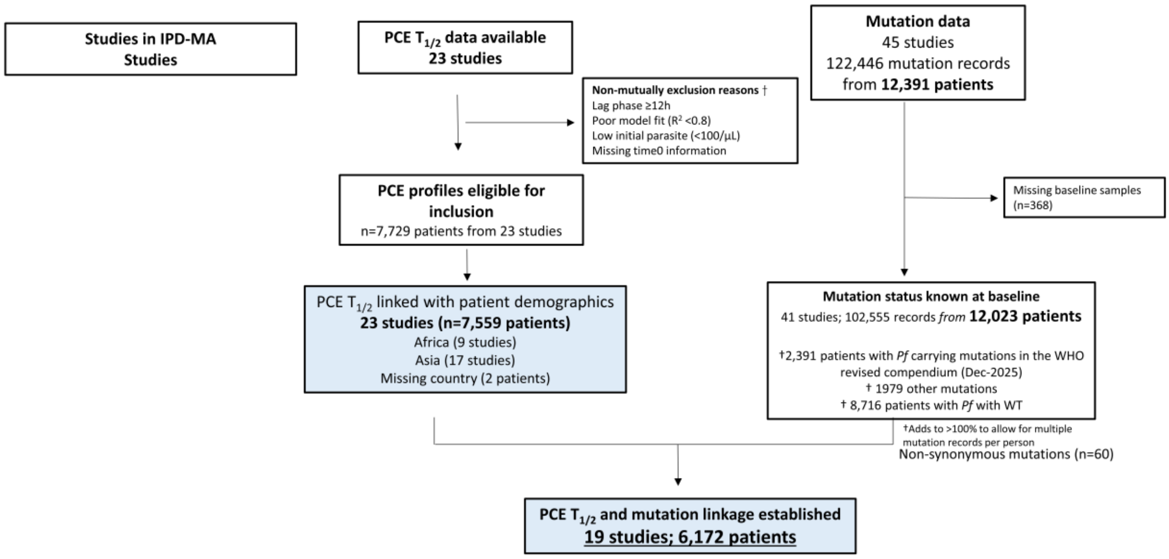

**Figure S2.** Study profile and data availability for parasite clearance analyses following treatment with artemisinin-based therapies. Definitions of specific exclusions are shown along with the numbers of studies, patients, and isolates included in each analysis. The figure summarises participants contributing to parasite clearance half-life (PC½) analyses and those with linked baseline *Kelch13* genotyping. Unsatisfactory fit was defined as a pseudo-$R^2$ <0.8. Insufficient parasite data included patients with too few parasitaemia measurements to reliably estimate PC½, including patients with only daily parasite counts.

**Table S1:** Summary of studies from which the pooled data were included in this analysis[1]

| | PCE[2] | Day 3 | Country[3] | Sites | Year | Participants | PUBMED or Clinical Trial ID. |
|---|---|---|---|---|---|---|---|
| 1 | NA | 240 | Somalia | Jowhar, Bosaso | 2016-2017 | 339 | 31296223 (1) |
| 2 | NA | 179 | India | Bhilai, Durg, Chhattisgarh | 2015-2017 | 179 | 33976269 (2) |
| 3 | 172; SS5 | 173 | Tanzania | Dar es Salaam | 2022 | 176 | 38352311 (3) |
| 4 | 166; SS3 | 92 | Vietnam | Binh Phuoc | 2010-2011 | 166 | 25180241 (4); 27036739 (5) |
| 5 | 62; SS5 | 73 | Ghana | Ga West Municipal Hospital, Amasaman, Oduman health centre, Mayera health centre | 2018 | 78 | 33163636 (6) |
| 6 | NA | NA | Multicenter | Gulu | 2017-2019 | 27 | 34666779 (7) |
| 7 | 40; SS3 | 7 | Thailand | Mae Sot | 2008 | 40 | 25180241 (4) |
| 8 | 79; SS3 | 36 | Cambodia | Pailin | 2008-2010 | 79 | 25180241 (4) |
| 9 | 1072; SS3 | 1037 | Cambodia; Bangladesh; India; Laos; Myanmar; Thailand; Vietnam; The Democratic Republic of the Congo | Cambodia (Pursat, Ratanakiri, Pailin, Preah Vihear); Bangladesh (Ramu); India (Agartala, Midnapur, Rourkela); Laos (Sekong); Myanmar (Ann); | 2015-2018 | 1110 | 32171078, (8) |

| | PCE[2] | Day 3 | Country[3] | Sites | Year | Participants | PUBMED or Clinical Trial ID. |
|---|---|---|---|---|---|---|---|
| | | | | Thailand (Khun Han, Phusing); Vietnam (Binh Phuoc); The Democratic Republic of the Congo | | | |
| 10 | NA | NA | Bangladesh; Cambodia; Cambodia; Cambodia; Cambodia; DRC; India; India; Laos; Myanmar; Myanmar; Myanmar; Myanmar; Thailand; Thailand; Vietnam | India (Agartala, Midnapur, Rourkela); Bangladesh (Ramu); Cambodia (Pailin, Preah Vihear, Pursat, Ratanakiri); DRC (Kinshasa); Laos (Sekong); Myanmar (Ann, Pyay, Pyin Oo lwin, Thabeikkyin); Thailand (Khun Han, Phusing); Vietnam (Binh Phuoc) | 2015-2018 | 143 | 32171078 (8); 25180241 (4) |

| | PCE[2] | Day 3 | Country[3] | Sites | Year | Participants | PUBMED or Clinical Trial ID. |
|---|---|---|---|---|---|---|---|
| 11 | 388; SS5 | 406 | Mali | Faladje, Bougoula-Hameau | 2015-2016 | 406 | 32320811 (9) |
| 12 | NA | 319 | Tanzania | Kibaha-Coast region, Mkuzi-Tanga, Mlimba-Morogoro, Ujiji-Kigoma | 2019 | 344 | 30898164 (10) |
| 13 | NA | 22 | Ghana | Accra | 2018 | 115 | 36803541 (11) |
| 14 | NA | 48 | Uganda | Gulu | 2014 | 61 | 28068997 (12) |
| 15 | NA | NA | Rwanda | Huye district | 2019 | 67 | 33350925 (13) |
| 16 | NA | 317 | Kenya | Siaya County, western Kenya | 2016-2017 | 320 | 32795367 (14) |
| 17 | NA | 302 | The Democratic Republic of Congo | Kapondo | 2017-2018 | 1356 | 34491220 (15) |
| 18 | NA | 91 | Myanmar | Kayin State, Chin State | 2013 | 91 | 25537878 (16) |
| 19 | 213; SS3 | 211 | Kenya | Kilifi | 2018-2019 | 218 | 34111412 (17) |
| 20 | 88; SS6 | 83 | Vietnam | Quang Nam Province | 2012-2013 | 95 | 25224002 (18) |
| 21 | NA | 74 | Indonesia | Batubara, Langkat, and South Nias | 2015 | 301 | 32420402 (19); 32393498 (20) |
| 22 | NA | 227 | Niger | Gaya, Tessaoua, Agadez | 2020 | 255 | 38741101 (21) |

| | PCE[2] | Day 3 | Country[3] | Sites | Year | Participants | PUBMED or Clinical Trial ID. |
|---|---|---|---|---|---|---|---|
| 23 | 297; SS3 | 305 | Cambodia; Vietnam | Stung Treng; Binh Phuoc | 2018-2020 | 312 | 35276064 (22); NCT03355664 |
| 24 | 1547; SS5 | 1638 | Bangladesh; Burkina Faso; Cambodia; Cambodia; DRC; Niger; Gambia; Nigeria; Republic of Guinea; Rwanda; Tanzania | Bangladesh (Cox's Bāzār, Chittagong, Bangladesh); Burkina Faso; Cambodia; Siem Pang, Stung Treng, Cambodia, ; DRC (Kingasani Health Center); Epicenter, Niger (Epicenter, Niger); Gambia (Fajara, City of Banjul, The Gambia); Nigeria (Ilorin, Kwara State, Nigeria); Republic of Guinea (Centre National de Recherche | 2020-2024 | 2686 | NCT03939104; NCT03923725 |

| | PCE[2] | Day 3 | Country[3] | Sites | Year | Participants | PUBMED or Clinical Trial ID. |
|---|---|---|---|---|---|---|---|
| | | | | et de Formation en Sante Rurale de Maferinyah); Rwanda (Kigali); Tanzania (Tnaga) | | | |
| 25 | NA | 50 | Multicenter | Mali, Uganda, Gabon, Ghana, Rwanda | 2018-2019 | 50 | 34410358 (23); 34216470 (24) |
| 26 | NA | 8 | Ethiopia | Sanja, Negade Bahir, Maksegnet, Aykel, Addis Zemen | 2009-2010 | 97 | 26483118 (25) |
| 27 | 58; SS6 | 45 | Vietnam | Krong Pa District | 2015-2017 | 60 | 32437557 (26) |
| 28 | NA | 8 | Africa travellers in Belgium | Belgium | 2022-2023 | 8 | 38157311 (27) |
| 29 | 114; SS1 | 114 | Myanmar | Thabeikkyin, Myitkyina | 2013-2014 | 114 | 27036739 (5) |
| 30 | NA | 662 | Vietnam | Bu Gia Map, Gia Lai, Phuoc Thang | 2011-2015 | 692 | 28086775 (28) |
| 31 | 43; SS3 | 33 | Laos | Savannakhet | 2010; 2014 | 44 | 25180241 (29) |
| 32 | 1818; SS1 | 1228 | Vietnam | Mawkertai, Maela, Mae Khon Ken, Wang Pha | 2001-2011 | 2013 | 28934435 (30) |

| | PCE[2] | Day 3 | Country[3] | Sites | Year | Participants | PUBMED or Clinical Trial ID. |
|---|---|---|---|---|---|---|---|
| 33 | 36; SS3 | 32 | Myanmar | Kayin state | 2013 | 36 | 29178921 (31) |
| 34 | 107; SS1 | 103 | Cambodia | Anlong Veng District Hospital, Oddar Meanchey Province | 2012-2014 | 119 | 25877962 (32) |
| 35 | 202; SS2 | 238 | Cambodia | Pursat, Preah Vihear, Ratanakiri | 2012-2013 | 241 | 26774243 (33); 25180241 (4) |
| 36 | NA | 75 | Sudan | Darelsalam, Elzohoor | 2015-2016 | 148 | 31034031(26) |
| 37 | 1167; SS1 | 1162 | Multicenter | Multicenter, 491 sites | 2011-2013 | 1234 | 25075834 (34); 28289193 (35) |
| 38 | 133; SS6 | 123 | China | Tengchong, Yingjiang | 2009-2012 | 142 | 25910630 (36) |
| 39 | 807; SS1 | 736 | Uganda | Agago, Busia, Arua | 2022-2023 | 808 | 40845863 (37); PACTR202301796134887 |
| 40 | NA | 215 | Chad | Kelo, Doba, Koyom | 2020-2021 | 215 | 37612601 (38) |
| 41 | NA | 80 | Uganda | Adjumani, | 2022 | 80 | 38712186 (39) |
| 42 | 222; SS1 | 240 | Uganda | Gulu, Northern Uganda | 2017-2019 | 240 | 34551228 (40) |
| 43 | NA | 903 | Sudan | South Darfur, Blue Nile, Gadaref, Sennar, West Darfur, Kassala | 2016-2020 | 948 | 37705047 (41) |
| 44 | NA | 231 | Tanzania | Ujiji, Kigoma; | 2011-2015 | 447 | 30333022 (42) |

| | PCE[2] | Day 3 | Country[3] | Sites | Year | Participants | PUBMED or Clinical Trial ID. |
|---|---|---|---|---|---|---|---|
| | | | | Kibaha, Pwani; Kilombero, Zanzibar North; Muheza, Tanga; Butimba, Mwanza; Chamwino, Dodoma; Nagaga, Mtwara; Kyela, Mbeya; Rufiji, Pwani | | | |
| 45 | NA | 106 | Mozambique | Massinga | 2018 | 114 | 34641867 (43) |

[1]Abbreviations: NA: Not available

[2]Parasite sampling schedules were classified as follows: SS1, 6-hourly until parasite-negative; SS2, 0, 2, 4, 6, 8, and 12 hours, then 6-hourly until parasite-negative; SS3, 0, 4, 6, 8, and 12 hours, then 6-hourly until parasite-negative; SS4, 0, 2, 4, 8, 16, and 24 hours, then 12-hourly until parasite-negative; SS5, 8-hourly until parasite-negative; and SS6, 12-hourly until parasite-negative.

[3]According to WHO (2025), artemisinin partial resistance has been confirmed in Rwanda (2020), Uganda (2021), Eritrea (2022), and the United Republic of Tanzania (2024), and is suspected in Ethiopia, Sudan, and Zambia. In the Greater Mekong Subregion, artemisinin partial resistance was first documented in Cambodia (2008–2009), followed by Thailand (2011–2012), Myanmar (2012–2014), Viet Nam (2015–2016), and the Lao People's Democratic Republic (2016–2017), and is now established throughout the region. Validated *Kelch13* resistance-associated mutations have also been reported in Guyana and Papua New Guinea.

**Table S2: Genotyping results of included studies**

| | Patients | Genotyped patients | Count genotypes | Mutations | WT (n) | Mutations (n) | Non-synonymous changes between | |
|---|---|---|---|---|---|---|---|---|
| | | | | | | | Codon 1-440 | Codon 441-726 |
| | | | | | | | N | N |
| 1 | 339 | 240 | 4 | M476I, R622I, V566I, WT | 240 | 3 | 0 | 3 |
| 2 | 179 | 179 | 3 | A675V, N29L, WT | 170 | 9 | 6 | 3 |
| 3 | 176 | 173 | 2 | A561H, WT | 134 | 39 | 0 | 39 |
| 4 | 166 | 92 | 7 | C580Y, I543T, P553L, P574L, V568G, WT, Y493H | 58 | 35 | 0 | 94 |
| 5 | 78 | 73 | 13 | A578S, A578V, F395S, F614S, K390R, L429S, M460V, N523S, R513H, S577P, V534A, W518L, WT | 52 | 22 | 2 | 35 |
| 6 | 27 | 27 | 2 | C580Y, WT | 24 | 3 | 0 | 3 |
| 7 | 40 | 7 | 5 | E252Q, N458Y, P527H, R561H, WT | 3 | 4 | 1 | 3 |
| 8 | 79 | 36 | 7 | A481V, C580Y, D584V, G449A, R539T, WT, Y493H | 2 | 34 | 1 | 33 |
| 9 | 1110 | 1995 | 29 | A578S, C580Y, D353N, | 1265 | 885 | 6 | 879 |

| | Patients | Genotyped patients | Count genotypes | Mutations | WT (n) | Mutations (n) | Non-synonymous changes between | |
|---|---|---|---|---|---|---|---|---|
| | | | | | | | Codon 1-440 | Codon 441-726 |
| | | | | | | | N | N |
| | | | | D399N, D464N, E567K, E668K, F446I, F662Y, G453S, G496D, G545R, G548V, G625E, G690D, G709D, G718S, H697N, K658R, N704T, Q613E, R365K, R539T, R561H, S695T, V637I, WT, Y493H, Y511H | | | | |
| 10 | 143 | 46 | 4 | C580Y, R539T, WT, Y493H | 8 | 38 | 0 | 38 |
| 11 | 406 | 406 | 10 | A578S, C469C, G548G, L440L, R404I, R471R, S364S, S459S, V581V, WT | 396 | 11 | 4 | 7 |
| 12 | 344 | 319 | 11 | A578S, C469C, E433D, I416V, P417P, | 302 | 17 | 4 | 13 |

| | Patients | Genotyped patients | Count genotypes | Mutations | WT (n) | Mutations (n) | Non-synonymous changes between | |
|---|---|---|---|---|---|---|---|---|
| | | | | | | | Codon 1-440 | Codon 441-726 |
| | | | | | | | N | N |
| | | | | Q613E, R471R, R471S, R539R, V487V, WT | | | | |
| 13 | 115 | 22 | 5 | C469C, K189N, K189T, L258M, R255K | 0 | 22 | 21 | 1 |
| 14 | 61 | 48 | 1 | WT | 48 | 0 | 0 | 0 |
| 15 | 67 | 66 | 7 | A578S, A675V, C469F, G533A, R561H, V555A, WT | 58 | 8 | 0 | 8 |
| 16 | 320 | 317 | 4 | A578S, C580C, S522C, WT | 311 | 9 | 1 | 8 |
| 17 | 1356 | 302 | 1 | WT | 303 | 0 | 0 | 0 |
| 18 | 91 | 91 | 16 | A481V, C580Y, D452E, D516Y, G449A, K479I, M476I, N458Y, N537I, P441L, R515T, R539T, R575K, R582T, V496F, WT | 62 | 29 | 0 | 29 |
| 19 | 218 | 211 | 6 | A486S, A578S, C542R, | 209 | 9 | 1 | 8 |

| | Patients | Genotyped patients | Count genotypes | Mutations | WT (n) | Mutations (n) | Non-synonymous changes between | |
|---|---|---|---|---|---|---|---|---|
| | | | | | | | Codon 1-440 | Codon 441-726 |
| | | | | | | | N | N |
| | | | | D399N, G665S, WT | | | | |
| 20 | 95 | 83 | 3 | I543T, WT, Y493H | 16 | 68 | 0 | 68 |
| 21 | 301 | 74 | 18 | D464A, D464G, E461K, E461N, F451L, F673L, L462F, L462I, L488V, M476I, N645A, Q468H, Q654L, S600F, T474A, T478A, WT, Y653C | 65 | 19 | 0 | 19 |
| 22 | 255 | 227 | 11 | C469C, E612E, E691E, G496G, G690G, K248K, N594K, P715P, R255K, V714S, WT | 211 | 16 | 5 | 11 |
| 23 | 312 | 305 | 5 | C580C, C580Y, R539T, WT, Y493H | 131 | 175 | 0 | 175 |
| 24 | 2686 | 1638 | 32 | A557S, A569V, A578S, A627V, A675V, A676S, | 1526 | 228 | 1 | 227 |

| | Patients | Genotyped patients | Count genotypes | Mutations | WT (n) | Mutations (n) | Non-synonymous changes between | |
|---|---|---|---|---|---|---|---|---|
| | | | | | | | Codon 1-440 | Codon 441-726 |
| | | | | | | | N | N |
| | | | | A724P, C469F, C532S, C580Y, E643K, E691D, G665S, K189T, L422I, M579I, N458D, N537I, N629Y, P441L, P475T, P574H, Q613H, R561H, S522C, S695T, T535K, V487I, V637I, V650F, WT, Y493H | | | | |
| 25 | 50 | 50 | 5 | P574L, P667S, Q661E, R561H, WT | 41 | 9 | 0 | 9 |
| 26 | 97 | 8 | 2 | R622I, V510V | 0 | 8 | 0 | 8 |
| 27 | 60 | 45 | 5 | C469F, C580Y, P553L, WT, Y511H | 7 | 39 | 0 | 39 |
| 28 | 8 | 8 | 6 | A675V, K189Y, R561H, S549P, WT | 2 | 8 | 3 | 6 |
| 29 | 114 | 114 | 7 | A676D, F446I, G538V, K189T, | 80 | 34 | 1 | 33 |

| | Patients | Genotyped patients | Count genotypes | Mutations | WT (n) | Mutations (n) | Non-synonymous changes between | |
|---|---|---|---|---|---|---|---|---|
| | | | | | | | Codon 1-440 | Codon 441-726 |
| | | | | | | | N | N |
| | | | | P443S, P574L, WT | | | | |
| 30 | 692 | 662 | 7 | C580Y, I543T, N485S, P553L, V568G, WT, Y493H | 379 | 283 | 0 | 283 |
| 31 | 44 | 33 | 1 | WT | 33 | 0 | 0 | 0 |
| 32 | 2013 | 1228 | 24 | A481V, A675V, C580Y, D281V, D516E, E252Q, F446I, G533A, G538V, K438N, M476I, N458Y, N537I, P441L, P527H, P527L, P574L, P667T, R239Q, R515K, R561H, S485N, S695S, WT | 890 | 338 | 107 | 231 |
| 33 | 36 | 32 | 6 | C580Y, F446I, I205T, M476I, R561H, WT | 17 | 15 | 14 | 1 |
| 34 | 119 | 103 | 2 | C580Y, R539T | 0 | 103 | 0 | 103 |
| 35 | 241 | 238 | 6 | C580Y, D584V, H719N, | 127 | 111 | 0 | 111 |

| | Patients | Genotyped patients | Count genotypes | Mutations | WT (n) | Mutations (n) | Non-synonymous changes between | |
|---|---|---|---|---|---|---|---|---|
| | | | | | | | Codon 1-440 | Codon 441-726 |
| | | | | | | | N | N |
| | | | | R539T, WT, Y493H | | | | |
| 36 | 148 | 75 | 8 | D389N, E433D, F375S, K378R, K430K, N594K, P443P, WT | 75 | 7 | 5 | 2 |
| 37 | 1234 | 1162 | 44 | A175T, A481V, A578S, A675V, C580Y, D281V, D584V, E252Q, F446I, F614L, G112E, G449A, G538V, G687G, H719N, I543T, K189N, K189T, K438N, K92N, L119L, L143P, M476I, N458Y, N525D, N537I, N725Y, N87K, P441L, P553L, P574L, P667L, Q613E, R223K, R255K, | 656 | 538 | 72 | 466 |

| | Patients | Genotyped patients | Count genotypes | Mutations | WT (n) | Mutations (n) | Non-synonymous changes between | |
|---|---|---|---|---|---|---|---|---|
| | | | | | | | Codon 1-440 | Codon 441-726 |
| | | | | | | | N | N |
| | | | | R471R, R539T, R561H, S522C, T149S, T535T, V568G, WT, Y493H | | | | |
| 38 | 142 | 123 | 11 | A481V, C469Y, C580Y, F446I, F483S, L492S, N458Y, N537I, P553L, P574L, WT | 52 | 71 | 0 | 19 |
| 39 | 808 | 736 | 6 | A569S, A578S, A675V, C469Y, C580Y, WT | 614 | 122 | 0 | 122 |
| 40 | 215 | 215 | 3 | A578S, N489Y, WT | 208 | 7 | 0 | 7 |
| 41 | 80 | 80 | 6 | A569S, A578S, A675V, C469Y, F491S, WT | 65 | 15 | 0 | 15 |
| 42 | 240 | 240 | 8 | A578S, A675V, C469Y, L708I, S695T, T685P, V555A, WT | 216 | 39 | 0 | 39 |
| 43 | 948 | 903 | 12 | A578S, A626S, C469Y, E461G, | 877 | 26 | 0 | 26 |

| | Patients | Genotyped patients | Count genotypes | Mutations | WT (n) | Mutations (n) | Non-synonymous changes between | |
|---|---|---|---|---|---|---|---|---|
| | | | | | | | Codon 1-440 | Codon 441-726 |
| | | | | | | | N | N |
| | | | | E620G, R561C, R561H, R622I, S600F, V494I, V589I, WT | | | | |
| 44 | 447 | 231 | 14 | C469C, E556K, E602D, G496S, G665G, L463S, M476I, M562T, R539R, V510M, V510V, V568V, V66V, WT | 219 | 14 | 1 | 12 |
| 45 | 114 | 106 | 1 | WT | 107 | 0 | 0 | 107 |

**Table S3: Demographic summary of participants included in the pooled analysis.**

| | Age category (years) | | | | |
|---|---|---|---|---|---|
| | <1 | 1-4 | 5-12 | >12 | Unknown |
| Overall | 328 (2·9%) | 2550 (22·5%) | 3542 (31·2%) | 4522 (39·9%) | 113 (3·5%) |
| Africa | 288 (4·3%) | 2498 (37·5%) | 3063 (46·0%) | 700 (10·5%) | 109 (1·6%) |
| Asia | 44 (0·8%) | 201 (3·6%) | 812 (14·4%) | 4293 (76·2%) | 286 (5·1%) |
| East Africa | 118 (2·8%) | 1813 (42·3%) | 1763 (41·2%) | 562 (13·1%) | 28 (0·7%) |
| Greater Mekong Sub-region | 44 (0·9%) | 184 (3·8%) | 646 (13·5%) | 3627 (75·8%) | 286 (6·0%) |
| Southern Africa | 20 (20·4%) | 78 (79·6%) | 0 | 0 | 0 |
| West Africa | 126 (6·1%) | 464 (22·6%) | 1252 (61·0%) | 129 (6·3%) | 81 (3·9%) |

| | | | | | |
|---|---|---|---|---|---|
| Africa (other) | 24 (10·7%) | 143 (63·8%) | 48 (21·4%) | 9 (4·0%) | 0 |
| Asia (other) | 0 | 17 (2·0%) | 166 (19·6%) | 666 (78·4%) | 0 |
| | **Transmission setting** | | | | |
| Very low | 0 | 29 (1·0%) | 309 (11·0%) | 2192 (78·0%) | 282 (10·0%) |
| Low | 57 (1·2%) | 598 (12·4%) | 1497 (31·1%) | 2653 (55·1%) | 8 (0·2%) |
| Moderate | 206 (6·2%) | 1375 (41·6%) | 1546 (46·8%) | 74 (2·2%) | 105 (3·2%) |
| High | 69 (5·1%) | 697 (51·2%) | 523 (38·4%) | 73 (5·4%) | 0 |

**Supplementary references**

1. Warsame M, Hassan AM, Hassan AH, Jibril AM, Khim N, Arale AM, et al. High therapeutic efficacy of artemether–lumefantrine and dihydroartemisinin–piperaquine for the treatment of uncomplicated falciparum malaria in Somalia. Malaria journal. 2019;18(1):231. 31296223

2. Das S, Kar A, Manna S, Mandal S, Mandal S, Das S, et al. Artemisinin combination therapy fails even in the absence of *Plasmodium falciparum kelch13* gene polymorphism in Central India. Scientific reports. 2021;11(1):9946.

3. Ishengoma, Deus S., et al. "Evidence of artemisinin partial resistance in northwestern Tanzania: clinical and molecular markers of resistance." *The Lancet Infectious Diseases* 24.11 (2024): 1225-1233.

4. Takala-Harrison S, Jacob CG, Arze C, Cummings MP, Silva JC, Dondorp AM, et al. Independent emergence of artemisinin resistance mutations among *Plasmodium falciparum* in Southeast Asia. J Infect Dis. 2015;211(5):670-9.

5. Tun KM, Jeeyapant A, Imwong M, Thein M, Aung SSM, Hlaing TM, et al. Parasite clearance rates in Upper Myanmar indicate a distinctive artemisinin resistance phenotype: a therapeutic efficacy study. Malaria journal. 2016;15(1):185.

6. Aninagyei E, Tetteh CD, Oppong M, Boye A, Acheampong DO. Efficacy of Artemether-Lumefantrine on various *Plasmodium falciparum Kelch 13* and Pfmdr1 genes isolated in Ghana. Parasite Epidemiology and Control. 2020;11:e00190.

7. Yoshida N, Yamauchi M, Morikawa R, Hombhanje F, Mita T. Increase in the proportion of *Plasmodium falciparum* with *kelch13* C580Y mutation and decline in pfcrt and pfmdr1 mutant alleles in Papua New Guinea. Malaria Journal. 2021;20(1):410.

8. van Der Pluijm RW, Tripura R, Hoglund RM, Phyo AP, Lek D, Ul Islam A, et al. Triple artemisinin-based combination therapies versus artemisinin-based combination therapies for uncomplicated *Plasmodium falciparum* malaria: a multicentre, open-label, randomised clinical trial. The Lancet. 2020;395(10233):1345-60.

9. Kone A, Sissoko S, Fofana B, Sangare CO, Dembele D, Haidara AS, *et al.* Different *Plasmodium falciparum* clearance times in two Malian villages following artesunate monotherapy. International Journal of Infectious Diseases. 2020;95:399-405.

10. Ishengoma DS, Mandara CI, Francis F, Talundzic E, Lucchi NW, Ngasala B, et al. Efficacy and safety of artemether-lumefantrine for the treatment of uncomplicated malaria and prevalence of *Pfk13* and Pfmdr1 polymorphisms after a decade of using artemisinin-based combination therapy in mainland Tanzania. Malaria Journal. 2019;18(1):88.

11. Ahorhorlu SY, Quashie NB, Jensen RW, Kudzi W, Nartey ET, Duah-Quashie NO, *et al*. Assessment of artemisinin tolerance in *Plasmodium falciparum* clinical isolates in children with uncomplicated malaria in Ghana. Malaria Journal. 2023;22(1):58.

12. Balikagala B, Mita T, Ikeda M, Sakurai M, Yatsushiro S, Takahashi N, et al. Absence of in vivo selection for *K13* mutations after artemether–lumefantrine treatment in Uganda. Malaria Journal. 2017;16(1):23.

13. Bergmann C, van Loon W, Habarugira F, Tacoli C, Jäger JC, Savelsberg D, et al. Increase in Kelch 13 polymorphisms in *Plasmodium falciparum*, southern Rwanda. Emerging Infectious Diseases. 2021;27(1):294.

14. Chebore W, Zhou Z, Westercamp N, Otieno K, Shi YP, Sergent SB, et al. Assessment of molecular markers of anti-malarial drug resistance among children participating in a therapeutic efficacy study in western Kenya. Malaria Journal. 2020;19(1):291.

15. Moriarty LF, Nkoli PM, Likwela JL, Mulopo PM, Sompwe EM, Rika JM, *et al*. Therapeutic efficacy of artemisinin-based combination therapies in Democratic Republic of the Congo and investigation of molecular markers of antimalarial resistance. The American Journal of Tropical Medicine and Hygiene. 2021;105(4):1067.

16. Nyunt MH, Hlaing T, Oo HW, Tin-Oo LL, Phway HP, Wang B, et al. Molecular assessment of artemisinin resistance markers, polymorphisms in the *k13* propeller, and a multidrug-resistance gene in the eastern and western border areas of Myanmar. Clin Infect Dis. 2015;60(8):1208-15.

17. Hamaluba M, van der Pluijm RW, Weya J, Njuguna P, Ngama M, Kalume P, et al. Arterolane–piperaquine–mefloquine versus arterolane–piperaquine and artemether–lumefantrine in the treatment of uncomplicated Plasmodium falciparum malaria in Kenyan children: a single-centre, open-label, randomised, non-inferiority trial. The Lancet Infectious Diseases. 2021;21(10):1395-406.

18. Thriemer K, Hong NV, Rosanas-Urgell A, Phuc BQ, Ha do M, Pockele E, *et al*. Delayed parasite clearance after treatment with dihydroartemisinin-piperaquine in *Plasmodium falciparum* malaria patients in central Vietnam. Antimicrob Agents Chemother. 2014;58(12):7049-55.

19. Lubis IND, Wijaya H, Lubis M, Lubis CP, Beshir KB, Staedke SG, et al., editors. Recurrence of Plasmodium malariae and *P. falciparum* following treatment of uncomplicated malaria in North Sumatera with dihydroartemisinin-piperaquine or artemether-lumefantrine. Open Forum Infectious Diseases; 2020: Oxford University Press US.

20. Lubis IN, Wijaya H, Lubis M, Lubis CP, Beshir KB, Sutherland CJ. Plasmodium falciparum Isolates Carrying *pf k13* Polymorphisms Harbor the SVMNT Allele of pfcrt in Northwestern Indonesia. Antimicrobial Agents and Chemotherapy. 2020;64(8):10.1128/aac. 02539-19.

21. Laminou IM, Issa I, Adehossi E, Maman K, Jackou H, Coulibaly E, *et al.* Therapeutic efficacy and tolerability of artemether–lumefantrine for uncomplicated *Plasmodium falciparum* malaria in Niger, 2020. Malaria Journal. 2024;23(1):144.

22. Peto TJ, Tripura R, Callery JJ, Lek D, Nghia HDT, Nguon C, et al. Triple therapy with artemether–lumefantrine plus amodiaquine versus artemether–lumefantrine alone for

artemisinin-resistant, uncomplicated falciparum malaria: an open-label, randomised, multicentre trial. The Lancet Infectious Diseases. 2022;22(6):867-78.

23. Schmitt EK, Ndayisaba G, Yeka A, Asante KP, Grobusch MP, Karita E, et al. Efficacy of cipargamin (KAE609) in a randomized, phase II dose-escalation study in adults in sub-Saharan Africa with uncomplicated *Plasmodium falciparum* malaria. Clinical Infectious Diseases. 2022;74(10):1831-9.

24. Straimer J, Gandhi P, Renner KC, Schmitt EK. High prevalence of *Plasmodium falciparum K13* mutations in Rwanda is associated with slow parasite clearance after treatment with artemether-lumefantrine. The Journal of Infectious Diseases. 2022;225(8):1411-4.

25. Bayih AG, Getnet G, Alemu A, Getie S, Mohon AN, Pillai DR. A unique *Plasmodium falciparum Kelch 13* gene mutation in northwest Ethiopia. The American Journal of Tropical Medicine and Hygiene. 2016;94(1):132.

26. Rovira-Vallbona E, Van Hong N, Kattenberg JH, Huan RM, Hien NTT, Ngoc NTH, et al. Efficacy of dihydroartemisinin/piperaquine and artesunate monotherapy for the treatment of uncomplicated *Plasmodium falciparum* malaria in Central Vietnam. Journal of Antimicrobial Chemotherapy. 2020;75(8):2272-81.

27. Pierreux J, Bottieau E, Florence E, Maniewski U, Bruggemans A, Malotaux J, *et al*. Failure of artemether-lumefantrine therapy in travellers returning to Belgium with *Plasmodium falciparum* malaria: an observational case series with genomic analysis. Journal of Travel Medicine. 2024;31(3):taad165.

28. Thanh NV, Thuy-Nhien N, Tuyen NTK, Tong NT, Nha-Ca NT, Dong LT, et al. Rapid decline in the susceptibility of *Plasmodium falciparum* to dihydroartemisinin–piperaquine in the south of Vietnam. Malaria journal. 2017;16(1):27.

29. Takala-Harrison S, Jacob CG, Arze C, Cummings MP, Silva JC, Dondorp AM, et al. Independent emergence of artemisinin resistance mutations among *Plasmodium falciparum* in Southeast Asia. The Journal of Infectious Diseases. 2015;211(5):670-9.

30. Ataíde R, Powell R, Moore K, McLean A, Phyo AP, Nair S, et al. Declining transmission and immunity to malaria and emerging artemisinin resistance in Thailand: a longitudinal study. The Journal of Infectious Diseases. 2017;216(6):723-31.

31. Bonnington CA, Phyo AP, Ashley EA, Imwong M, Sriprawat K, Parker DM, *et al. Plasmodium falciparum Kelch 13* mutations and treatment response in patients in Hpa-Pun District, northern Kayin State, Myanmar. Malaria journal. 2017;16(1):480.

32. Spring MD, Lin JT, Manning JE, Vanachayangkul P, Somethy S, Bun R, et al. Dihydroartemisinin-piperaquine failure associated with a triple mutant including *kelch13* C580Y in Cambodia: an observational cohort study. Lancet Infect Dis. 2015;15(6):683-91.

33. Amaratunga C, Lim P, Suon S, Sreng S, Mao S, Sopha C, et al. Dihydroartemisinin–piperaquine resistance in *Plasmodium falciparum* malaria in Cambodia: a multisite prospective cohort study. The Lancet infectious diseases. 2016;16(3):357-65.

34. Ashley EA, Dhorda M, Fairhurst RM, Amaratunga C, Lim P, Suon S, *et al*. Spread of artemisinin resistance in *Plasmodium falciparum* malaria. New England Journal of Medicine. 2014;371(5):411-23.

35. Ataide R, Ashley EA, Powell R, Chan J-A, Malloy MJ, O'Flaherty K, et al. Host immunity to *Plasmodium falciparum* and the assessment of emerging artemisinin resistance in a multinational cohort. Proceedings of the National Academy of Sciences. 2017;114(13):3515-20.

36. Huang F, Takala-Harrison S, Jacob CG, Liu H, Sun X, Yang H, et al. A Single Mutation in K13 Predominates in Southern China and Is Associated With Delayed Clearance of Plasmodium falciparum Following Artemisinin Treatment. J Infect Dis. 2015;212(10):1629-35.

37. Kamya MR, Nankabirwa JI, Ebong C, Asua V, Kiggundu M, Orena S, et al. Efficacies of Artemether-Lumefantrine, Artesunate-Amodiaquine, Dihydroartemisininpiperaquine and Artesunate-Pyronaridine for the Treatment of Uncomplicated *Plasmodium falciparum* Malaria in Children in Uganda: A Randomized, Open-Label Phase IV Clinical Trial. 2025. The Lancet Infectious Diseases, 26(1), 67-78.

38. Issa MS, Warsame M, Mahamat MHT, Saleh IDM, Boulotigam K, Djimrassengar H, et al. Therapeutic efficacy of artesunate–amodiaquine and artemether–lumefantrine for the treatment of uncomplicated falciparum malaria in Chad: clinical and genetic surveillance. Malaria Journal. 2023;22(1):240.

39. Angwe MK, Mwebaza N, Nsobya SL, Vudriko P, Dralabu S, Omali D, et al. Day 3 parasitemia and *Plasmodium falciparum Kelch 13* mutations among uncomplicated malaria patients treated with artemether-lumefantrine in Adjumani district, Uganda. Plos one. 2024;19(6):e0305064.

40. Balikagala B, Fukuda N, Ikeda M, Katuro OT, Tachibana S-I, Yamauchi M, et al. Evidence of artemisinin-resistant malaria in Africa. New England Journal of Medicine. 2021;385(13):1163-71.

41. Adam M, Nahzat S, Kakar Q, Assada M, Witkowski B, Tag Eldin Elshafie A, et al. Antimalarial drug efficacy and resistance in malaria-endemic countries in HANMAT-PIAM_net countries of the Eastern Mediterranean Region 2016–2020: Clinical and genetic studies. Tropical Medicine & International Health. 2023;28(10):817-29.

42. Kakolwa MA, Mahende MK, Ishengoma DS, Mandara CI, Ngasala B, Kamugisha E, et al. Efficacy and safety of artemisinin-based combination therapy, and molecular markers for artemisinin and piperaquine resistance in Mainland Tanzania. Malaria journal. 2018;17(1):369.

43. Chidimatembue A, Svigel SS, Mayor A, Aíde P, Nhama A, Nhamussua L, et al. Molecular surveillance for polymorphisms associated with artemisinin-based combination therapy resistance in *Plasmodium falciparum* isolates collected in Mozambique, 2018. Malaria journal. 2021;20(1):398.

**Contents**

- **Table S4:** Distribution of included studies and patients for Day 2 and Day 3 parasite positivity
- **Supplementary Results (Text S1) Day 2 and Day 3 Parasite Positivity**
- **Table S5:** Association between *Kelch13* mutation classification and Day 3 parasitaemia in patients with uncomplicated falciparum malaria
- **Supplementary Results (Text S2)**: Association between geographic region, resistance status, transmission setting and Day 3 parasitaemia in patients with falciparum malaria
- **Table S6:** Association between geographic region and Day 3 parasitaemia in patients with falciparum malaria
- **Figure S4:** Study-specific crude associations between *Plasmodium falciparum Kelch13* mutation status and Day 3 parasite positivity
- **Table S7:** Cross-reference between forest plot labels, study characteristics, and corresponding publication
- **Figure S5:** Study-specific crude associations between *Kelch13* status and parasite clearance half-life
- **Figure S6:** Day 3 parasitaemia by country resistance status.
- **Supplementary text S3:** Exploratory effect-modification analysis by transmission intensity.
- **Supplementary References 2**

**Supplementary file 2**

**Table S4:** Distribution of included studies and patients for Day 2 and Day 3 parasite positivity by region and transmission settings, with linked clinical and *Kelch13* genotype data[1]

| **Region** | **Studies** | **Participants** | **Hyper-parasitaemic** | **Day 2 positivity** | **Day 3 positivity** |
|---|---|---|---|---|---|
| Africa | 25 | 6659 | 26% (1752/6659) | 50% (2041/4066) | 4% (159/4337) |
| Asia | 20 | 5637 | 48% (2729/5637) | 81% (3829/4749) | 46% (1507/3253) |
| South East Asia | 17 | 4788 | 52% (2492) | 75% (3591) | 31% (1469) |
| Rest of Asia | 6 | 850 | 26% (225) | 25% (215) | 25% (213) |
| East Africa | 19 | 4285 | 26% (1132) | 35% (1498) | 3% (127) |
| Horn of Africa | 2 | 1226 | 15% (187) | 14% (171) | 0.08%% (1) |
| West Africa | 8 | 2053 | 29% (595) | 63% (1299) | 1% (29) |
| Southern Africa | 1 | 98 | 26% (25) | 10% (10) | 0% (0) |
| Rest of Africa | 3 | 225 | 0% (0) | 96% (215) | 1% (3) |
| Total | 42[2] | 12294 | 36% (4481) | 67% (5870) | 21% (1666) |
| **Transmission setting** | | | | | |
| Very low | 12 | 2621 | 34% (901) | 53% (1380) | 23% (606) |
| Low | 21 | 4782 | 43% (2057) | 57% (2746) | 18% (878) |
| Moderate | 15 | 3287 | 30% (988) | 23% (772) | 1% (44) |
| High | 8 | 1347 | 31% (423) | 59% (792) | 5% (67) |

[1]Hyperparasitaemia was defined as a baseline (0 h) asexual *Plasmodium falciparum* parasitaemia >100,000 parasites/µL by microscopy.

[2]Several studies included multiple regions

**Supplementary text 1 Day 2 and Day 3 Parasite Positivity**

Regionally, Southeast Asia (SEA) contributed 4,788 patients and showed high positivity on Day 2 and Day 3 (75% [3,591] and 31% [1,469], respectively). In contrast, East Africa (4,285) had lower positivity (35% [1,498] and 3% [127] Days 2 and 3, respectively), while West Africa (2,053) showed high Day 2 but minimal Day 3 positivity (63% [1,299] and 1% [29]). In the rest of Asia (850), Day 2 and Day 3 positivity rates were similar (25% [215] and 25% [213], respectively). Smaller contributions from the Horn of Africa (1,226), Southern Africa (98), and Africa-unspecified studies (225) showed considerable heterogeneity in parasite positivity.

*Kelch13* mutation proportions increased across WHO ART-R country classification strata at the time of patient treatment (1), from 10·4% (95% CI 9·7–11·1) in settings where resistance was not confirmed, to 26·3% (95% CI 24·4–28·4) in settings with suspected resistance, and to 56·3% (95% CI 54·5–58·1) in settings with confirmed resistance. This was paralleled by increasing Day 2 positivity (57·0%, 81·3%, and 83·4%, respectively), while Day 3 positivity was markedly higher in confirmed resistance settings (61·3% (1022/1666) confirmed vs 24·8% (414/1666) not confirmed and 13·8% (230/1666) suspected).

In the pooled dataset, the prevalence of *Kelch13* mutations and WHO ART-R classification showed a pronounced age gradient, increasing from 9·3% (95% CI, 8·2–10·4) in children aged 1–<4 years to 40·7% (95% CI, 39·3–42·0) in those aged <12 years. This was accompanied by a shift from predominantly unconfirmed resistance (92·1%) in younger children to confirmed resistance (47·5%) in adults, together with markedly higher Day 3 positivity in adults (41·8% vs <10% among participants aged <12 years). This pattern is consistent with the geographic and transmission-setting distribution of the dataset, with higher resistance burdens in Asia and in low-transmission settings that include older participant populations.

Across transmission settings, the largest number of participants were from low-transmission settings (4,782), followed by moderate (3,287), very low (2,621), and high (1,347). Day 2 positivity was highest in high- and low-transmission settings (59% [792] and 57% [2,746], respectively), followed by very low settings (53% [1,380]), and lowest in moderate settings (23% [772]). In contrast, Day 3 positivity was highest in very low-transmission settings (23% [606]) and low-transmission settings (18% [878]), and lower in high-transmission settings (5% [67]) and moderate-transmission settings (1% [44]).

Day 2 and day 3 parasite positivity across *Kelch13* mutation classes, from wild type and other mutations to WHO potential, candidate, and validated mutations. Excluding baseline hyperparasitaemia reduced absolute positivity estimates but did not alter the overall gradient, indicating that the association between *Kelch13* status and post-treatment parasite persistence was not solely driven by high baseline parasite density. Notably, the WHO potential mutations showed high Day 3 positivity, approaching that of candidate and validated mutations, and this pattern persisted after excluding baseline hyperparasitaemia (**Figure 4; main text**).

Parasite positivity was higher among mutant *Kelch13* genotypes than wild-type infections on both Day 2 and Day 3 (92·5% vs 55·5% at 48h; 63·9% vs 5·8% at 72h). In multivariable Day

3 models adjusted for age category, geographic region, transmission setting, and baseline hyperparasitaemia, WHO-validated, candidate, and potential *Kelch13* mutations remained strongly associated with parasite positivity compared with wild-type (**Table S5**), including WHO-potential mutations (adjusted OR 27·1, 95% CI 12·0–61·5; p<0·001), WHO-validated mutations (19·8, 16·6–23·6; p<0·001), and WHO-candidate mutations (13·7, 8·9–21·3; p<0·001). Infections classified as "other" mutations were also associated with increased odds of day 3 positivity (2·45, 1·64–3·67; p<0·001).

Baseline hyperparasitaemia (>100,000 asexual *P. falciparum* parasites per µL) was independently associated with increased odds of Day 3 positivity (adjusted OR 2·12, 95% CI 1·80–2·50; p<0·001). Compared with individuals aged <12 years, younger age groups did not show increased odds of Day 3 positivity, while individuals with unknown age had reduced odds (0·59, 0·43–0·81; p=0·001). Marked regional differences were observed, with substantially lower odds of Day 3 positivity in Africa compared with Asia (0·18, 0·13–0·25; p<0·001)· Transmission setting was not associated with Day 3 positivity in multivariable analysis after adjustment for age category, geographic region (Africa vs Asia), *Kelch13* mutation category, and baseline hyperparasitaemia (AOR: 0·88, 0·61–1·25; p=0·47).

**Table S5:** Association between *Kelch13* mutation classification and Day 3 parasitaemia in patients with uncomplicated falciparum malaria[1]

| Variable | n | Day 3 positive n (%) | Adjusted OR | 95% CI | p-value |
|---|---|---|---|---|---|
| ***Kelch13* classification** | | | | | |
| Wild-type (ref) | 5,475 | 316 (5·8%) | 1·00 | – | – |
| WHO-Validated | 1,691 | 1,189 (70·3%) | 19·8 | 16·6–23·6 | <0·001 |
| WHO-Candidate | 120 | 89 (74·2%) | 13·7 | 8·9–21·3 | <0·001 |
| WHO-Potential | 45 | 37 (82·2%) | 27·1 | 12·0–61·5 | <0·001 |
| Other | 256 | 34 (13·3%) | 2·45 | 1·64–3·67 | <0·001 |
| **Hyperparasitaemia** | | | | | |
| No (ref) | 5,138 | 808 (15·7%) | 1·00 | – | – |
| Yes | 2,449 | 857 (35·0%) | 2·12 | 1·80–2·50 | <0·001 |
| **Age category (years)** | | | | | |
| <12 (ref) | 2,95 | 1,234 (41·8%) | 1·00 | – | – |

| | | | | | |
|---|---|---|---|---|---|
| 1–<4 | 1,751 | 111 (6·3%) | 1·29 | 0·93–1·80 | 0·12 |
| 4–<12 | 2,304 | 228 (9·9%) | 0·91 | 0·72–1·15 | 0·41 |
| Unknown | 582 | 92 (15·8%) | 0·59 | 0·43–0·81 | 0·001 |
| **Region** | | | | | |
| Asia (ref) | 3,252 | 1,507 (46·3%) | 1·00 | – | – |
| Africa | 4,335 | 158 (3·6%) | 0·18 | 0·13–0·25 | <0·001 |
| **Transmission setting** | | | | | |
| Low and very-low (ref) | 4,611 | 1,554 (33·7%) | 1·00 | – | – |
| Moderate and high | 2,976 | 111 (3·7%) | 0·88 | 0·61–1·25 | 0·47 |

[1]Odds ratios are adjusted for age category, geographic region (Africa vs Asia), transmission setting, and baseline hyperparasitaemia. Wild type, no hyperparasitaemia, age <12 years, Asia, and low/very low transmission settings were used as reference categories. OR = odds ratio; CI = confidence interval.

**Supplementary Text S2**

In more granular regional analyses, all African regions showed substantially lower odds of Day 3 parasitaemia than in Southeast Asia (SEA), consistent with the broader Africa-Asia comparison (**Table S6**).

**Table S6:** Association between geographic region and Day 3 parasitaemia in patients with falciparum malaria[1]

| Region | n | Day 3 positive n (%) | Adjusted OR | 95% CI | p-value |
|---|---|---|---|---|---|
| **SEA (ref)** | 2,825 | 1,469 (52·0%) | 1·00 | – | – |
| Asia (non-SEA) | 427 | 38 (8·9%) | 0·088 | 0·062–0·124 | <0·001 |
| East Africa | 3,159 | 127 (4·0%) | 0·041 | 0·030–0·057 | <0·001 |

| | | | | | |
|---|---|---|---|---|---|
| West Africa | 965 | 29 (3·0%) | 0·034 | 0·021–0·057 | <0·001 |

[1]Odds ratios are adjusted for age category, transmission setting, and baseline hyperparasitaemia. The South East Asia (SEA), was used as the reference group. Asia (non-GMS) includes all Asian study sites outside the SEA. Observations from sites without a specific regional classification were excluded from this analysis. OR = odds ratio

To contextualise the relationship between mutation classification and parasite clearance, we examined Day 3 positivity across resistance categories. In analyses adjusted for age category, geographic region, transmission setting, and baseline hyperparasitaemia, confirmed resistance was associated with increased odds of Day 3 parasitaemia (adjusted OR 2·33, 95% CI 1·96–2·78), whereas suspected resistance was associated with lower odds (0·49, 0·40–0·60). This likely reflects the heterogeneous nature of suspected resistance, including high-transmission settings where partial immunity facilitates parasite clearance despite resistance-associated mutations.

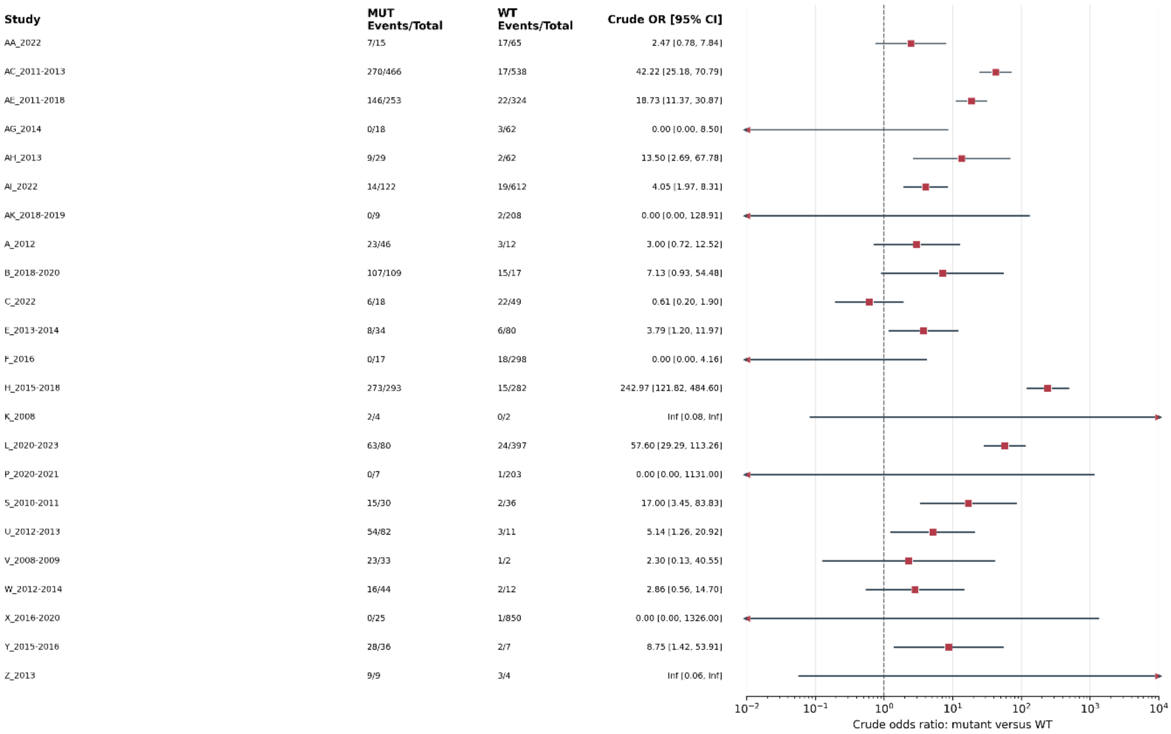

**Supplementary Figure S4. Study-specific crude associations between *Plasmodium falciparum Kelch13* mutation status and Day 3 parasite positivity**

Forest plot showing study-specific crude odds ratios (ORs) for Day 3 parasite positivity among participants with Kelch13 mutant (MUT) versus wild-type (WT) infections. Events and totals are presented for each group. Squares represent study-specific ORs, and horizontal lines represent 95% confidence intervals (CIs). The dashed vertical line indicates the null value (OR = 1), and arrowheads indicate CIs extending beyond the displayed range. ORs > 1 indicate higher odds of Day 3 positivity among participants with mutant infections. Exact conditional estimates were used for informative studies with zero cells; consequently, some estimates or confidence limits are reported as zero or infinity (Inf). Studies are identified with anonymised labels, with the sampling year appended, and can be cross-referenced with Table S1 (Supplementary file 1; Table S7 below). No pooled estimate is shown because the primary synthesis used a one-stage adjusted individual participant data model.

**Table S7: Cross-reference between forest plot labels, study characteristics, and corresponding publication**

| Sampling year | Forest label | Country | Table S1 row | PubMed/clinical-trial identifier and Ref listed in Supplementary file Table S1 |
|---|---|---|---|---|
| 2012 | A_2012 | Vietnam | 20 | 25224002 (18) |
| 2018–2020 | B_2018-2020 | Cambodia; Vietnam | 23 | 35276064 (22); NCT03355664 |
| 2022 | C_2022 | Tanzania | 3 | 38352311 (3) |
| 2013–2014 | E_2013-2014 | Myanmar | 29 | 27036739 (5) |
| 2001–2011 | G_2001-2011 | Myanmar; Thailand | 32 | 28934435 (30) |
| 2015–2018 | H_2015-2018 | Bangladesh; Cambodia; +6 more | 9 | 32171078 (8) |
| 2008 | K_2008 | Thailand | 7 | 25180241 (4) |
| 2020–2023 | L_2020-2023 | Bangladesh; Burkina Faso; +8 more | 24 | NCT03939104; NCT03923725 |
| 2019 | N_2019 | Uganda | 42 | 34551228 (40) |
| 2017 | O_2017 | Ghana | 5 | 33163636 (6) |

| | | | | |
|---|---|---|---|---|
| 2010–2011 | S_2010-2011 | Vietnam | 4 | 25180241 (4); 27036739 (5) |
| 2012–2013 | U_2012-2013 | Cambodia | 35 | 26774243 (33); 25180241 (4) |
| 2008–2009 | V_2008-2009 | Cambodia | 8 | 25180241 (4) |
| 2012–2014 | W_2012-2014 | People's Republic of China | 38 | 25910630 (36) |
| 2015–2016 | Y_2015-2016 | Vietnam | 27 | 32437557 (26) |
| 2013 | Z_2013 | Myanmar | 33 | 29178921 (31) |
| 2011–2013 | AC_2011-2013 | Bangladesh; Cambodia; +6 more | 37 | 25075834 (34); 28289193 (35) |
| 2011–2018 | AE_2011-2018 | Vietnam | 30 | 28086775 (28) |
| 2015–2017 | AF_2015-2017 | Mali | 11 | 32320811 (9) |
| 2022 | AI_2022 | Uganda | 39 | 40845863 (37); PACTR202301796134887 |
| 2018–2019 | AK_2018-2019 | Kenya | 19 | 34111412 (17) |
| 2013 | AO_2013* | Cambodia | 34 | 25877962 (32) |
| 2010–2014 | AP_2010-2014* | Laos | 31 | 25180241 (29) |

| Study | MUT n; GM PC½ (h) | WT n; GM PC½ (h) | Crude GMR [95% CI] |
|---|---|---|---|
| A_2012 | 67; 5.77 | 16; 6.42 | 0.90 [0.80, 1.01] |
| B_2018-2020 | 168; 6.52 | 128; 2.98 | 2.19 [1.96, 2.44] |
| C_2022 | 39; 3.63 | 130; 4.04 | 0.90 [0.77, 1.06] |
| E_2013-2014 | 34; 4.35 | 80; 2.66 | 1.63 [1.36, 1.96] |
| G_2001-2011 | 55; 3.61 | 110; 3.24 | 1.11 [0.96, 1.30] |
| H_2015-2018 | 470; 6.29 | 564; 2.51 | 2.50 [2.36, 2.65] |
| K_2008 | 4; 5.90 | 3; 3.47 | 1.70 [1.12, 2.57] |
| L_2020-2023 | 127; 5.74 | 1308; 3.05 | 1.88 [1.70, 2.08] |
| N_2019 | 33; 2.59 | 199; 1.78 | 1.46 [1.19, 1.78] |
| O_2017 | 20; 4.27 | 42; 4.27 | 1.00 [0.93, 1.08] |
| S_2010-2011 | 34; 6.87 | 58; 2.65 | 2.59 [2.21, 3.04] |
| U_2012-2013 | 100; 7.85 | 100; 3.66 | 2.15 [1.85, 2.49] |
| V_2008-2009 | 34; 6.37 | 2; 3.57 | 1.78 [0.03, 92.64] |
| W_2012-2014 | 71; 4.96 | 49; 3.06 | 1.62 [1.41, 1.86] |
| Y_2015-2016 | 39; 6.94 | 6; 2.82 | 2.46 [1.84, 3.29] |
| Z_2013 | 15; 3.68 | 17; 3.06 | 1.20 [0.78, 1.85] |
| AC_2011-2013 | 504; 5.04 | 600; 2.53 | 1.99 [1.88, 2.10] |
| AE_2011-2018 | 239; 5.77 | 360; 2.46 | 2.35 [2.18, 2.53] |
| AF_2015-2017 | 10; 2.49 | 378; 2.21 | 1.13 [0.97, 1.30] |
| AI_2022 | 122; 4.17 | 613; 2.98 | 1.40 [1.29, 1.52] |
| AK_2018-2019 | 7; 2.45 | 195; 2.62 | 0.94 [0.65, 1.34] |

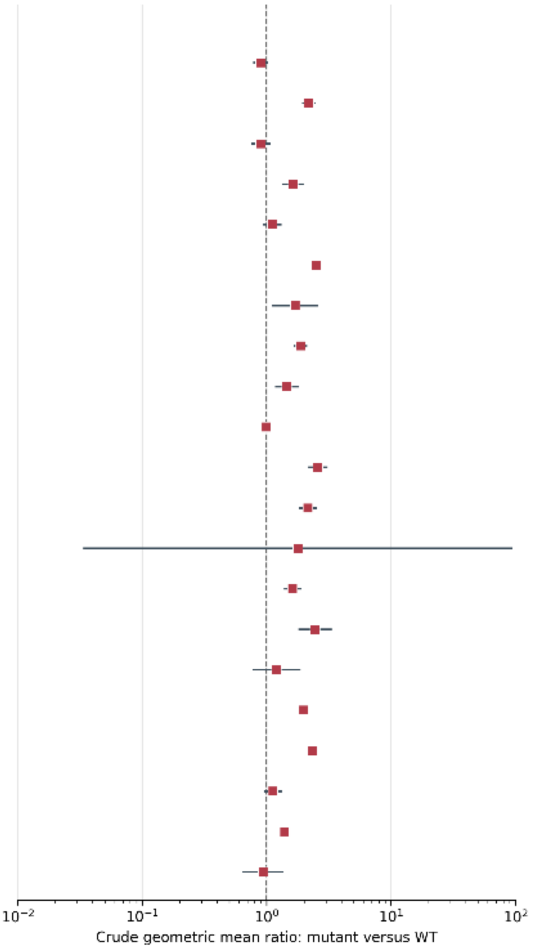

**Supplementary Figure S5. Study-specific crude associations between *Kelch13* status and parasite clearance half-life**

Forest plot showing study-specific crude geometric mean ratios (GMRs) for parasite clearance half-life (PC½) in participants with *Kelch13* mutant (MUT) versus wild-type (WT) infections. Group sample sizes and geometric mean PC½ values are presented. Squares represent GMRs, horizontal lines represent 95% confidence intervals (CIs), and the dashed line indicates no difference (GMR = 1). A GMR greater than 1 indicates a longer PC½ in mutant infections. Of 7,285 participants with linked *Kelch13* and valid PC½ data, 7,150 from 21 studies contributed to estimable comparisons. Two studies comprising 135 participants were not plotted because they contained only MUT infections (n = 103) or only WT infections (n = 32). A total of 106 participants with both WT and MUT records were classified as MUT according to the "any detected mutation" rule. The overall PC½ population reported elsewhere (n = 7,791) represents the broader PC½ analysis population rather than the *Kelch13*-linked population used for this figure. PC½ values were analysed on the natural-log scale, and 95% CIs were calculated using Welch's two-sample method.

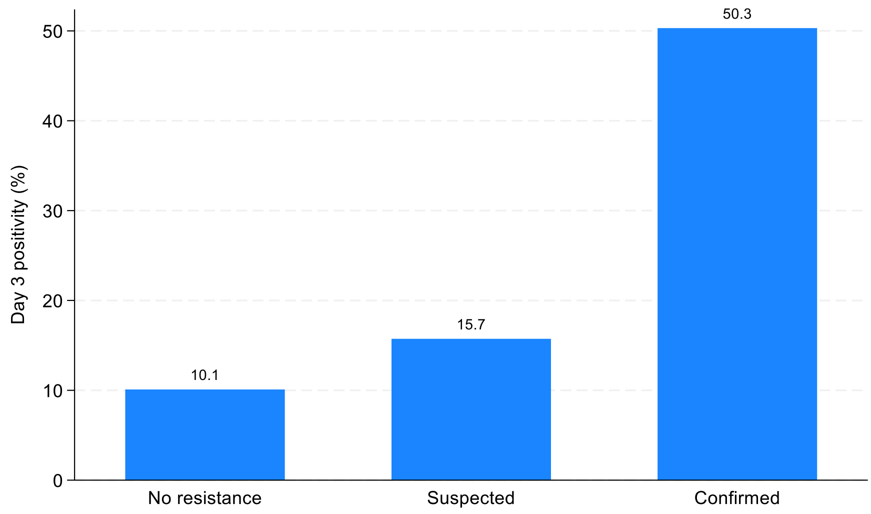

**Figure S6:** Day 3 parasitaemia by country resistance status. The proportion of patients with Day 3 positivity increased progressively from sites with no resistance (10%; 414/4093) to suspected resistance (16%; 230/1463) and confirmed resistance (50%; 1021/2031), demonstrating a clear gradient consistent with the emergence and consolidation of artemisinin resistance

Resistance-confirmed observations were almost exclusively derived from Asian settings, with 99·5% (2875/2922) of confirmed cases originating from Asia. This strong geographical clustering indicates that resistance status and region are not independent, reflecting the temporal emergence of ART-R. As such, resistance status and region were modelled separately to disentangle biological and epidemiological effects.

**Supplementary text S3:Exploratory effect-modification analysis by transmission intensity.** Among 7,464 participants from 521 study sites, transmission intensity modified the association between *Kelch13* mutation status and Day 3 positivity (ratio of odds ratios for moderate/high versus very low/low transmission: 0.26; 95% CI, 0.14–0.50; p for interaction = 0.0001). *Kelch13* mutations were associated with greater odds of Day 3 positivity in both settings, but the association was stronger in very low/low transmission settings (adjusted OR 18.42, 95% CI 13.67–24.81) than in moderate/high transmission settings (adjusted OR 4.82, 95% CI 2.69–8.62). Site-level heterogeneity was substantial (ICC 0.65, 95% CI 0.57–0.72; random-intercept variance 6.01). The direction of association was positive in both pooled transmission strata, although the magnitude differed between them.

**Strength of evidence.** Evidence was strongest for associations of WHO-validated *Kelch13* mutations with Day 3 positivity and prolonged parasite-clearance half-life (PC½), supported by large participant numbers and relatively precise estimates. The Day 3 association remained positive in both transmission settings, although it was attenuated in moderate- to high-transmission settings. Evidence for candidate, potential, and newly identified mutations was less certain because of smaller sample sizes and wider confidence intervals.

**Supplementary References 2**

1. World Health Organization. The World Malaria Report 2025

**Supplementary file 3**

**Contents**

- **Supplementary text 4 (S4):** Correspondence between $PC_1/_2$ >5h and Day 3 parasitaemia
- **Table S8:** Diagnostic of individual performance of Day 3 parasitaemia for detecting delayed parasite clearance (PC½ >5h).
- **Supplementary Figure S7.** Predicted probability of Day 3 parasitaemia by parasite clearance half-life ($PC_1/_2$), overall and stratified by transmission intensity.
- **Supplementary text S5.** Correspondence between Day 3 positivity and $PC_1/_2$ >5 h
- **Table S9:** Diagnostic performance of Day 2 parasitaemia

**Supplementary text 4 (S4):** Correspondence between $PC_1/_2$ >5h and Day 3 parasitaemia

Stratification by baseline parasitaemia demonstrated substantial variation in diagnostic performance. Among patients without hyperparasitaemia, sensitivity was 68.9% (639/928) and specificity was 97.9% (1716/1753), whereas among those with hyperparasitaemia, sensitivity increased to 85.9% (480/559) with lower specificity of 92.6% (1098/1186).

Regional differences were also observed, with sensitivity higher in Asia (81.6%, 1042/1277) than in Africa (36.7%, 77/210), despite higher specificity in Africa (97.7% vs 86.6%). These differences were largely explained by transmission intensity. In very low and low transmission settings, sensitivity was 80.8% (1076/1331; 95% CI 78.6–82.8), compared with 27.6% (43/156; 95% CI 20.7–35.8) in moderate and high transmission settings, where specificity was correspondingly higher (98.8%, 1512/1530; 95% CI 98.1–99.3).

These findings indicate that Day 3 parasitaemia performance is strongly influenced by the epidemiological context, with reduced sensitivity in higher-transmission settings likely reflecting the effects of host immunity and infection complexity on parasite clearance.

**Table S8: Diagnostic of individual performance of Day 3 parasitaemia for detecting delayed parasite clearance (PC½ >5h).**

| | TP | FN | FP | TN | Sensitivity % (95% CI) | Specificity % (95% CI) |
|---|---|---|---|---|---|---|
| Overall | 1119 | 368 | 125 | 2814 | 75.3 (73.0–77.0) | 95.7 (95.0–96.0) |
| No hyperparasitaemia | 639 | 289 | 37 | 1716 | 68.9 (65.8–72.0) | 97.9 (97.1–98.5) |
| Hyperparasitaemia | 480 | 79 | 88 | 1098 | 85.9 (82.7–88.7) | 92.6 (90.9–94.0) |
| Africa | 77 | 133 | 21 | 1543 | 36.7 (30.1–43.5) | 98.7 (98.0–99.2) |
| Asia | 1042 | 235 | 104 | 1271 | 81.6 (79.4–83.7) | 92.4 (90.9–93.7) |
| Very low/low transmission | 1076 | 255 | 107 | 1302 | 80.8 (78.6–82.8) | 92.4 (90.9–93.7) |
| Moderate/high transmission | 43 | 113 | 18 | 1512 | 27.6 (20.7–35.8) | 98.8 (98.1–99.3) |

True positives (TP) represent patients with both Day 3 positivity and delayed parasite clearance; false negatives (FN) represent patients with delayed parasite clearance but no

Day 3 positivity; false positives (FP) represent patients with Day 3 positivity but without delayed parasite clearance; and true negatives (TN) represent patients with neither Day 3 positivity nor delayed parasite clearance. Sensitivity and specificity are presented as percentages with 95% confidence intervals. Hyperparasitaemia was defined as baseline parasitaemia >100,000 parasites per *μL*. Transmission settings were classified as very low/low or moderate/high. Regional categories were defined according to study site classification.

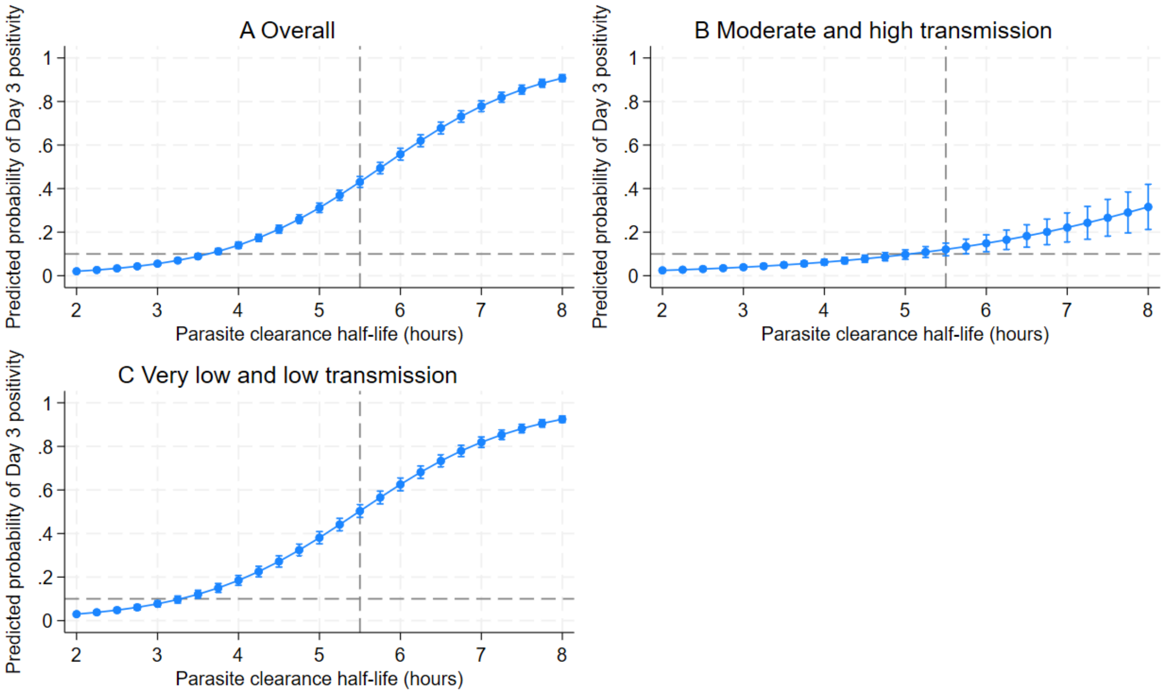

**Supplementary Figure S7. Predicted probability of Day 3 parasitaemia by parasite clearance half-life ($PC_1/_2$), overall and stratified by transmission intensity.**

Predicted probabilities of Day 3 parasitaemia were estimated from logistic regression models across the pooled dataset. The relationship between parasite clearance half-life and Day 3 positivity is shown for **(A)** the overall dataset, **(B)** moderate- and high-transmission settings, and **(C)** very low- and low-transmission settings. Predicted probabilities increased nonlinearly with increasing $PC_1/_2$, but the relationship differed across transmission settings. At comparable $PC_1/_2$ values, predicted Day 3 positivity was lower in moderate and high transmission settings than in very low and low transmission settings. Error bars represent 95% confidence intervals.

**Supplementary text S5. Correspondence between Day 3 positivity and $PC_1/_2$ >5 h**

Day 2 parasitaemia was highly sensitive but poorly specific for delayed parasite clearance. Day 2 positivity identified 98% (1867/1902) of patients with PC1/2 >5h, but specificity was only 44.4% (1771/3991), indicating that many patients without delayed clearance remained parasite-positive at 48h. Stratification by baseline parasitaemia showed similar findings. Among patients without hyperparasitaemia, sensitivity was 97.7% (1232/1261) and specificity was 47% (1051/2229), whereas among those with hyperparasitaemia, sensitivity increased to 99.1% (635/641) but specificity declined to 41% (720/1762). Performance also differed by transmission setting. In very low and low transmission settings, sensitivity was 99% (1643/1658; 95% CI 98.5–99.5) and specificity was 37.1% (754/2031; 95% CI 35.0–39.1). In moderate- and high-transmission settings, sensitivity was lower at 92% (224/244; 95% CI, 87.6–95.0), whereas specificity increased modestly to 51.9% (1017/1960; 95% CI, 49.6–54.1). These findings indicate that although Day 2 parasitaemia is a highly sensitive marker of delayed clearance, it is a poor discriminator of clinically meaningful resistance, as many infections that remain parasite-positive at 48 h clear by 72 h.

**Table S9: Diagnostic performance of Day 2 parasitaemia[1]**

| Stratum | TP | FN | FP | TN | Sensitivity % (95% CI) | Specificity % (95% CI) |
|---|---|---|---|---|---|---|
| Overall | 1867 | 35 | 2220 | 1771 | 98.2 (97.4–98.8) | 44.4 (42.8–46.0) |
| No hyperparasitaemia | 1232 | 29 | 1178 | 1051 | 97.7 (96.7–98.4) | 47.2 (45.1–49.3) |
| Hyperparasitaemia | 635 | 6 | 1042 | 720 | 99.1 (98.0–99.7) | 40.9 (38.6–43.2) |
| Very low/low transmission | 1643 | 15 | 1277 | 754 | 99.1 (98.5–99.5) | 37.1 (35.0–39.1) |
| Moderate/high transmission | 224 | 20 | 943 | 1017 | 91.8 (87.6–95.0) | 51.9 (49.6–54.1) |

[1]True positives (TP) represent patients with both Day 2 positivity and delayed parasite clearance; false negatives (FN) represent patients with delayed parasite clearance but no Day 2 positivity; false positives (FP) represent patients with Day 2 positivity but without delayed parasite clearance; and true negatives (TN) represent patients with neither Day 2 positivity nor delayed parasite clearance. Sensitivity and specificity are presented as percentages with 95% confidence intervals. Hyperparasitaemia was defined as baseline parasitaemia >100,000 parasites/μL. Transmission settings were classified as very low/low or moderate/high.

**Supplemental file 4: Additional results summarising parasite clearance slope half-life (PC½)**

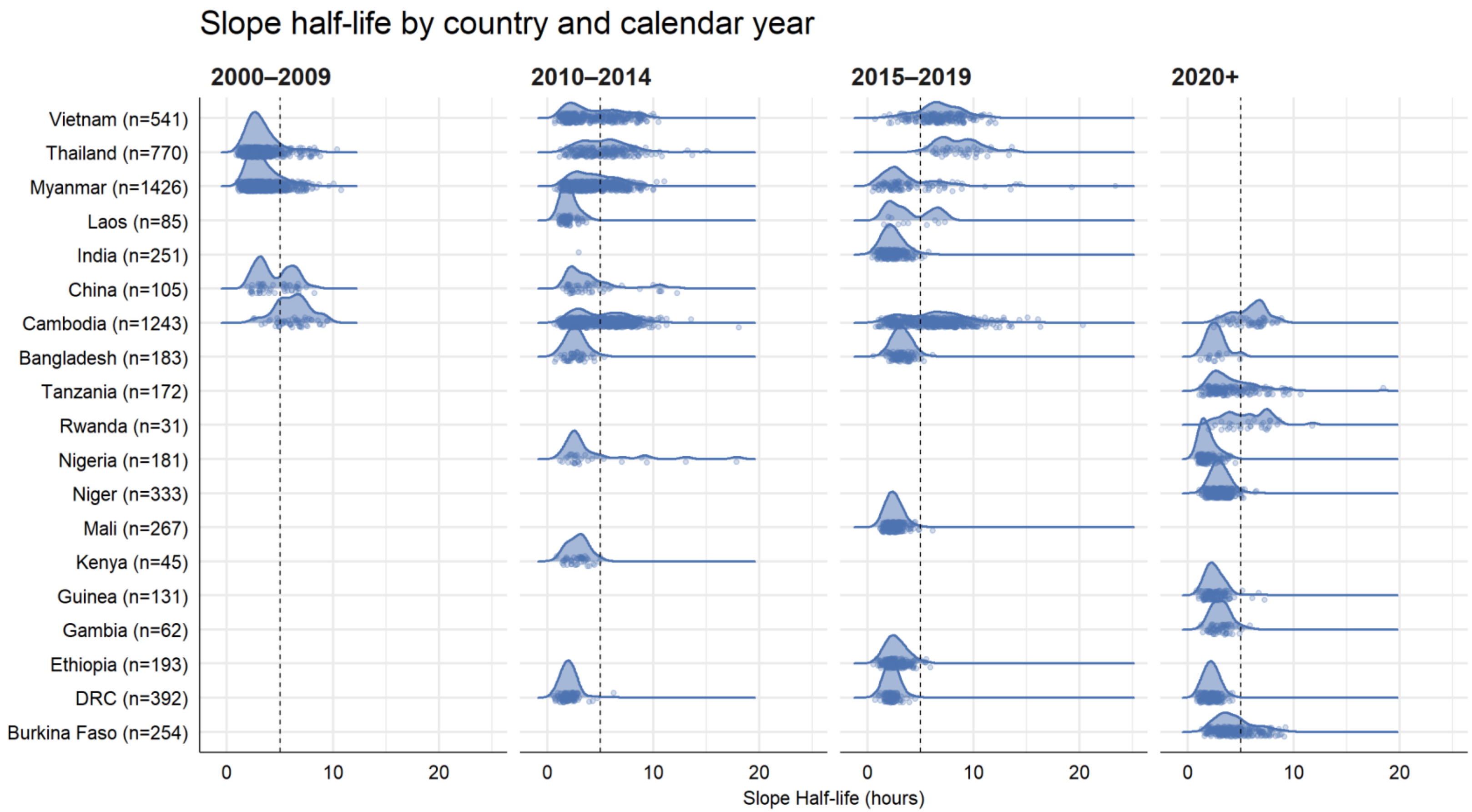

**S Figure 4.1**: Distribution of slope half-life by country and calendar period.

**Legend:** Vertical line shown at 5 hours. DRC = Democratic Republic of Congo. Data for each country is displayed as jitter. Ghana is excluded from the plot as there were only 3 patients.

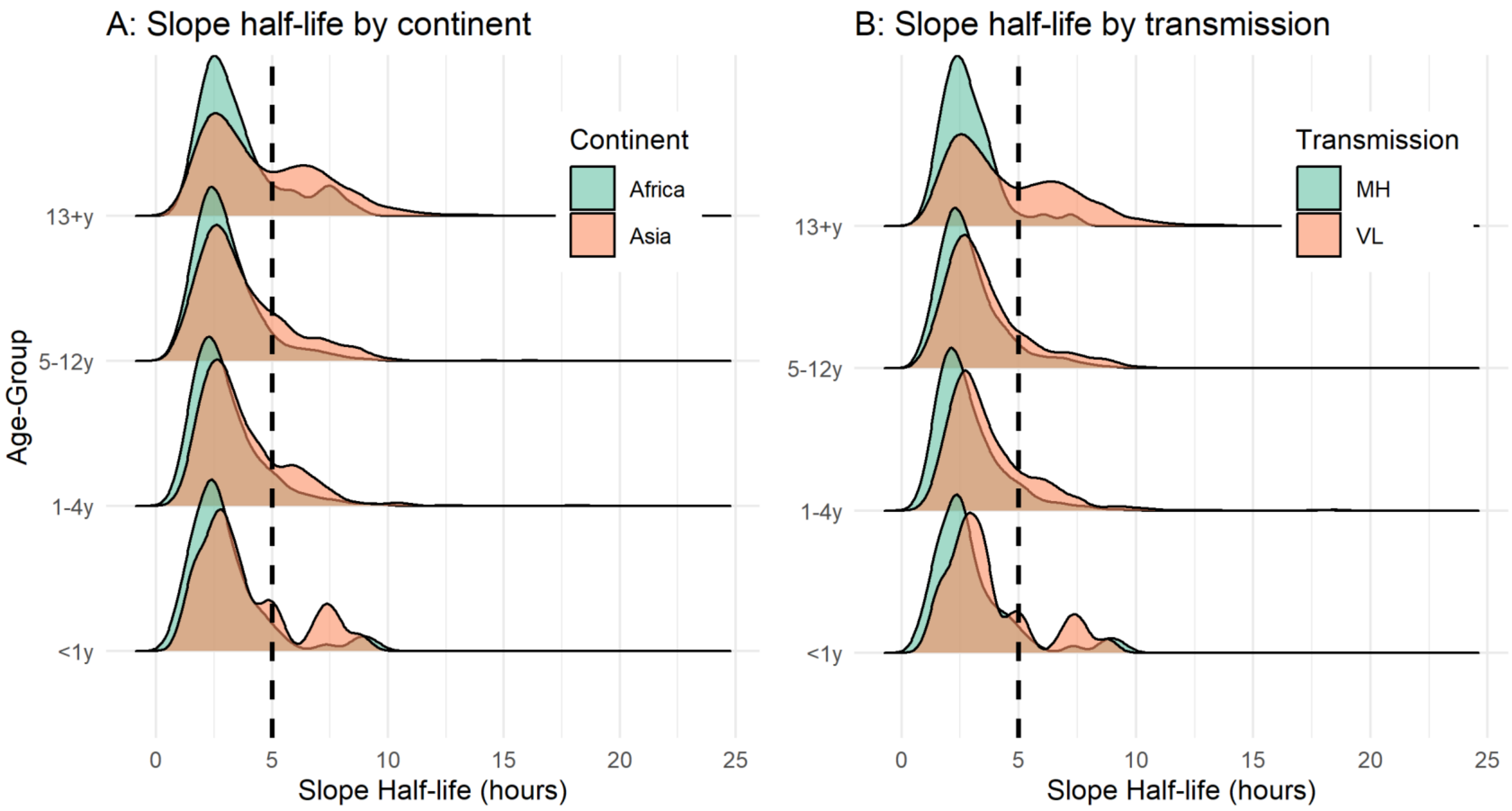

**S Figure 4.2**: Distribution of slope half-life by transmission, continent, and age-group.
MH = moderate-to-high transmission settings; VL = low-to-very-low transmission settings. Vertical dotted line shown at 5h.

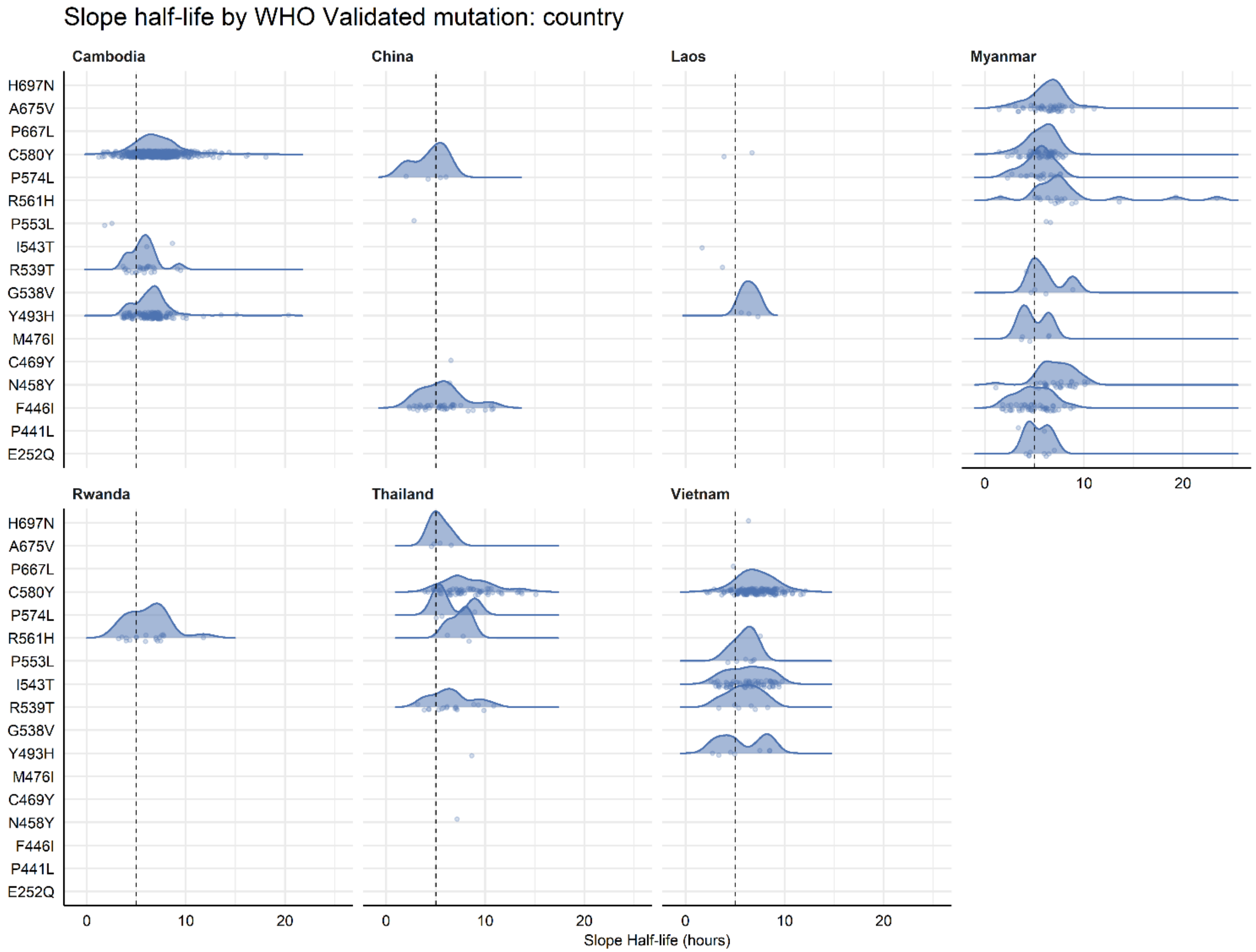

**S Figure 4.3**: Distribution of slope half-life for WHO-Validated mutations, by country.
Legend: Vertical dotted line shown at 5h.

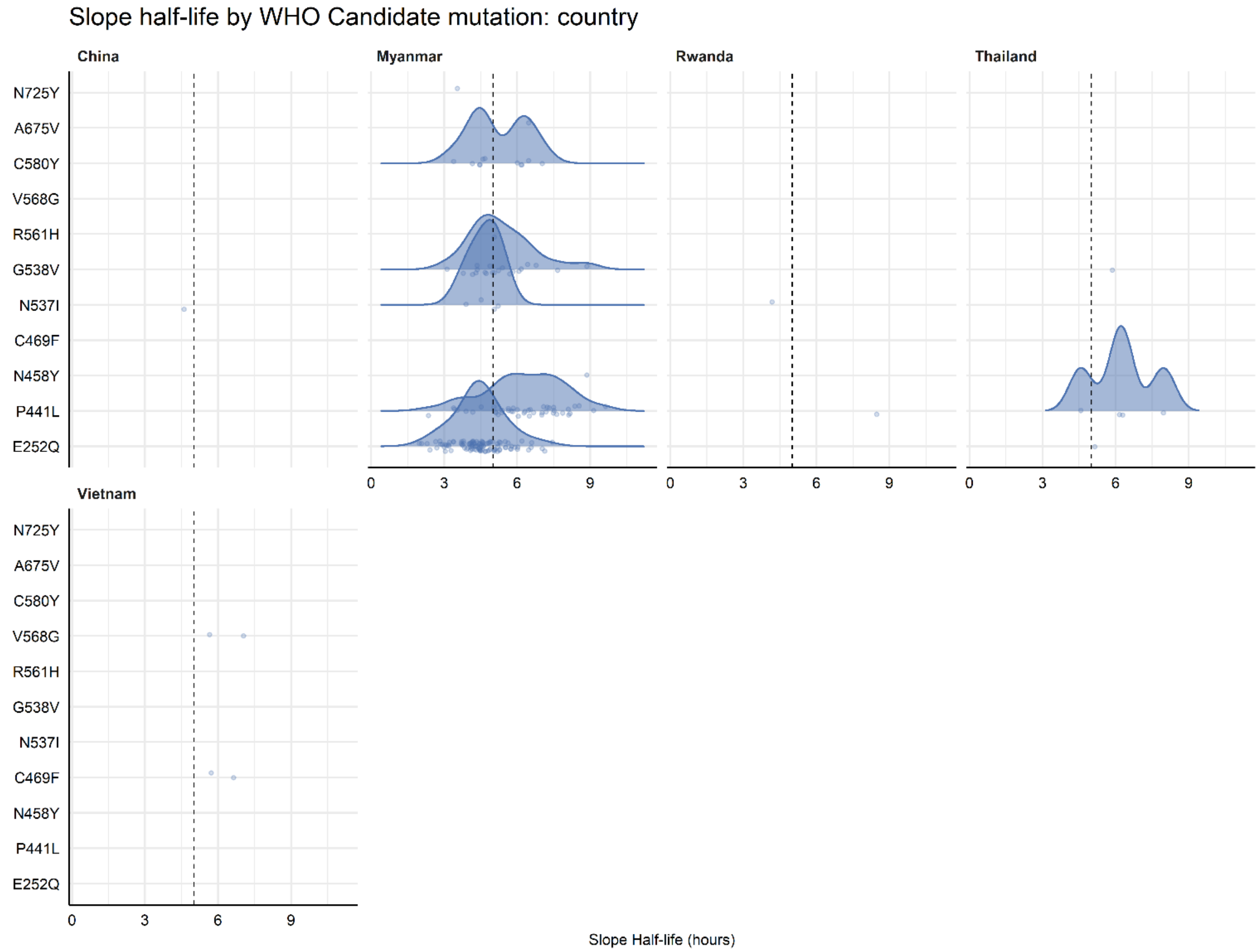

**S Figure 4.4**: Distribution of slope half-life for WHO candidate mutations, by country.
Legend: Vertical dotted line shown at 5h.

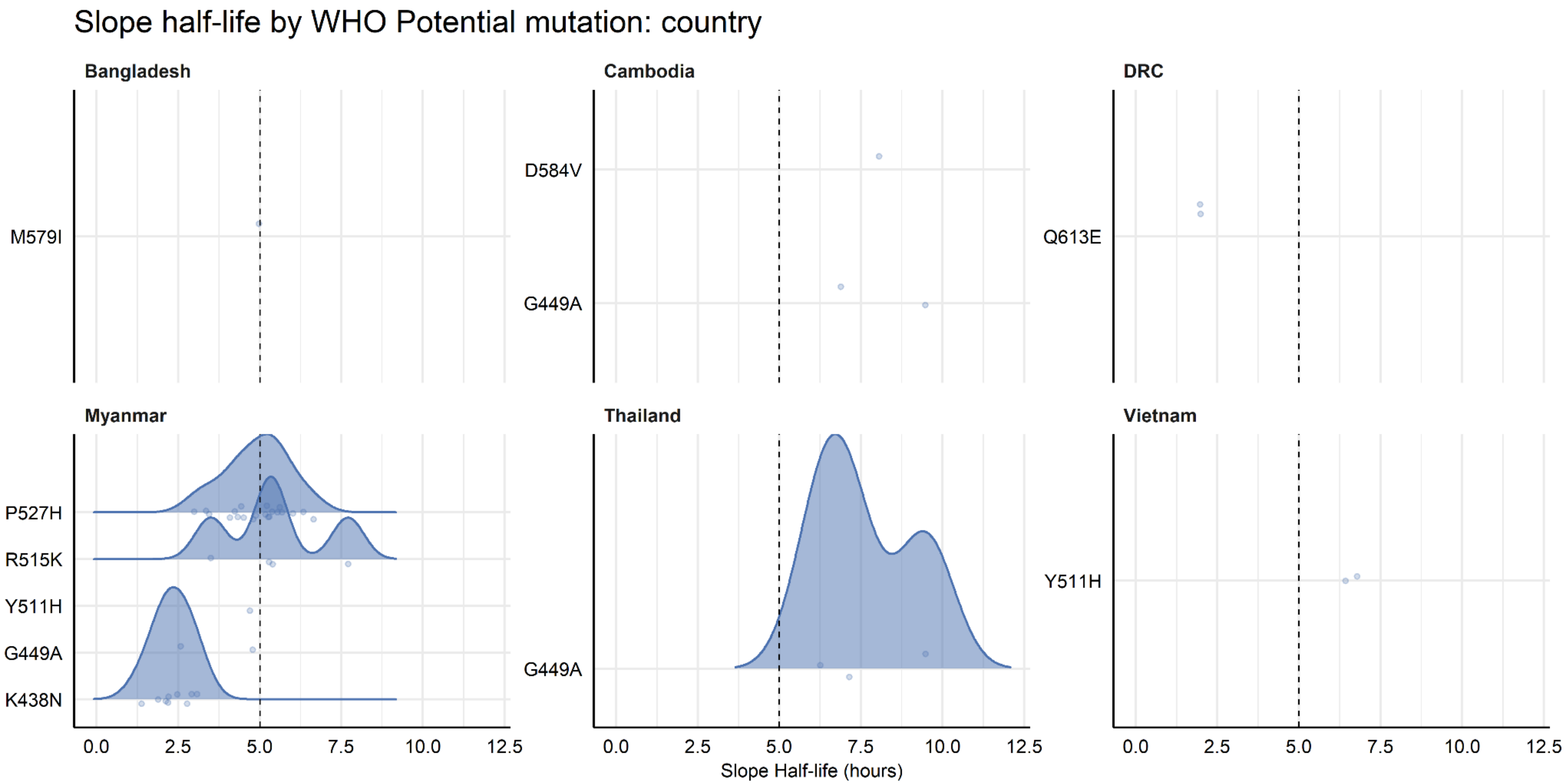

**S Figure 4.5:** Distribution of slope half-life for WHO potential mutations, by country.
Legend: Vertical dotted line shown at 5h.

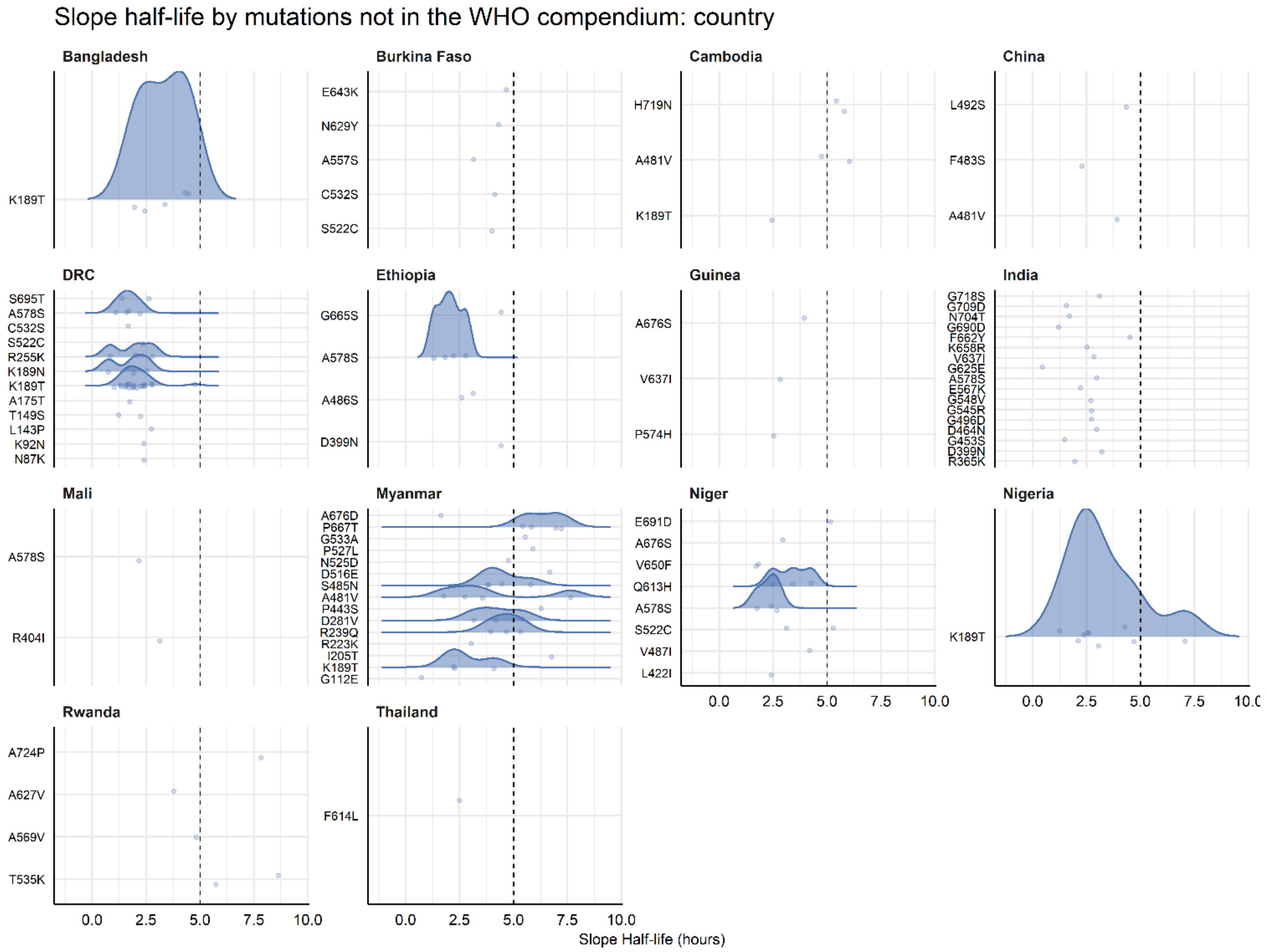

**S Figure 4.6**: Distribution of slope half-life for mutations not current in the 2025 WHO compendium of markers, by country. Legend: Vertical dotted line shown at 5h.

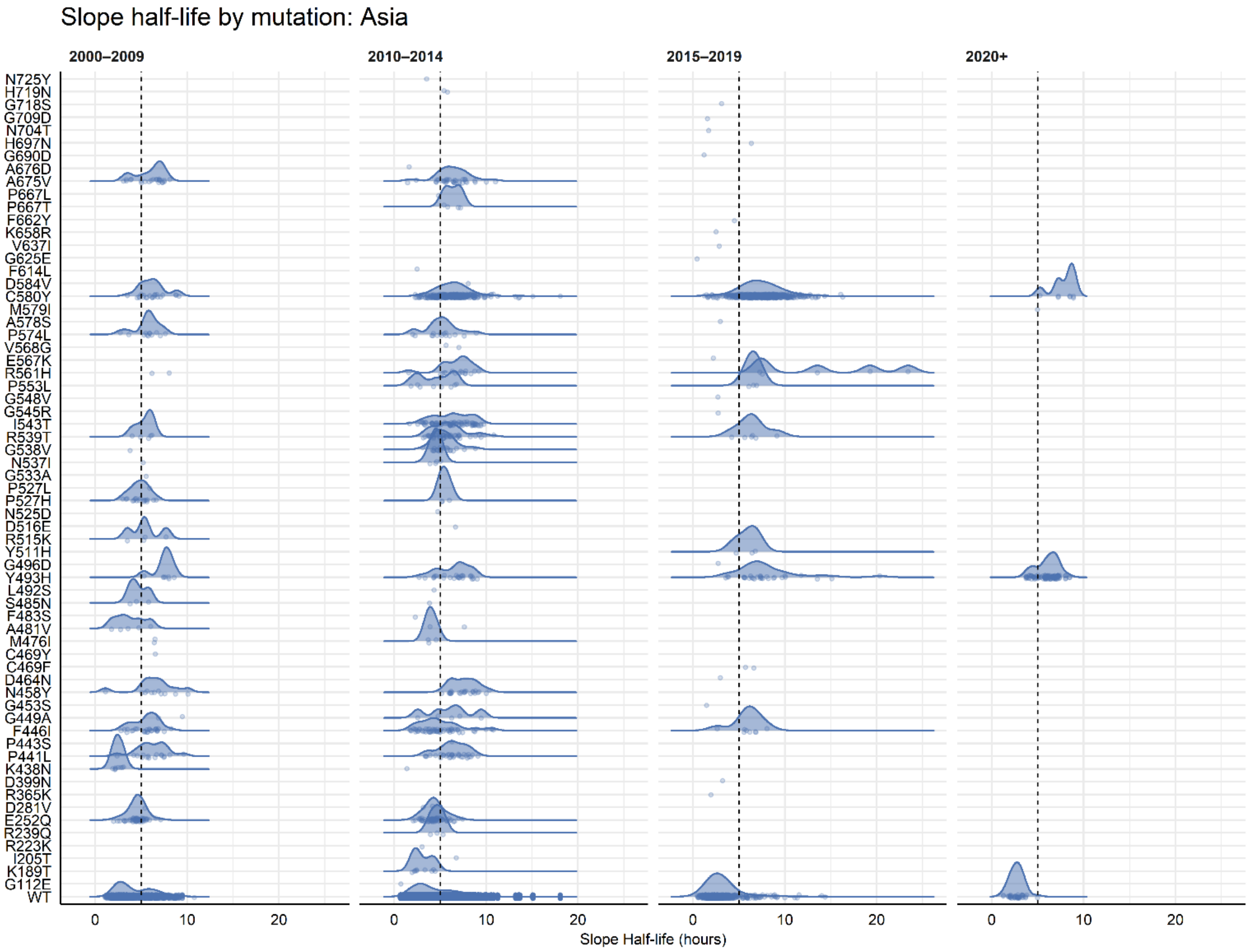

**S Figure 4.7**: Slope half-distribution by evolution of individual mutation over time period, Asia. Legend: Vertical dotted line shown at 5h.

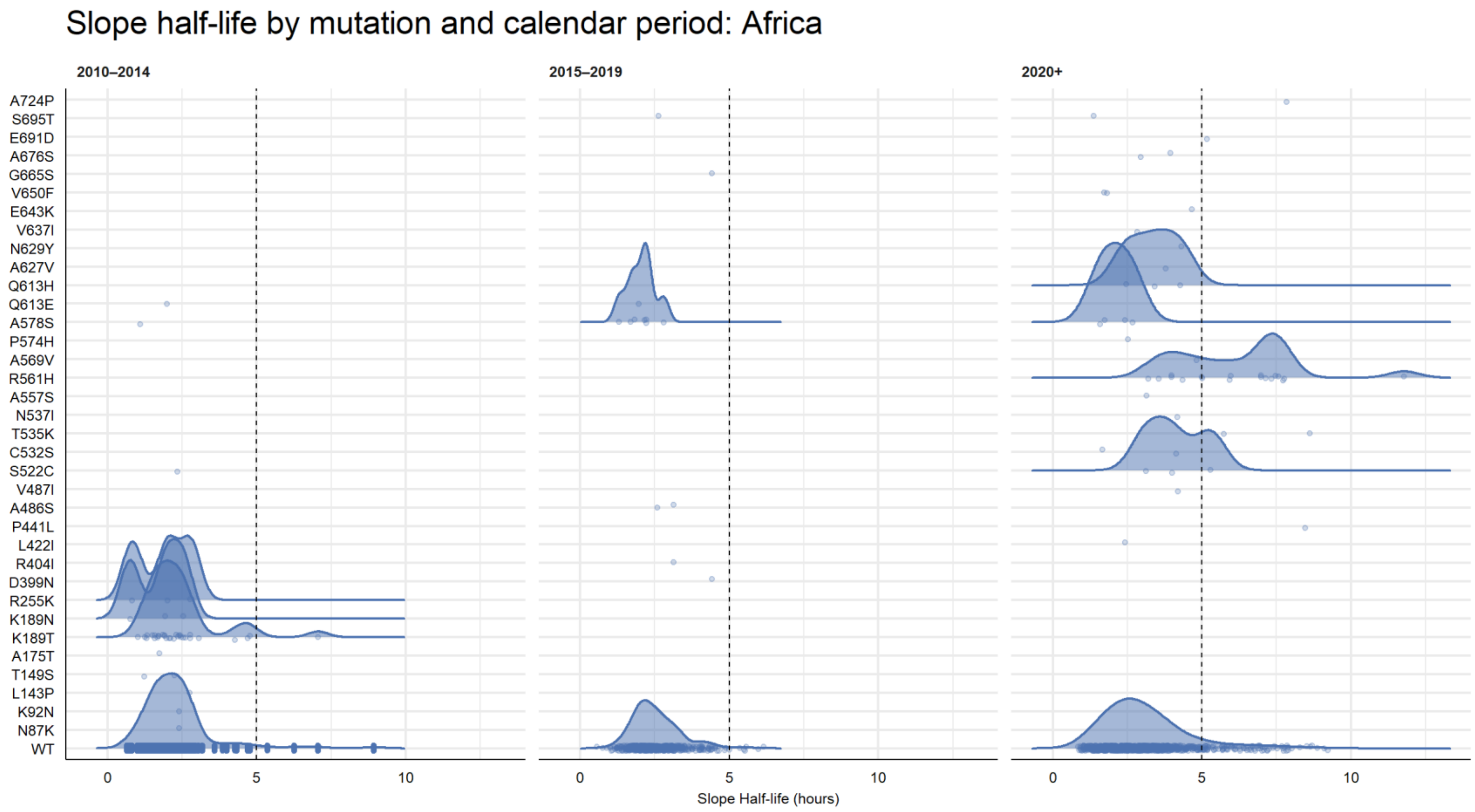

**S Figure 4.8**: Slope half-distribution by evolution of individual mutation over time period, Africa.
Legend: Vertical dotted line shown at 5h.

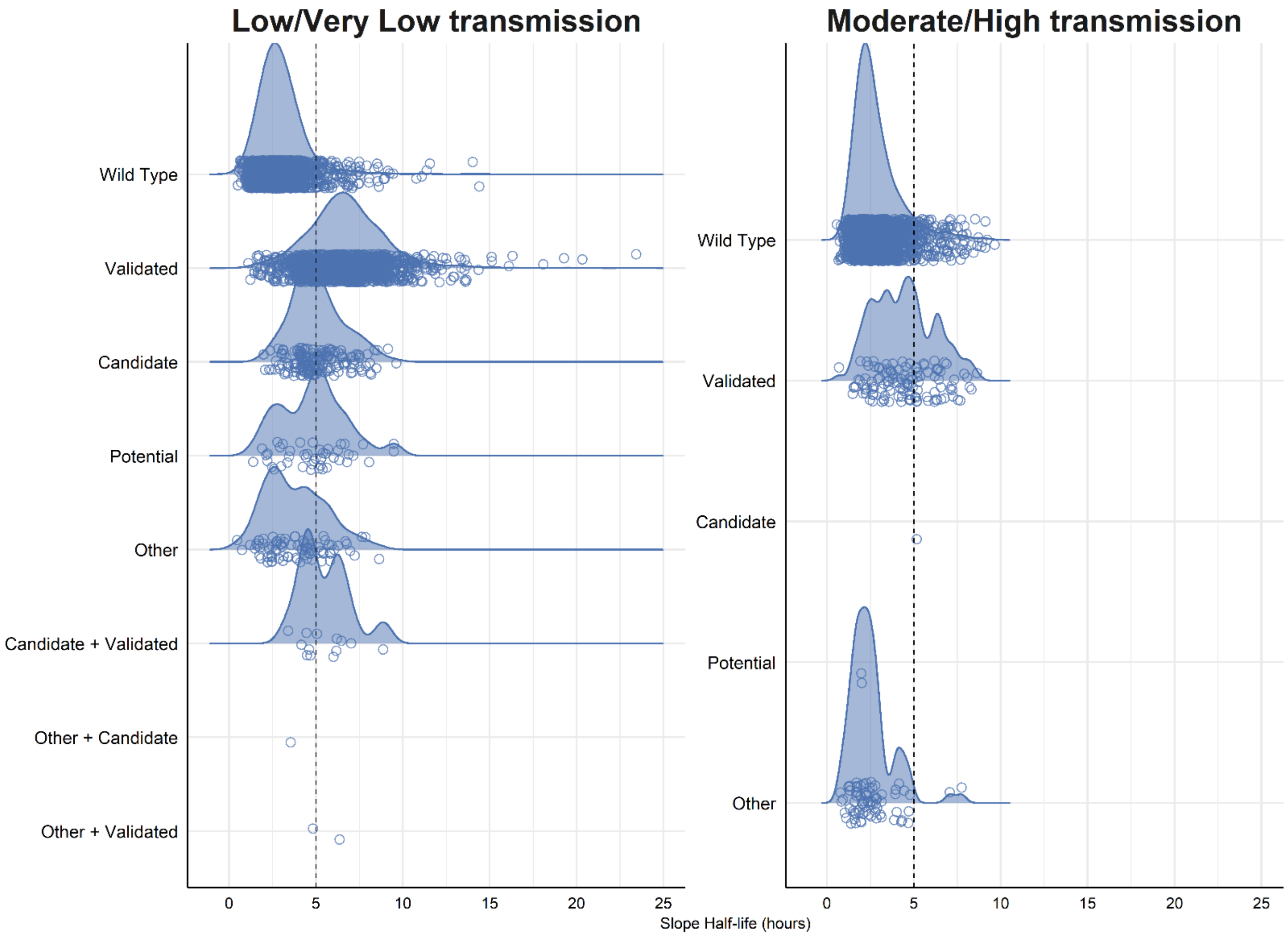

**S Figure 4.9**: Slope half-distribution by mutation category, by transmission groups.
Legend: Vertical dotted line shown at 5h. Validated = WHO validated, Candidate = WHO candidate, Potential = WHO potential mutations.

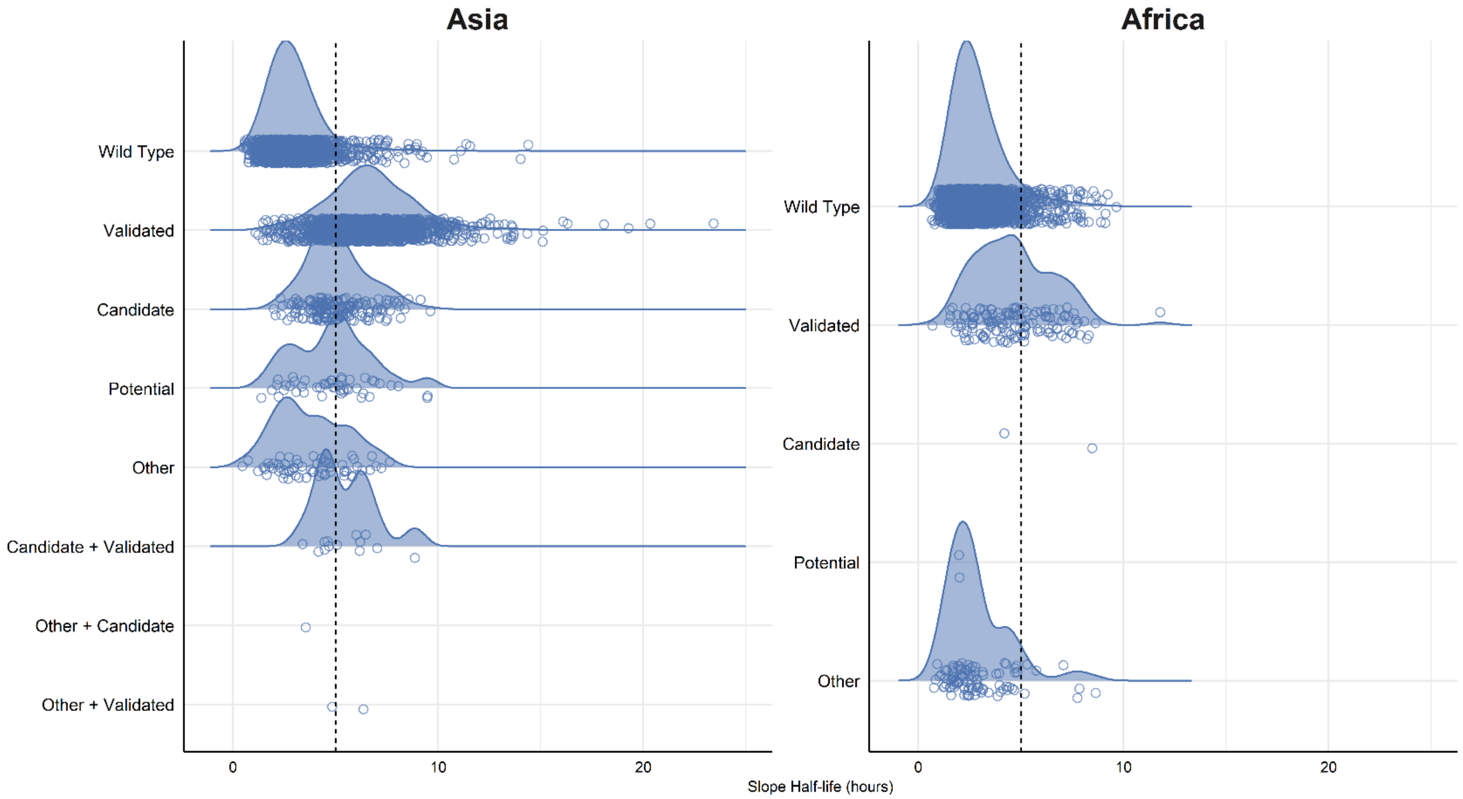

**S Figure 4.10**: Slope half-distribution by mutation category, by continent.
Legend: Vertical dotted line shown at 5h. Validated = WHO validated, Candidate = WHO candidate, Potential = WHO potential mutations.

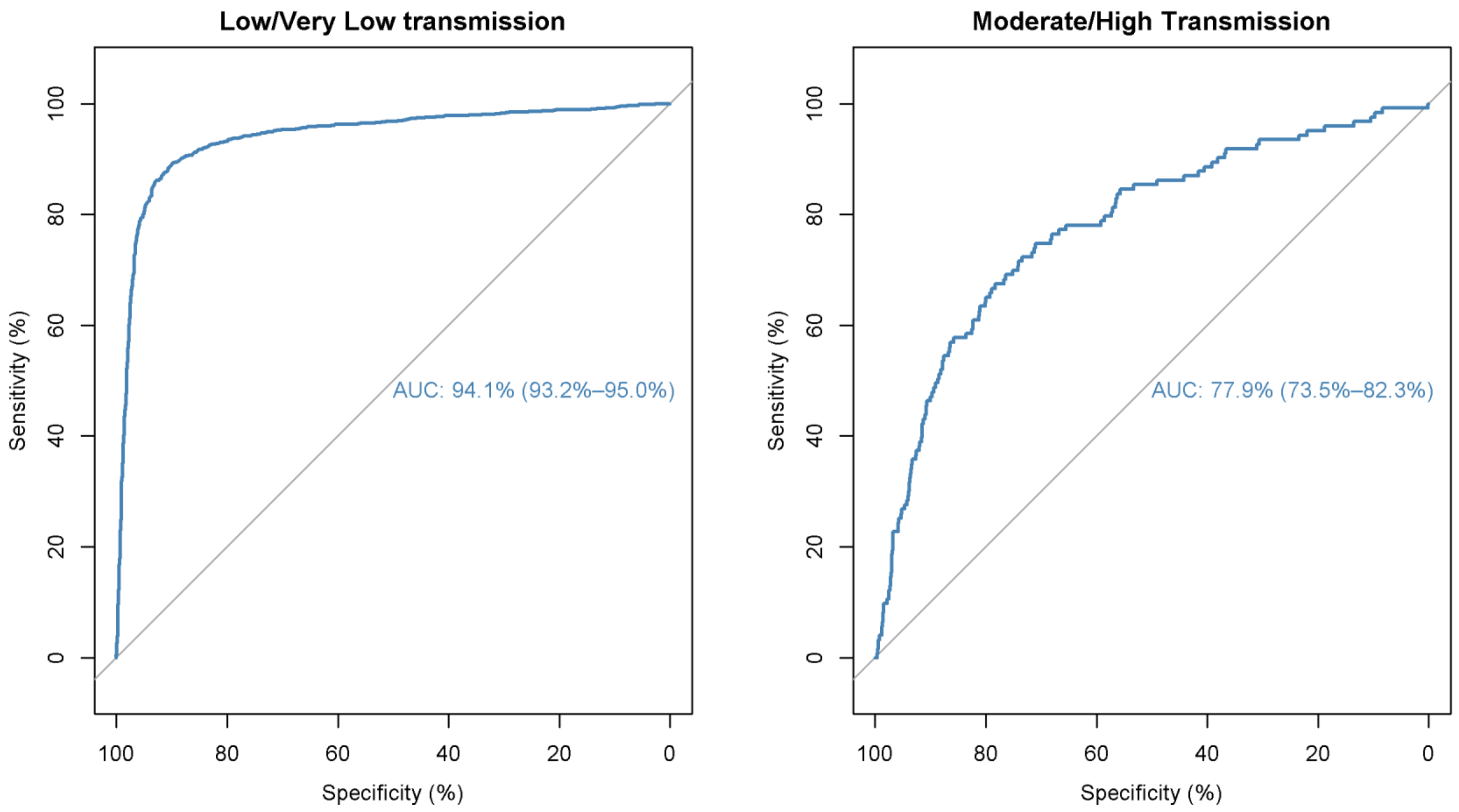

**S Figure 4.11**: ROC curves for diagnostic accuracy of slope half thresholds for identifying a WHO-validated mutation vs. WT, by transmission.

### Supplemental Tables

S **Table 4.1**: Parasite clearance slope half-life by regions

The overall median parasite clearance slope half-life (PC½) was 3.6 h [IQR: 2.5-6.0; n=4,604] in Asia and 2.7h [IQR: 2.0–3.6; n=2,953] in Africa. The PCE ½ was 18% faster in Africa than in Asia (geometric mean ratio 0.82; 95% CI, 0.71-0.95; study sites modelled as random effects). When stratified by transmission setting, the PC½ was similarly faster in the areas of moderate to high transmission compared to low or very low (GMR: 0.79, 95% CI: 0.68-0.93); corresponding PC½ was: 3.5h [IQR: 2.5–5.8; n=5,170] in areas of low/very low transmission and 2.5h [IQR: 2.0–3.5; n=2,389] in moderate/high transmission. The proportion of patients with PC½ >5h was 34.5% (1,589/4,604) in Asia compared to 9.3% (275/2,953) in Africa [odds ratio (OR); 7.8, 95% CI: 3.5-17.6, ref: Africa], 32.2% (1664/5170) in low/very low-transmission settings, and 8.4% (200/2389) in moderate/high-transmission settings [OR: 9.7, 95% CI: 3.9-24.0, ref: low/very low]. For all age groups except infants (<1y), the PC½ was shorter in areas of moderate-to-high transmission and in Africa, reflecting the role of immunity.

**S Table 4.1A:** By continent

| | **Africa** | | | **Asia** | | | **PC½ prolongation ratio for patients in Asia (reference: Africa)** | |
|---|---|---|---|---|---|---|---|---|
| | *n* | Median PC½ [IQR] | PC½ >5h | *n* | Median PC½ [IQR] | PC½ >5h | Ratio of median PC½ [a] | Geometric mean ratio [95% CI] [b] |
| <1y | 67 | 2.55 [1.93–3.46] | 9% (6/67) | 28 | 3.12 [2.51–4.83] | 25% (7/28) | 1.22 | 0.94 [0.59–1.51] |
| 1-4y | 1076 | 2.62 [1.98–3.61] | 10.5% (113/1076) | 331 | 3.16 [2.37–4.56] | 20.8% (69/331) | 1.21 | 1.27 [1.03–1.57] |
| 5-12y | 1682 | 2.68 [2.06–3.55] | 8.2% (138/1682) | 810 | 3.17 [2.32–4.65] | 22.1% (179/810) | 1.19 | 1.21 [1.01–1.46] |
| 13+y | 104 | 2.98 [2.30–4.17] | 17.3% (18/104) | 3435 | 3.89 [2.56–6.40] | 38.8% (1334/3435) | 1.31 | 1.35 [0.98–1.85] |
| NA | 24 | 1.94 [1.83–2.23] | 0% (0/24) | - | - | | - | - |
| Overall | 2953 | 2.65 [2.03–3.56] | 9.3% (275/2,953) | 4604 | 3.64 [2.50–6.04] | 34.5% (1,589/4,604) | 1.37 | 1.25 [1.09–1.44] |

PC½ = Parasite clearance slope half-life; *n* = Number of patients; IQR = Interquartile range

[a] Unadjusted ratio of median half-life for moderate-to-high transmission with respect to low-very low transmission.

[b] Geometric mean ratio obtained from a mixed effects linear regression with study sites as random effects and transmission setting as fixed effect

**S Table 4.1B:** By Transmission settings

| | Mod/High Transmission | | | Low/V Low Transmission | | | Comparison between transmission settings (reference: Low/Very Low) | |
|---|---|---|---|---|---|---|---|---|
| | *n* | Median PC½ [IQR] | PC½ >5h | *n* | Median PC½ [IQR] | PC½ >5h | Ratio of median PC½ [a] | Geometric mean ratio [95% CI] [b] |
| <1y | 63 | 2.42 [1.87–3.52] | 9.5% (6/63) | 32 | 3.27 [2.55–4.57] | 22% (7/32) | 0.74 | 1.04 [0.67-1.61] |
| 1–4y | 883 | 2.47 [1.91–3.42] | 8.7% (77/883) | 524 | 3.17 [2.51–4.52] | 20% (105/524) | 0.78 | **0.78 [0.64-0.95]** |
| 5–12y | 1,346 | 2.58 [2.01–3.47] | 8.3% (112/1346) | 1,146 | 3.10 [2.38–4.26] | 17.9% (205/1146) | 0.83 | **0.77 [0.63-0.93]** |
| >12y | 73 | 2.61 [2.11–3.40] | 6.8% (5/73) | 3,468 | 3.90 [2.56–6.41] | 38.8% (1347/3468) | 0.67 | **0.65 [0.46-0.92]** |
| Missing | 24 | 1.94 [1.83–2.23] | 0% (0/24) | - | - | - | - | - |
| Overall | 2,389 | 2.53[1.96–3.44] | 8.4% (200/2389) | 5,170 | 3.53 [2.50–5.81] | 32.2% (1664/5170) | 0.72 | **0.77 [0.66–0.90]** |

PC½ = Parasite clearance slope half-life; *n* = Number of patients; IQR = Interquartile range. Bold indicates statistically significant comparison between the transmission settings.

[a] Unadjusted ratio of median half-life for moderate-to-high transmission with respect to low and very low transmission.

[b] Geometric mean ratio obtained from mixed effects linear regression on log transformed slope half-life with study sites as random effects, and transmission setting as the only fixed effect.

**S Table 4.2**: Distribution of Slope half-life (PC½) by country

| Country | Continent | Number of studies | Number of patients | Geometric mean PC½ | Median PC½ /h | 25th | 75th | 95th | Min | Max |
|---|---|---|---|---|---|---|---|---|---|---|
| **Asia** | | | | | | | | | | |
| Bangladesh | Asia | 3 | 183 | 2.87 | 2.95 | 2.40 | 3.59 | 4.51 | 0.56 | 6.20 |
| Cambodia | Asia | 7 | 1,243 | 4.84 | 5.54 | 3.24 | 7.28 | 9.80 | 0.67 | 20.33 |
| China | Asia | 1 | 105 | 3.85 | 3.61 | 2.61 | 5.61 | 8.67 | 1.53 | 12.28 |
| India | Asia | 2 | 251 | 2.19 | 2.25 | 1.72 | 2.90 | 4.11 | 0.45 | 5.80 |
| Laos | Asia | 3 | 85 | 2.07 | 1.92 | 1.61 | 2.27 | 3.94 | 1.13 | 7.31 |
| Myanmar | Asia | 5 | 1,426 | 3.38 | 3.24 | 2.42 | 4.83 | 7.49 | 0.49 | 23.43 |
| Thailand | Asia | 4 | 770 | 3.59 | 3.40 | 2.50 | 5.18 | 8.52 | 0.90 | 15.10 |
| Vietnam | Asia | 6 | 541 | 4.61 | 5.60 | 2.98 | 7.29 | 9.21 | 0.72 | 12.11 |
| **Africa** | | | | | | | | | | |
| Burkina Faso | Africa | 1 | 254 | 4.03 | 4.02 | 3.05 | 5.19 | 7.76 | 1.35 | 9.22 |
| DRC | Africa | 3 | 392 | 2.08 | 2.16 | 1.72 | 2.55 | 3.27 | 0.65 | 6.26 |
| Ethiopia | Africa | 1 | 193 | 2.55 | 2.59 | 2.02 | 3.34 | 4.42 | 0.56 | 5.97 |
| Gambia | Africa | 1 | 62 | 2.98 | 3.09 | 2.43 | 3.68 | 4.91 | 1.28 | 5.92 |
| Ghana | Africa | 1 | 3 | 5.54 | 4.70 | 4.64 | 6.31 | 7.60 | 4.57 | 7.92 |
| Guinea | Africa | 1 | 131 | 2.43 | 2.46 | 1.99 | 3.05 | 3.99 | 0.85 | 7.26 |
| Kenya | Africa | 1 | 45 | 2.71 | 2.94 | 2.25 | 3.47 | 4.45 | 0.88 | 5.00 |
| Mali | Africa | 1 | 267 | 2.42 | 2.40 | 2.01 | 2.93 | 3.86 | 1.24 | 6.16 |
| Niger | Africa | 1 | 333 | 2.93 | 2.94 | 2.50 | 3.58 | 4.52 | 1.07 | 6.47 |
| Nigeria | Africa | 2 | 181 | 1.94 | 1.78 | 1.37 | 2.56 | 4.12 | 0.85 | 17.88 |
| Rwanda | Africa | 1 | 31 | 5.28 | 5.74 | 3.98 | 7.40 | 8.55 | 2.00 | 11.77 |
| Tanzania | Africa | 2 | 172 | 3.71 | 3.56 | 2.57 | 5.31 | 8.98 | 1.10 | 18.45 |
| Uganda | Africa | 2 | 889 | 2.81 | 2.69 | 2.05 | 3.87 | 6.34 | 0.68 | 12.10 |
| **Missing** | 0 | 1 | 2 | 2.22 | 2.35 | 1.96 | 2.74 | 3.05 | 1.57 | 3.13 |

PC½ = Parasite clearance slope half-life reported in hours; DRC = Democratic Republic of Congo

**S Table 4.3:** Distribution of Slope half-life (PC½) by country, above the given half-life threshold

| | Number of patients | PC½ >3h | PC½>3.5h | PC½>4h | PC½>4.5 | PC½>5h | PC½>5.5h |
|---|---|---|---|---|---|---|---|
| **Asia** | | | | | | | |
| Bangladesh | 183 | 48.1% (88/183) | 28.4% (52/183) | 14.8% (27/183) | 5.5% (10/183) | 2.7% (5/183) | 0.5% (1/183) |
| Cambodia | 1,243 | 79.2% (985/1243) | 71.2% (885/1243) | 65.8% (818/1243) | 60.8% (756/1243) | 55.9% (695/1243) | 50.6% (629/1243) |
| China | 105 | 68.6% (72/105) | 52.4% (55/105) | 42.9% (45/105) | 36.2% (38/105) | 33.3% (35/105) | 25.7% (27/105) |
| India | 251 | 21.9% (55/251) | 12.4% (31/251) | 6% (15/251) | 2.8% (7/251) | 1.2% (3/251) | 0.4% (1/251) |
| Laos | 85 | 14.1% (12/85) | 8.2% (7/85) | 4.7% (4/85) | 4.7% (4/85) | 4.7% (4/85) | 4.7% (4/85) |
| Myanmar | 1,426 | 56.2% (801/1426) | 44.4% (633/1426) | 36.5% (520/1426) | 28.5% (406/1426) | 23.5% (335/1426) | 18.2% (260/1426) |
| Thailand | 770 | 60% (462/770) | 47.1% (363/770) | 38.1% (293/770) | 32.1% (247/770) | 26.9% (207/770) | 22.9% (176/770) |
| Vietnam | 541 | 74.9% (405/541) | 68.4% (370/541) | 64.3% (348/541) | 61.2% (331/541) | 56.4% (305/541) | 50.5% (273/541) |
| **Africa** | | | | | | | |
| Burkina Faso | 254 | 76.4% (194/254) | 65% (165/254) | 50.4% (128/254) | 39.8% (101/254) | 28.3% (72/254) | 22.4% (57/254) |
| DRC | 392 | 9.2% (36/392) | 3.6% (14/392) | 2% (8/392) | 0.8% (3/392) | 0.3% (1/392) | 0.3% (1/392) |
| Ethiopia | 193 | 36.3% (70/193) | 19.2% (37/193) | 12.4% (24/193) | 4.7% (9/193) | 3.1% (6/193) | 1.6% (3/193) |
| Gambia | 62 | 53.2% (33/62) | 33.9% (21/62) | 14.5% (9/62) | 8.1% (5/62) | 3.2% (2/62) | 1.6% (1/62) |
| Ghana | 3 | 100% (3/3) | 100% (3/3) | 100% (3/3) | 100% (3/3) | 33.3% (1/3) | 33.3% (1/3) |
| Guinea | 131 | 26.7% (35/131) | 14.5% (19/131) | 5.3% (7/131) | 3.1% (4/131) | 3.1% (4/131) | 2.3% (3/131) |
| Kenya | 45 | 48.9% (22/45) | 24.4% (11/45) | 8.9% (4/45) | 4.4% (2/45) | 2.2% (1/45) | 0% (0/45) |
| Mali | 267 | 20.6% (55/267) | 7.9% (21/267) | 4.9% (13/267) | 1.5% (4/267) | 0.4% (1/267) | 0.4% (1/267) |
| Niger | 333 | 46.8% (156/333) | 27.9% (93/333) | 12.9% (43/333) | 5.4% (18/333) | 2.7% (9/333) | 0.6% (2/333) |
| Nigeria | 181 | 16% (29/181) | 9.9% (18/181) | 6.1% (11/181) | 4.4% (8/181) | 3.3% (6/181) | 2.8% (5/181) |
| Rwanda | 31 | 90.3% (28/31) | 83.9% (26/31) | 71% (22/31) | 64.5% (20/31) | 58.1% (18/31) | 54.8% (17/31) |
| Tanzania | 172 | 61.6% (106/172) | 52.9% (91/172) | 41.9% (72/172) | 35.5% (61/172) | 29.1% (50/172) | 23.8% (41/172) |
| Uganda | 889 | 41.8% (372/889) | 31.2% (277/889) | 22.9% (204/889) | 16.5% (147/889) | 11.7% (104/889) | 7.9% (70/889) |

PC½ = Parasite clearance slope half-life; DRC = Democratic Republic of Congo

S Table 4.4: Additional mutations observed in areas of moderate/high transmissions which were not listed in the WHO 2025 compendium

| Mutation | Number of patients | Median PC½ | PC½ >5h |
|---|---|---|---|
| N87K | 1 | 2.4 [2.4–2.4] | 0% (0/1) |
| K92N | 1 | 2.4 [2.4–2.4] | 0% (0/1) |
| L143P | 1 | 2.7 [2.7–2.7] | 0% (0/1) |
| T149S | 2 | 1.7 [1.5–2] | 0% (0/2) |
| A175T | 1 | 1.7 [1.7–1.7] | 0% (0/1) |
| K189N | 3 | 1.9 [1.3–2.2] | 0% (0/3) |
| **K189T** | **31** | **2.1 [1.7–2.6]** | **3.2% (1/31)** |
| R255K | 3 | 2 [1.4–2.4] | 0% (0/3) |
| D399N | 1 | 4.4 [4.4–4.4] | 0% (0/1) |
| R404I | 1 | 3.1 [3.1–3.1] | 0% (0/1) |
| A486S | 2 | 2.9 [2.7–3] | 0% (0/2) |
| S522C | 2 | 3.2 [2.8–3.6] | 0% (0/2) |
| C532S | 2 | 2.9 [2.3–3.5] | 0% (0/2) |
| A557S | 1 | 3.1 [3.1–3.1] | 0% (0/1) |
| **A569S** | **2** | **5.3 [4.1–6.5]** | **50% (1/2)** |
| P574H | 1 | 2.5 [2.5–2.5] | 0% (0/1) |
| A578S | 15 | 2.2 [1.7–2.8] | 0% (0/15) |
| N629Y | 1 | 4.3 [4.3–4.3] | 0% (0/1) |
| V637I | 1 | 2.8 [2.8–2.8] | 0% (0/1) |
| E643K | 1 | 4.7 [4.7–4.7] | 0% (0/1) |
| G665S | 1 | 4.4 [4.4–4.4] | 0% (0/1) |
| A676S | 1 | 3.9 [3.9–3.9] | 0% (0/1) |
| T685P | 1 | 2.1 [2.1–2.1] | 0% (0/1) |
| S695T | 3 | 1.4 [1.1–2] | 0% (0/3) |
| L708I | 1 | 1.6 [1.6–1.6] | 0% (0/1) |

[a] Bold indicates at least one patient had a PC½ >5h for the given mutation

S Table 4.5: Additional mutations observed in areas of low/very low transmissions which were not listed in the WHO 2025 compendium

| **Mutation** | **Number of patients** | **Median PC½** | **PC½ >5h** |
|---|---|---|---|
| G112E | 1 | 0.7 [0.7–0.7] | 0% (0/1) |
| K189T | 9 | 2.5 [2.3–4.1] | 0% (0/9) |
| **I205T** | **1** | **6.8 [6.8–6.8]** | **100% (1/1)** |
| R223K | 1 | 3 [3–3] | 0% (0/1) |
| **R239Q** | **3** | **4.7 [4.3–5]** | **33.3% (1/3)** |
| **D281V** | **3** | **4.2 [3.7–4.8]** | **33.3% (1/3)** |
| R365K | 1 | 1.9 [1.9–1.9] | 0% (0/1) |
| D399N | 1 | 3.2 [3.2–3.2] | 0% (0/1) |
| L422I | 1 | 2.4 [2.4–2.4] | 0% (0/1) |
| **P443S** | **1** | **6.3 [6.3–6.3]** | **100% (1/1)** |
| G453S | 1 | 1.5 [1.5–1.5] | 0% (0/1) |
| D464N | 1 | 3 [3–3] | 0% (0/1) |
| **A481V** | **7** | **3.9 [3.2–5.4]** | **28.6% (2/7)** |
| F483S | 1 | 2.3 [2.3–2.3] | 0% (0/1) |
| **S485N** | **4** | **4.2 [3.8–4.8]** | **25% (1/4)** |
| V487I | 1 | 4.2 [4.2–4.2] | 0% (0/1) |
| L492S | 1 | 4.3 [4.3–4.3] | 0% (0/1) |
| G496D | 1 | 2.7 [2.7–2.7] | 0% (0/1) |
| **D516E** | **1** | **6.7 [6.7–6.7]** | **100% (1/1)** |
| W518L | 1 | 4.6 [4.6–4.6] | 0% (0/1) |
| S522C | 2 | 4.2 [3.7–4.7] | 50% (1/2) |
| N525D | 1 | 4.8 [4.8–4.8] | 0% (0/1) |
| **P527L** | **1** | **5.9 [5.9–5.9]** | **100% (1/1)** |
| **G533A** | **1** | **5.5 [5.5–5.5]** | **100% (1/1)** |

| Mutation | Number of patients | Median PC½ | PC½ >5h |
|---|---|---|---|
| **T535K** | **2** | **7.2 [6.5–7.9]** | **100% (2/2)** |
| G545R | 1 | 2.7 [2.7–2.7] | 0% (0/1) |
| G548V | 1 | 2.7 [2.7–2.7] | 0% (0/1) |
| E567K | 1 | 2.2 [2.2–2.2] | 0% (0/1) |
| A569V | 1 | 4.8 [4.8–4.8] | 0% (0/1) |
| A578S | 5 | 2.7 [2.4–3] | 0% (0/5) |
| Q613H | 3 | 3.4 [2.9–3.8] | 0% (0/3) |
| F614L | 1 | 2.5 [2.5–2.5] | 0% (0/1) |
| G625E | 1 | 0.5 [0.5–0.5] | 0% (0/1) |
| A627V | 1 | 3.8 [3.8–3.8] | 0% (0/1) |
| V637I | 1 | 2.8 [2.8–2.8] | 0% (0/1) |
| V650F | 2 | 1.8 [1.7–1.8] | 0% (0/2) |
| K658R | 1 | 2.5 [2.5–2.5] | 0% (0/1) |
| F662Y | 1 | 4.5 [4.5–4.5] | 0% (0/1) |
| P667L | 1 | 4.8 [4.8–4.8] | 0% (0/1) |
| **P667T** | **4** | **6.4 [5.7–7]** | **100% (4/4)** |
| A676D | 1 | 1.6 [1.6–1.6] | 0% (0/1) |
| A676S | 1 | 2.9 [2.9–2.9] | 0% (0/1) |
| G690D | 1 | 1.2 [1.2–1.2] | 0% (0/1) |
| **E691D** | **1** | **5.2 [5.2–5.2]** | **100% (1/1)** |
| **H697N** | **1** | **6.4 [6.4–6.4]** | **100% (1/1)** |
| N704T | 1 | 1.7 [1.7–1.7] | 0% (0/1) |
| G709D | 1 | 1.6 [1.6–1.6] | 0% (0/1) |
| G718S | 1 | 3.1 [3.1–3.1] | 0% (0/1) |
| **H719N** | **2** | **5.6 [5.5–5.7]** | **100% (2/2)** |
| **A724P** | **1** | **7.8 [7.8–7.8]** | **100% (1/1)** |
| N725Y | 1 | 3.5 [3.5–3.5] | 0% (0/1) |

[a] Bold indicates at least one patient had a PC½ >5h for the given mutation

**S Table 4.6**: Diagnostic accuracy of different slope half-life in identifying WHO validated mutations, by transmission settings

| Transmission settings | PC½ (/h) | TP | FP | TN | FN | Total patients | Sensitivity [95% CI] | Specificity [95% CI] |
|---|---|---|---|---|---|---|---|---|
| **Low/very Low settings** | | | | | | | | |
| | 3 | 1,275 | 954 | 1,379 | 50 | 3,658 | 96.2% (95.1 - 97.1) | 59.1% (57.1 - 61.1) |
| | 3.5 | 1,248 | 561 | 1,772 | 77 | 3,658 | 94.2% (92.8 - 95.3) | 76% (74.2 - 77.6) |
| | 4 | 1,202 | 315 | 2,018 | 123 | 3,658 | 90.7% (89 - 92.2) | 86.5% (85.1 - 87.8) |
| | 4.5 | 1,142 | 175 | 2,158 | 183 | 3,658 | 86.2% (84.2 - 87.9) | 92.5% (91.4 - 93.5) |
| | 5 | 1,070 | 120 | 2,213 | 255 | 3,658 | 80.8% (78.5 - 82.8) | 94.9% (93.9 - 95.7) |
| | 5.5 | 966 | 78 | 2,255 | 359 | 3,658 | 72.9% (70.4 - 75.2) | 96.7% (95.8 - 97.3) |
| **Moderate/High settings** | | | | | | | | |
| | 3 | 94 | 638 | 1,354 | 29 | 2,115 | 76.4% (68.2 - 83.1) | 68% (65.9 - 70) |
| | 3.5 | 82 | 430 | 1,562 | 41 | 2,115 | 66.7% (57.9 - 74.4) | 78.4% (76.6 - 80.2) |
| | 4 | 71 | 293 | 1,699 | 52 | 2,115 | 57.7% (48.9 - 66.1) | 85.3% (83.7 - 86.8) |
| | 4.5 | 58 | 200 | 1,792 | 65 | 2,115 | 47.2% (38.6 - 55.9) | 90% (88.6 - 91.2) |
| | 5 | 44 | 138 | 1,854 | 79 | 2,115 | 35.8% (27.9 - 44.6) | 93.1% (91.9 - 94.1) |
| | 5.5 | 33 | 97 | 1,895 | 90 | 2,115 | 26.8% (19.8 - 35.3) | 95.1% (94.1 - 96) |

PC½ = Parasite clearance slope half-life; 95% CI: 95% confidence interval; TP = True positive, FP = False positive, TN = True Negative, FN = False Negative; 95% CI estimated using all data for the respective transmission settings and ignores clustering.

**Supplemental file for PCE1/2: Risk of availability bias due to unavailable IPD in studies with parasite clearance slope half-life (PC½) measured**

Table S5.1: Risk of bias in the studies based on profile exclusion

For each individual study, risk was classified as low L, moderate M, or high H using the following rules: PC1/2 exclusions due to sparse data or unsatisfactory model fit: Low < 20% of patients excluded, Moderate if 20% -25% patients excluded, High ≥25% patients excluded.

| **Study ID** | **Sampling Design** | **Parasite-time profiles** | | | | |
|---|---|---|---|---|---|---|
| | | **Total patients** | **Low R-squared (<80%)** | **Profiles with lag≥12h detected** | **Unsatisfactory fit profiles** | **Study Classification** |
| 38352311 | 0, 8, 16, 24, 32, …. | 172 | 4 | 14 | 10·5% | L |
| 23101492 | 6h until negative | 166 | 4 | 13 | 10·2% | L |
| 33163636 | 8h until negative | 62 | 37 | 10 | 75·8% | H |
| - | 0, 4, 8, 12, 18,24, 30, 36h, … | 40 | 1 | 2 | 7·5% | L |
| 23175556 | 0,2,4,6,8,12, then 6h until negative | 79 | 2 | 5 | 8·9% | L |
| 32171078, 31345710 | 0,4,6,8,12, then 6h until negative | 1095 | 21 | 62 | 7·6% | L |
| 32320811 | 8h until negative | 406 | 75 | 2 | 19·0% | L/M |
| 34111412 | 0,4,6,8,12, then 6h until negative | 217 | 16 | 2 | 8·3% | L |
| 25224002 | 12h until negative | 89 | 11 | 5 | 18·0% | L/M |
| 35276064 | 0,4,6,8,12, then 6h until negative | 302 | 5 | 25 | 9·9% | L |
| - | 6h until negative | 2672 | 234 | 113 | 13·0% | L |
| 32437557 | 12h until negative | 58 | 0 | 9 | 15·5% | L |
| 27036739 | 6h until negative | 114 | 3 | 9 | 10·5% | L |
| 22403308 | 6h until negative | 43 | 3 | 2 | 11·6% | L |
| 28934435 | 6h until negative | 1836 | 69 | 101 | 9·3% | L |
| 29178921 | 0,4,6,8,12, then 6h until negative | 36 | 0 | 1 | 2·8% | L |
| 25877962 | 6h until negative | 107 | 1 | 3 | 3·7% | L |
| 26774243 | 0,2,4,6,8,12, then 6h until negative | 202 | 29 | 5 | 16·8% | L |
| 25075834 | 6h until negative | 1177 | 26 | 49 | 6·4% | L |
| 25910630 | 12h until negative | 139 | 11 | 7 | 12·9% | L |
| 40845863 | 6h until negative | 808 | 50 | 77 | 15·7% | L |
| 34551228, 36519341 | 0,4,6, 12, 18h, | 240 | 4 | 18 | 9·2% | L |

L = Low; M= Moderate; H = High

S Table 5.2: Assessment of bias due to studies not available for IPD-MA from Asia (n=24)

| PubMed ID (Registration) | Study period | Country | Enrolled | Sampling frequency | Tested for K13 | Key K13 findings | Likely impact on IPD-MA | Justification for impact assessment |
|---|---|---|---|---|---|---|---|---|
| 31660398 | ~2015 (deduced from text) | Cambodia | 205 | 4 and 8 hours after the first dose, then every 8 hours until 2 consecutive negatives | - Entire *Kelch13* propeller domain | All treatment failures were C580Y positive, with C580Y parasites cleared more slowly than non-C580Y parasites | L | C580Y is well represented in the IPD-MA |
| 31345710 | 2015-2018 | Thailand, Cambodia, Vietnam | 292 | PC1/2 assessed in 133 patients | 137 | *Kelch13* C580T mutations have increased substantially. 109 (82%) of 133 had a PC½ of more than 5·5 h.<br><br>*Kelch13* mutation prevalence: 124/137 (91%; 84·3–94·9) | L/M | Thai-Myanmar/Vietnam is well represented in the IPD-MA |
| 31833468 | 2014-2015 | Myanmar | 359 | - | 292 | 46.2% (135/292) with F446I the most common.<br><br>A676D reported in ~5%. All 3-recrudescence carried the F446I mutation. | M | |
| 26324266 | 2008-2013 | Myanmar, China-Myanmar border | 248 | Only assessed Day3 positivity (no evidence of PCE 1/2 data) | 57 | F446I reported | L | Only assessed the association with Day 3. Not applicable for PCE 1/2 assessment. |
| NCT02325180 | 2015 | Indonesia | 302 | - | - | Day 3, positive carried WT (wild-type *Kelch13*), but reported in another paper. | L | Only assessed the association with Day 3. Not applicable for PCE 1/2 assessment. |
| 35073756 | 2018-2019 | Bangladesh | 41 | Planned: 6, 12, 24, 36, and 48 h.<br>Field: every 6 h after administration of the first ACT | 41 | No correlation reported between *Kelch13* mutations and parasite clearance | L | Study reported no correlation reported between *Kelch13* mutations and parasite clearance |

| | | | | | | | | |
|---|---|---|---|---|---|---|---|---|
| | | | | (namely, at 6, 12, 18, and 24 h), followed by a time point collection every 12 h at 36, 48, 60, and 72 h | | | | |
| 32459308 | 2016-2017 | Cambodia | 63 | - | 63 | All 63 patients carried the C580Y mutation | L | The mutation is well represented in the IPDMA |
| 28549390 | 2012-2014 | India | 237 | Only assessed Day3 positivity (no evidence of PCE 1/2 data) | 135 | Analysis of 135 samples revealed a mutation in the *Kelch13* gene, with a non-synonymous single mutation at codon M579T (1.5%) and double mutations at codon M579T & N657H in 37%. | L | Only assessed the association with Day 3. Not applicable for PCE 1/2 assessment. |
| 37816434 | 2014-2023 | China-Myanmar border | 226 | Every 12 to 24 hours for the first three days. | 171 | G533S is independently associated with a slope half-life ≥5 hours;<br>G533S, P574L, and N458Y were associated with parasitaemia by<br>Day 3 and overall treatment failure, respectively.<br><br>The predominant mutation was F446I. G533S, C447R, C447S, N458Y, C469Y, and A676D. | L/M | F446I is well-represented in the IPDMA. The study also reports several mutations not in the IPDMA |
| 32524960 | 2017-2019 | Myanmar | 196 | - | - | R561H, C580Y, F446I, P574L There was no relationship between *Kelch13* mutations and day-3 parasite positivity rate. | L | The study assessed the association with Day 3 status. Not applicable for PCE 1/2 assessment. |
| 26926629 | 2014-2015 | Cambodia | 123 | Daily parasite sampling | 121 | 96% (166/121) carried the C580Y mutation. All cases of recrudescence harboured P. | L | The study assessed the association with Day 3 |

| | | | | | | | | |
|---|---|---|---|---|---|---|---|---|
| | | | | | | falciparum C580Y mutant parasites. | | status. Not applicable for PCE 1/2 assessment. |
| 30602520 | 2017 | Cambodia | 121 | Manuscript only reports Day 3 clearance (no evidence of PCE 1/2 data) | - | All three recrudescences patients carried C580Y | L | The study assessed the association with Day 3 status. Not applicable for PCE 1/2 assessment |
| 32854686 | 2015 | Myanmar | 41 | Manuscript only reports Day 3 clearance (no evidence of PCE 1/2 data) | - | The *Kelch13* K189T mutation (10.0%) was found in Myanmar for the first time. *Kelch13* mutations associated with artemisinin resistance were not found. | L | The study assessed the association with Day 3 status. Not applicable for PCE 1/2 assessment |
| 30428283 | 2013-2014 | India | 136 | Every 6h | - Codon 440 | Increased parasite clearance half-lives (>5 hours) observed in 14% of the patients; the majority of these patients carried the *Kelch13* polymorphism. The study identified G625R as a potential novel mutation along with R539T | M | Study reports mutations not identified in the IPD-MA. |
| 37553646 | 2010-2021 | Indonesia | 142 | - | - | Absence of mutations associated with ART resistance was noted. Several polymorphisms, such as L396F, I526V, N537S, and M579I, were found. | M | Indonesia is not represented in the IPD-MA. But unclear if frequent sampling is done for PCE 1/2 |
| 37948402 | 2012-2016 | China-Myanmar border | 198 | PCE1/2 not done | 174 | 46.9% (99/174) had the *Kelch13* mutation. F446I (79.80%, 79/99), P574L (7.07%, 7/99), C469Y (2.02%, 2/99), C580Y (2.02%, 2/99), N458Y (2.02%, 2/99) | L | The study assessed the association with Day 3 status. Not applicable for PCE 1/2 assessment |

| | | | | | | | | |
|---|---|---|---|---|---|---|---|---|
| 26911145 | 2013-2014 | Myanmar | 250 | | 206 (Codon sequenced: 210–726) | 33.5% (69/206) had a mutation in the propeller region.<br>Identified: P441L, F446I, G449A,<br>P574L, R561H, and A675 V. | L | The study assessed the association with Day 3 status. Not applicable for PCE 1/2 assessment |
| 28454557 | 2001-2014 | Thailand | 194 | Unclear | 194 | C580Y mutation is significantly associated with slow parasite clearance. | L | Thailand is well represented in the IPDMA, as is the C580Y mutation. |
| 30535043 | 2014-2016 | India | 226 | - | - | 24 patients (10.6%) had PC ½ >5 hours with pfkelch13 polymorphism after 441 codons. Of 15 patients with day 3 positive status, 13 had the *Kelch13* G625R polymorphism, whereas 2 had the R539T polymorphism. | M | Study reports mutations not identified in the IPDMA. |
| 28903755 | 2014 | Cambodia | 92 | - | 92 | The *Kelch13* assay results are interpretable for 76 of the 92 samples. Wild type: 9 (12%), C580Y: 64 (84%), Y493H: 3 (4%).<br>*Kelch13* mutants (67/76: 88%), only 1 of the 92 patients remained with a blood smear<br>Positive for *Plasmodium falciparum* on Day 3. | L | The study assessed the association with Day 3 status. Not applicable for PCE 1/2 assessment. |
| 34570647 | 2018-2019 | Vietnam | 50 | Every 12 h until blood films were negative on two consecutive samples | -<br>Codon sequenced: 430–622. | high prevalence of parasites carrying the *Kelch13* C580Y mutation<br>(92%, 46/50) | L | C580Y data from the region is well-represented in the IPDMA |

| | | | | | | | | |
|---|---|---|---|---|---|---|---|---|
| 28388902 | 2011-2014 | Myanmar | 470 | Daily parasite sampling | 288 | 33.7% (97/288) carried a *Kelch13* mutation. The F446I mutation was the most common, accounting for 66.0% of the mutations found. Seven out of nine Day 3 positive patients were infected with *Kelch13* wild-type parasites. The remaining two had the P574L mutation. | L | The study assessed the association with Day 3 status. Not applicable for PCE 1/2 assessment |
| 27737665 | 2014-2015 | India | 402 | - | 186 (Codon sequenced: 450–680) | Propeller region of *Kelch13* assessed in 186. Non-synonymous mutations were found in three samples at codon M579T. Treatment responses were good in all three patients harbouring NS *Kelch13* mutations. | L | Not applicable for PCE 1/2 assessment. |
| 34020652 | 2017 | India | 376 | - | 308 (Codon sequenced: 450–680.) | Q613H reported in 1 | L | Not applicable for PCE 1/2 assessment. |

S Table 5.3: Assessment of bias due to studies not available for IPD-MA from Africa (n=15)

| PubMed ID (Registration) | Study period | Country | Enrolled | Sampling frequency | Tested for K13 | Key K13 findings | Likely impact on IPD-MA | Justification for impact assessment |
|---|---|---|---|---|---|---|---|---|
| 32747827 | 2012-2015 | Rwanda | 954 | - | 507 (successful) | R561H found in 7.4% (19/257). Reported mutations: M460I, **C469Y,** R513L, V555A, R561H, **P574L,** R575I, A578S, G592E, E605K, A626E, V637I, E651K and P667R. No significant association between *Kelch13* nonsynonymous mutations and delayed parasite clearance as assessed by Day 3 | L/M | The manuscript reports an assessment of the association between *Kelch13* and Day 3. Not applicable for PCE 1/2 assessment. |
| 28450175 | 2015-2016 | Senegal | 181 | - | | Three synonymous mutations were detected in *Kelch13* (D464D, C469C and R471R); no non-synonymous mutations were detected. | L/M | From a country not represented in the HL analysis |
| 33468147 | 2016-2017 | Burkina Faso | 720 | - | | No *Kelch13* mutation previously associated with artemisinin-resistance was observed. N629Y (n = 1) and V517I (n = 2) mutants were observed in day 0 isolates collected from patients treated with AL and classified as recrudescence | L/M | Only assessed the association with Day 3. Not applicable for PCE 1/2 assessment. |
| 34569928 | 2012-2016 | Ivory coast | 770 | During the first 3 days, every 6 h until two consecutive negative parasitaemi a | 247 | D559N, and V510M reported;<br>6 patients had HL >5hr and it was associated with treatment failure. Linkage between mutation and treatment response is not clear from the publication, as the primary focus of the manuscript was on sickle cell/abnormal haemoglobin | Unclear | |
| 33864801 | 2018 | Rwanda | 228 | - | 218 | R561H (13%, 28/218); P574L (1%, 2/218) present. R561H is associated with Day 3 positivity | L/M | Only assessed the association with Day 3. Not applicable for PCE 1/2 assessment. |
| NCT03452475 | 2021 | Kenya | 217 | Baseline, 4, 6, 8, and 12h, and thereafter | 211 | Of these, 203 (96%) did not carry any non-synonymous mutations. A578S (n=4).<br>D399N, A486S, C542R, and G665S | L/M | Limited data from Kenya in the IPDMA, but a generally low |

| | | | | | | | | |
|---|---|---|---|---|---|---|---|---|
| | | | | every 6 h until two consecutive negatives | | | | prevalence of validated mutation reported. |
| 33107096 | 2017 | Tanzania | 1000 | - | - | None of the mutations associated (or confirmed) with delayed parasite clearance phenotypes was observed. Two samples carried the **R561H** mutation. | L | Reported in IPDMA |
| NCT04829695 | 2021 (estimated) | Cameroon | 181 | - | - | - | N/A | No information |
| NCT02940756 (PMID: 34491220) | 2017-2018 | Democratic Republic of the Congo | 1356 | - | 263 (codons: 389–649) | S477Y (n=1) identified. Other mutations were synonymous. Most samples (96%) included were WT. | L/M | Not applicable for PCE 1/2 assessment. |
| 32580771 | 2016 | Guinea | 421 | - | - | (98%, 380/389) of D0 and all day of failure (100%, 22/22) samples were wild type for *Kelch13*. The study reported 9 *Kelch13* mutations not associated with ART-R. | L/M | Only assessed theassociation with Day 3. Not applicable for PCE 1/2 assessment. |
| 38570791 | 2021-2022 | Togo | 357 | - | 357 | 99.2% (354/357) carried the *Kelch13* WT allele. A578S (n=2), A557S (n=1) | L/M | Not applicable for PCE 1/2 assessment. |
| 37754284 | 2016-2019 | Eritrea | 818 | - | - | R622I (13%, 109/818) was detected at baseline, and patients had 6-fold odds of Day 3 positivity | M/H | Only assessed the association with Day 3. Not applicable for PCE 1/2 assessment. |
| 29458380 | 2015 | Angola | 469 | - | 469 | The majority of pre-treatment samples (99%, 466/469) and all late-treatment failure samples (100%, 50/50) were WT for *Kelch13*. Three of the pre-treatment samples (1%) carried the A578S mutation. All-day late treatment failure samples were WT for Kelch13, but two recrudescent infections from Zaire carried one synonymous mutation each: T535T, which was also present in the participant's pre-treatment sample, and P553P, which was not present at enrolment. | L | Not applicable for PCE 1/2 assessment. |

| | | | | | | | | |
|---|---|---|---|---|---|---|---|---|
| 36519341 | 2018-2019 | Uganda | 133 | - | - | C469Y (n=5) and A675V (n=18)<br>Those with C469Y and A675V mutation had delayed clearance compared to WT; but overlapping distribution against WT | L/M | Uganda is not represented in the IPDMA |
| 36451216 | 2017 | Ethiopia | 181 | - | 50<br>(Codon sequenced: 389–649) | 9 haplotypes were assessed<br>(N456Y, Y493H, R539T, I543T, C580Y, F446I, M476I,<br>R561H, P553L), and no resistance-related mutants were observed. One *Kelch13* non-synonymous mutation, E433D, was observed in a day zero sample from Pawe.<br>No relation between the *Kelch13* mutation and reinfection. Almost every patient had a Day 3 negative, and hence, the relationship between mutation and parasitological response is difficult to assess. | L/M | Ethiopia is not represented in the IPDMA |

L= Low; L/M = Low to moderate; M/H= moderate/high

S Table 5.4: Assessment of bias due to studies not available for IPD-MA from S America (n=1)

| PubMed ID (Registration) | Study period | Country | Enrolled | Sampling frequency | Tested for K13 | Key K13 findings | Likely impact on IPD-MA | Justification for impact assessment |
|---|---|---|---|---|---|---|---|---|
| 32100686 | 2018-2019 | Colombia | 88 | - | 81 | A504D (n=1); all other were WT | L | Only assessed association with Day 3. Not applicable for PCE 1/2 assessment. |

L= Low
